# Polygenic basis and biomedical consequences of telomere length variation

**DOI:** 10.1101/2021.03.23.21253516

**Authors:** Veryan Codd, Qingning Wang, Elias Allara, Crispin Musicha, Stephen Kaptoge, Svetlana Stoma, Tao Jiang, Stephen E. Hamby, Peter S. Braund, Vasiliki Bountziouka, Charley A. Budgeon, Matthew Denniff, Chloe Swinfield, Manolo Papakonstantinou, Shilpi Sheth, Dominika E. Nanus, Sophie C. Warner, Minxian Wang, Amit V. Khera, James Eales, Willem H. Ouwehand, John R Thompson, Emanuele Di Angelantonio, Angela M. Wood, Adam S. Butterworth, John N. Danesh, Christopher P. Nelson, Nilesh J. Samani

**Affiliations:** Department of Cardiovascular Sciences, University of Leicester, Leicester, UK; NIHR Leicester Biomedical Research Centre, Glenfield Hospital, Leicester, UK; British Heart Foundation Cardiovascular Epidemiology Unit, Department of Public Health and Primary Care, University of Cambridge, Cambridge, UK; National Institute for Health Research Blood and Transplant Research Unit in Donor Health and Genomics, University of Cambridge, Cambridge, UK; British Heart Foundation Centre of Research Excellence, University of Cambridge, Cambridge, UK; School of Population and Global Health, University of Western Australia, Australia; Program in Medical and Population Genetics, Broad Institute of MIT and Harvard, Cambridge, MA, USA; Center for Genomic Medicine, Massachusetts General Hospital, Boston, MA, USA; Department of Medicine, Harvard Medical School, Boston, MA, USA; Cardiology Division, Department of Medicine, Massachusetts General Hospital, Boston, MA, USA; Division of Cardiovascular Sciences, University of Manchester, UK; Department of Haematology, University of Cambridge, Cambridge Biomedical Campus, Cambridge, UK; NHS Blood and Transplant, Cambridge Biomedical Campus, Cambridge, UK; Wellcome Sanger Institute, Wellcome Genome Campus, Hinxton, Cambridge, UK; Department of Health Sciences, University of Leicester, United Kingdom; Health Data Research UK Cambridge, Wellcome Genome Campus and University of Cambridge, Cambridge, United Kingdom; Medical Research Council Biostatistics Unit, Cambridge Institute of Public Health, University of Cambridge, Cambridge, United Kingdom; The Alan Turing Institute, London, United Kingdom; Department of Human Genetics, Wellcome Sanger Institute, Hinxton, United Kingdom

## Abstract

Telomeres, the end fragments of chromosomes, play key roles in cellular proliferation and senescence^1^. Here we characterize the genetic architecture of naturally-occurring variation in leucocyte telomere length (LTL) and identify causal links between LTL and biomedical phenotypes in 472,174 well-characterized participants in UK Biobank^2^. We identified 197 independent sentinel variants associated with LTL at 138 genomic loci (108 novel). Genetically-determined differences in LTL were associated with multiple biological traits, ranging from height to bone marrow function, as well as several diseases spanning neoplastic, vascular, and inflammatory pathologies. Finally, we estimated that at age 40 years, people with >1-SD shorter compared to ≥1-SD longer LTL than the population mean had 2.5 years lower life expectancy. Overall, we furnish novel insights into the genetic regulation of LTL, reveal LTL’s wide-ranging influences on physiological traits, diseases, and longevity, and provide a powerful resource available to the global research community.

## Introduction

Telomeres are nucleoprotein complexes at chromosome ends that shorten with each cell division, and play key roles in maintaining chromosomal integrity^1^. Telomere length (TL) is heritable, but there is incomplete understanding of its genetic determination^3-5^. Extreme shortening of telomeres − due to rare mutations in telomere regulatory genes − causes premature ageing syndromes^6^. By contrast, more subtle inter-individual variation in TL has been associated with risk of certain cancers, coronary artery disease and other common age-associated adult conditions^7-9^. As such, TL has been proposed as a biomarker of biological age^10^. Population biobanks afford opportunities to provide insight into the genetic architecture of TL and its links with biomedical phenotypes. Progress has been limited, however, because most biobanks have not been able to combine robust TL measurement, detailed genomic characterisation, extensive biomedical phenotyping and exceptional statistical power.

Here we interrogate a powerful population resource of peripheral leucocyte TL (LTL) measurements, a practicable measure of TL that correlates well with TL across different tissues^11^ within individuals, that we created in 472,174 well-characterized participants in UK Biobank (UKB)^12^. We increase knowledge of the genetic architecture of LTL several-fold, including identification of multiple novel rare and lower-frequency variants associated with LTL. Using the principle of Mendelian randomisation (MR), we find evidence to support causal roles for LTL with multiple physiological traits and diverse diseases. We also estimate that people with shorter LTL have lower life expectancy.

### Genetic determinants of telomere length

We used a well-validated qPCR assay to obtain LTL measurements in 472,174 UKB participants and undertook multiple quality checks to control and adjust for technical factors, as detailed elsewhere^12^. We also made paired LTL measurements from DNA taken at two time-points (mean interval: 5.5 years) in 1,351 participants to enable calculation of, and correction for, regression-dilution (**Methods**)^12^. Using standard genome-wide association analyses and exact joint conditional modelling in 464,716 participants with data available on 19.4 million imputed variants (minor allele frequency [MAF]≥0.1%) (**Methods & Supplementary Figure 1**), we identified 197 independent associations for LTL (**Supplementary Table 1**), exceeding a genome-wide significance threshold of p<8.31×10^−9^ (**Methods**). The sentinel variants were located within 138 genomic loci (>500Kb between sentinels), of which 108 were novel (>1Mb from a previously reported sentinel) and 30 previously reported at either genome-wide significance or false discovery rate (FDR)<5% (**Figure 1A, Supplementary Table 1, Supplementary Figures 2-3**).^4,5,13^ Collectively, the 197 variants explained 4.54% of the variance in LTL. In total, 714 independent variants, the majority of which are novel, were associated with LTL at an FDR threshold of <1% (**Supplementary Table 2**), increasing the amount of variance explained to 5.64%.

**Figure 1.**
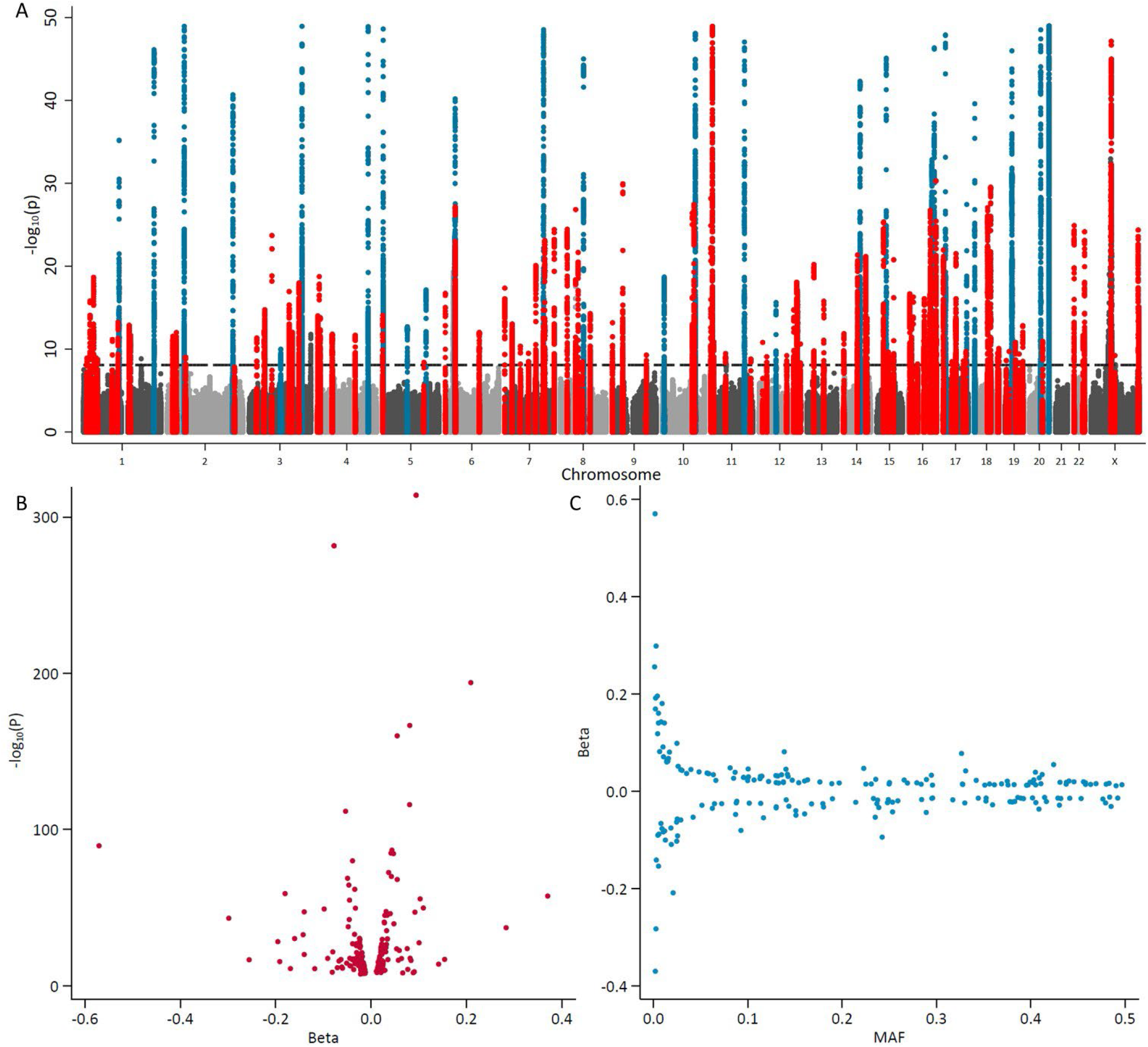
Conditionally independent genome-wide significant hits. A) Manhattan plot curtailed at p<1×10^−50^. We highlight the regions containing our 197 sentinel variants that are genome-wide significant (P<8.31×10^−9^; horizontal dashed reference line) in the exact joint conditional model (**Supplementary Table 1**). We define the region as known (blue) if a previous variant within 1Mb of our sentinel has previously been reported at either genome-wide significance or at a <5% FDR threshold. We consider our other regions novel (red) as the first evidence of a variant within 1Mb of our sentinel that reaches genome-wide significance. B) The estimated effect sizes (beta) against p-value from the GWAS. C) Estimated effect sizes (beta) against the minor allele frequency (MAF) from all participants in the GWAS.

Twenty of the genome-wide significant sentinel variants identified here for the first time (**Supplementary Table 1**) were lower frequency (MAF<1%), including novel association signals at several known loci (including *TERT1, TERF1*, and *RTEL1*) and novel loci (such as *EXOSC10, SMC4* and *SRSF6*). The estimated effects of sentinel variants were generally modest, i.e. less than 0.2 SDs per-allele (**Figures 1B** and **1C**). Most of the loci with the strongest evidence for association (p<1×10^−50^) were previously known but two are novel (**Supplementary Figure 2, Supplementary Table 1**): one on the X-chromosome not analysed in previous genome-wide association studies; the other rs334 in *HBB*, a variant known to cause sickle cell disease, predominantly seen in African ancestry individuals. As *HBB* was used as a control gene in our LTL assay, the fidelity of its apparent association with LTL is uncertain (**Supplementary Information, Supplementary Figure 4**). Except for rs334, none of the other associations were driven by inclusion of non-European ancestry participants (**Supplementary Table 1**).

Combining information on gene function, variant annotations and colocalising expression quantitative trait loci (eQTLs; **Methods, Supplementary Information, Supplementary Tables 3-6**), we were able to prioritise likely causal genes at 114 (83%) of the loci we discovered. Many biological candidates were supported by functional predictions and gene expression evidence, including strong eQTL support for *TEN1, STN1* and *RPA2* (**Supplementary Tables 4-6, Figure 2a**). Genes with known roles in telomere regulation were found in 44 loci, including genes encoding components of the SHELTERIN (*ACD (TPP1), TERF1, TERF2, POT1*), and CST complexes (*SNT1* (*OBFC1*), *TEN1, CTC1*), which act to cap the end of the telomere, suppressing inappropriate activation of the DNA damage response and regulating telomerase processivity (**Figure 2b**)^14^. Components of the alternative lengthening of telomeres (ALT) pathway (*ATRX, PML, SLX4*) were also among the novel loci as well as genes that post-translationally modify key telomere proteins, including *UPS7* which deubiquitinates both POT1 and ACD^15,16^. Genes within both known (*TERC, TERT, NAF1*) and novel (*DKC1, TEP1, SMG6, SHQ1, NOLC1, RUVBL1*) loci encode core components of proteins that regulate the assembly and activity of telomerase (**Supplementary Information**)^17-20^. Prior to telomerase assembly, *TERC* undergoes complex processing^21^. Genes involved in *TERC* stability, intra-cellular trafficking and processing were found in known (*SMUG1*) and novel loci (*PARN, TENT4B (PAPD5), TGS1, WRAP53*), including those associated with the RNA exosome (*EXOSC6, EXOSC9, EXOSC10, DIS3, ZCCHC8*) (**Figure 2b, Supplementary Information**)^21-24^.

**Figure 2.**
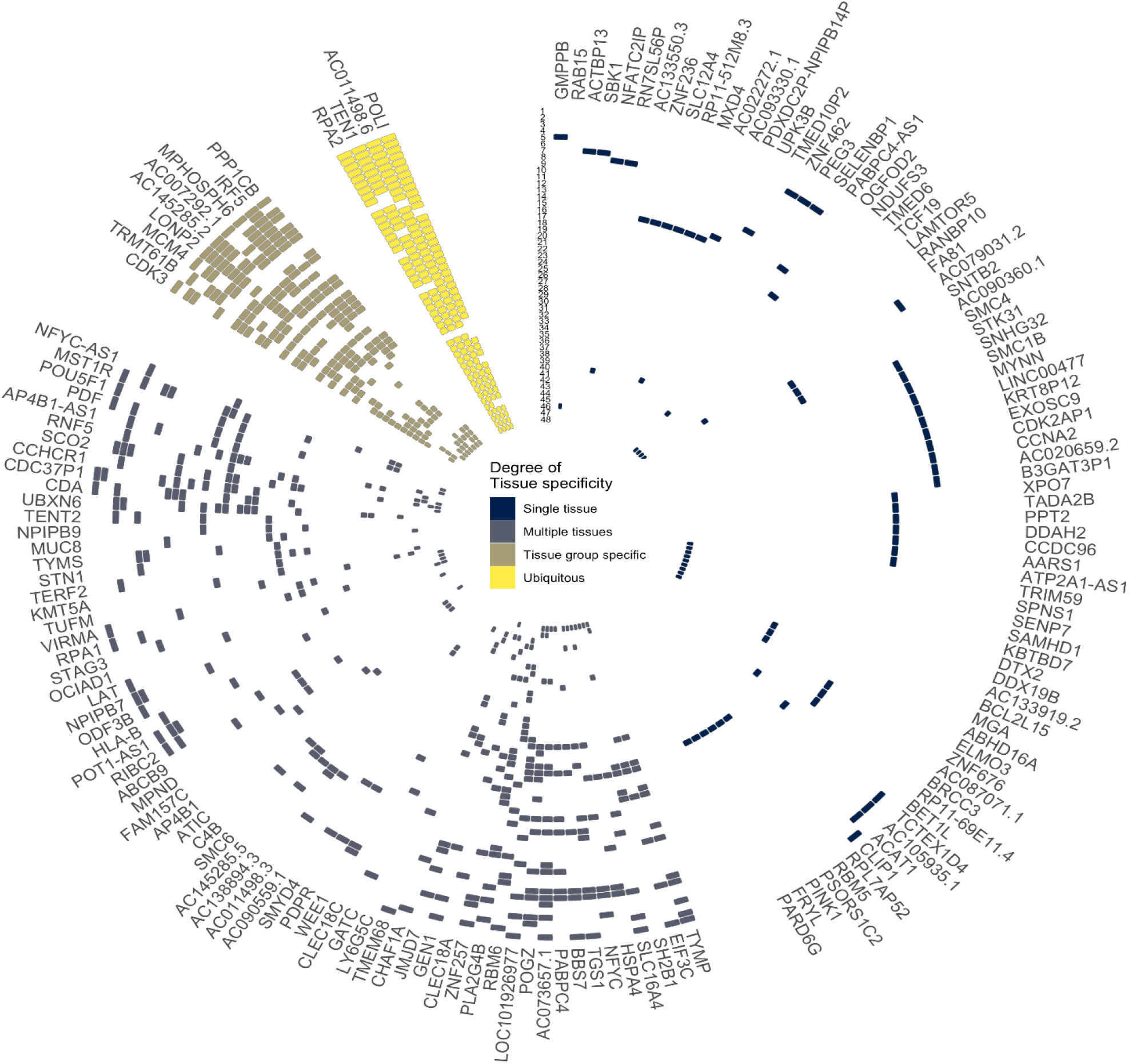
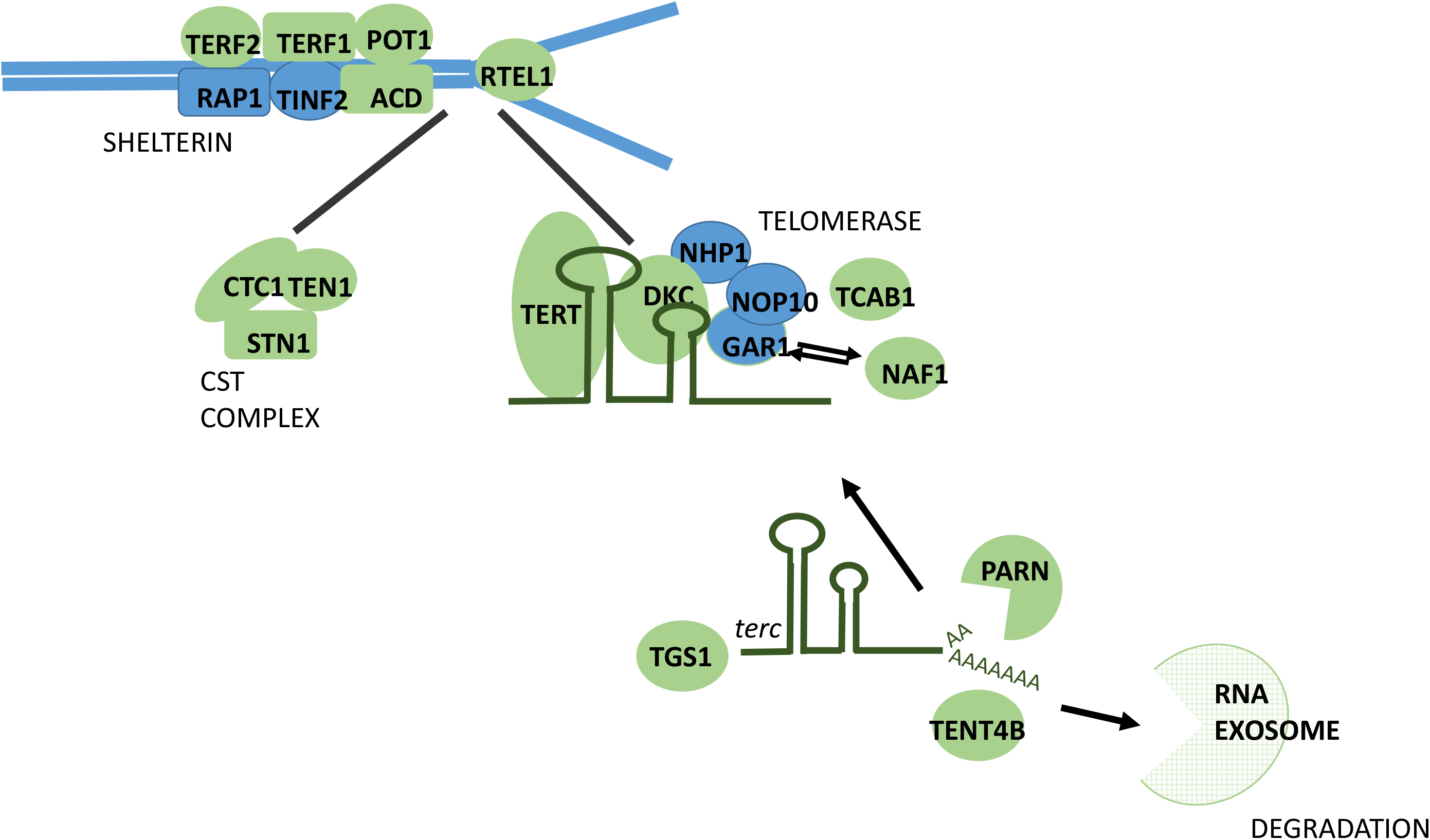
**Identification of eQTL signals at genome-wide significant loci and those with known regulatory roles in telomere maintenance.** A) Circular representation of colocalised eQTLs across 48 tissues in GTEx. Data for strong colocalisation is shown as a coloured tile, with the colour determined by the degree of tissue specificity of colocalisation: “Ubiquitous colocalisation” (yellow; greater than 32 tissues), “Tissue group specific” (brown; greater than 16 and less than 32 tissues), “Multiple tissues” (grey, greater than 1 and less than 16 tissues) and “Single tissue” (dark blue; a single tissue). Tissues are numerically with full details in Supplementary Table 5). Genes are labelled using HGNC gene symbols. Tissues are ordered by GTEx tissue groupings and genes are ordered by hierarchical clustering of the data, which groups genes with a similar colocalisation pattern. B) Key components of telomere regulatory complexes found within genome-wide significant loci. Proteins depicted in green are found within GWAS loci, those not found within GWAS loci are depicted in blue. We find the majority of components of core telomere binding complexes alongside many proteins involved in the formation and activity of telomerase. Note that not all components of the RNA exosome are shown.

Other genes of interest in novel loci are involved in DNA replication, recombination and repair. Two novel loci harbour components of the Replication Protein A complex (*RPA1, RPA2*), which is recruited to telomeres during DNA replication^25^. The complex is later removed in a process involving hnRNPA1 (within the *SMUG1* locus) and replaced by POT1^26^. DNA double strand break repair genes with known roles in telomere regulation were also observed (*SLX4, MCM4, SAMHD1)*^27,28^. Two other genes highlighted as likely causal are *POLI* and *POLN*. Neither is known to have a direct role in telomere maintenance; however, other DNA polymerases involved in translesion repair function in the ALT pathway^29^.

To provide more evidence for the candidacy of our prioritised genes, we investigated whether rare (<0.1% MAF) protein-altering variants in these genes were associated with LTL using gene-based tests (**Supplementary Methods**). The aggregated scores for eight genes (*RTEL1, TERF1, TERT, ATM, PARN, SAMHD1, POT1, CTC1*) were significantly associated with LTL after Bonferroni correction (**Supplementary Table 7**). The directions of association with LTL for the individual variants included in this analysis are consistent with the known biological functions of these genes in telomere regulation (**Supplementary Table 8, Supplementary Information**). For example, carriers of rare protein-truncating/altering variants throughout *RTEL1* were mostly associated with shorter LTL, consistent with data suggesting that the full length RTEL1 protein is required to facilitate telomere elongation by telomerase^30^.

To identify potentially novel pathways responsible for telomere length regulation, we tested for over-representation of prioritised genes in known biological processes (**Methods**). As expected, the most significantly associated pathways identified were related to the regulation of telomere maintenance. Two other enriched pathways, box H/ACA snoRNP assembly and snoRNA 3’-end processing, also highlight key components of TERC regulation within the associated loci. Extending our previous identification of the relevance of nucleotide metabolism to LTL,^4^ the current analysis more specifically prioritised pyrimidine metabolism (**Supplementary Table 9**).

An additional motivation for undertaking the genome-wide association study was to create genetic instruments to enable causal inference analysis of LTL with biomedical phenotypes. To minimise inclusion of correlated variants or those showing extensive pleiotropy in these analyses, we filtered the 197 sentinel variants further (**Methods**), yielding 131 conditionally independent, uncorrelated and “non-pleiotropic” genome-wide significant instruments used in the MR analyses described below (**Supplementary Table 1**).

### Influences on biomedical traits

Partly guided by previous reports (**Supplementary Table 10**), we prioritised 93 biomedical traits available in UKB, comparing MR results with observational results based on LTL levels corrected for the observed regression-dilution ratio of ∼0.65 (abbreviated “usual LTL”) (**Supplementary Methods**). We focused mainly on continuous traits related to body shape and size, cardiorespiratory function, reproductive health, physical fitness, bone marrow function, cognition, bone health and liver and endocrine function (**Supplementary Table 10**). After Bonferroni correction, 18 of the traits were significantly associated in the same direction with both genetically-determined LTL and usual LTL (**Figure 3A and Supplementary Table 10**). Genetically-determined LTL was more strongly related to most traits than usual LTL, likely reflecting lifelong influences (**Supplementary Table 10**). However, for all traits LTL explained only a small proportion of the variance (<0.5%). For an additional 12 traits we found nominally significant associations (p<0.05) with genetically-determined LTL, with most of these traits showing significant and concordant associations with usual LTL (**Figure 3A and Supplementary Table 10**). A further 38 traits showed Bonferroni-significant observational associations but no associations with genetically-determined LTL (**Supplementary Figure 5 and Supplementary Table 10**). Lack of concordance for these traits could reflect either residual bias in observational analyses or limited statistical power in MR analyses.

**Figure 3.**
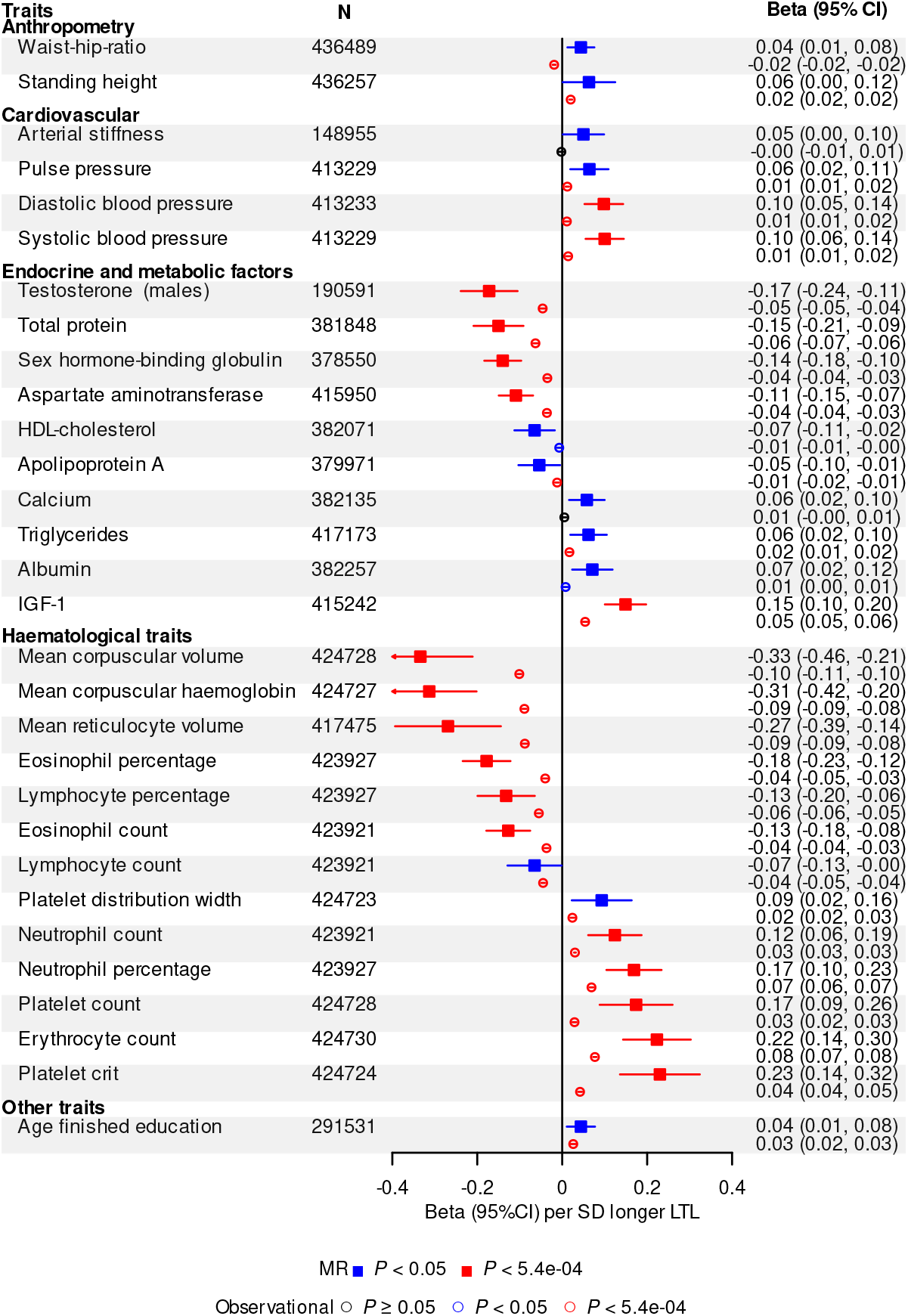
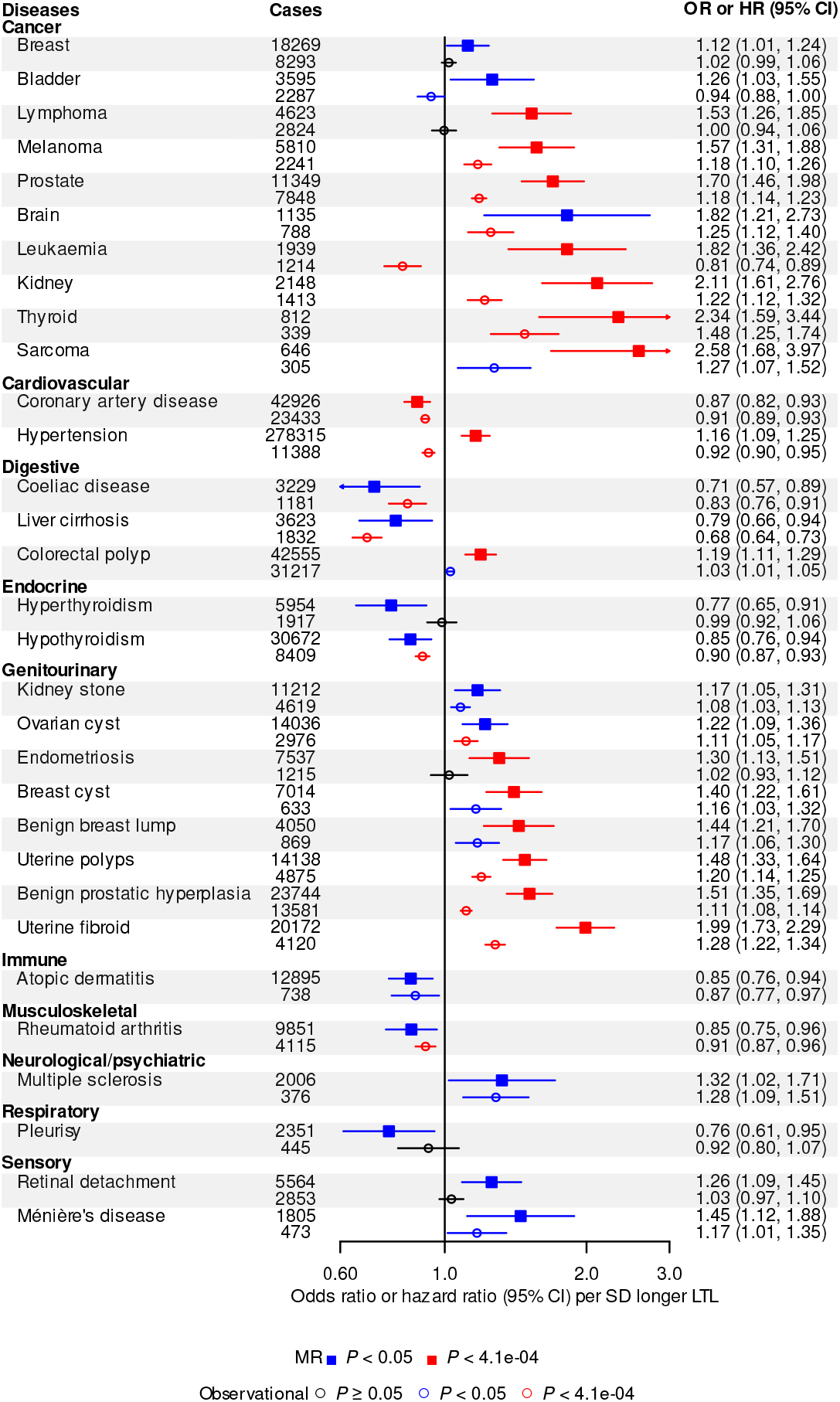
Biomedical traits and diseases associated with genetically-determined LTL. A) Biomedical trait Mendelian randomisation (MR) associations are shown with a solid square and expressed in beta per standard deviation (SD) longer genetically-determined leucocyte telomere length (LTL). Observational associations are shown with an empty circle and expressed in beta per SD longer usual LTL. B) Disease MR associations are shown with a solid square and expressed in odds ratio (OR) per SD longer genetically-determined LTL. Observational associations are shown with an empty circle and expressed in hazard ratio (HR) per SD longer usual LTL. CI, confidence interval.

Overall, our findings demonstrate that variation in LTL affects a wide range of biological and physiological traits, spanning multiple body systems. We confirmed associations of genetically-determined longer LTL with higher blood pressure and identified novel associations with circulating biomarkers of metabolic and endocrine function, including higher insulin-like growth factor 1 (IGF-1), and lower sex hormone binding globulin (**Figure 3A and Supplementary Table 10**). IGF-1 is a growth hormone associated with the pubertal growth in height. Adjusting for IGF-1 levels attenuated the association between height and longer usual LTL (beta=0.012 (0.010, 0.014), p=1.91×10^−27^), suggesting IGF-1 may partly mediate the relationship between LTL and height. Notably, we observed associations between genetically-determined LTL and multiple haematological traits (**Figure 3A**). Associations of longer LTL with higher counts of neutrophils, platelets and erythrocytes but lower counts of lymphocytes and eosinophils (**Figure 3A and Supplementary Figure 5**) suggest an effect of LTL variation on lineage fating at the lympho-erythromyeloid progenitor level^31^. The contrasting associations of LTL with erythrocyte count versus erythrocyte size and haemoglobin content may reflect a primary effect on maintenance of red cell mass^32^. However, for platelets, longer LTL was not only associated with a higher count, but also larger volume, resulting in an increased platelet mass (**Figure 3A and Supplementary Figure 5**), consistent with recent observations that megakaryocytes, platelet precursor cells that reside in the bone marrow, originate directly from megakaryocyte-primed haematopoietic stem cells^33^ and not from a precursor cell clonally related to erythroid precursors.

### Influences on disease outcomes

To identify causal links between LTL and disease outcomes using MR analyses, we prioritised 123 diseases defined using information available in UKB (**Supplementary Table 11, Supplementary Information**). We compared results from MR analyses with observational Cox regression analyses of incident cases. After Bonferroni correction, 16 of the diseases were significantly associated with genetically-determined LTL (**Figure 3B, Supplementary Table 12**). We confirmed associations of longer genetically-determined LTL with *lower* risk of coronary artery disease, as well as with *higher* risk of several organ-specific cancers, including prostate, melanoma, thyroid and kidney, and genitourinary tumours (uterine polyps and fibroids)^4,34^. We found novel associations of longer genetically-determined LTL with *higher* risk of sarcoma (a malignant tumour of connective or haematopoietic tissues) and endometriosis (the growth of endometrial tissue outside of the uterus). Results were consistent across sensitivity analyses, suggesting robustness to horizontal pleiotropy (**Supplementary Figure 6**).

Of the 16 diseases significantly associated with genetically-determined LTL, twelve were also Bonferroni-corrected or nominally (p< 0.05) significantly associated with usual LTL in the same direction (**Figure 3B**). For most conditions causally linked to LTL, we identified approximately log-linear dose-response relationships of usual LTL with incident outcomes (**Supplementary Figure 7**). As for biomedical traits, we found genetically-determined LTL was more strongly related to diseases than usual LTL (**Figure 3B**). For two conditions (leukaemia and hypertension), we observed significant results in opposing directions for MR and observational analyses (**Figure 3B**). For leukaemia, it was likely due to a U-shaped association with usual LTL (**Supplementary Figure 7**); for hypertension it was likely due to residual bias in the observational analysis (**Supplementary Figure 8, Supplementary Table 13, Supplementary Information**). We did not find evidence that blood pressure or plasma lipid levels (high density lipoprotein-cholesterol, low density lipoprotein-cholesterol, triglycerides) explained the association between shorter genetically-determined LTL and higher risk of coronary artery disease.

For an additional 15 diseases we found nominally significant (p<0.05) associations with genetically-determined LTL (**Figure 3B**). Of these, 10 also had Bonferroni or nominally significant and concordant associations with usual LTL (**Figure 3B**), suggesting future more powerful MR studies may strengthen the evidence for causality. These included novel associations with liver cirrhosis, kidney stones and atopic dermatitis. For 26 diseases, we found Bonferroni-significant associations with usual LTL but non-significant associations with genetically-determined LTL (**Supplementary Figure 9, Supplementary Table 12**). These findings could reflect either residual bias in the observational analysis or limited power in the MR analyses (**Supplementary Information**). Finally, for 66 diseases we found no association in either MR or observational analyses (**Supplementary Table 12**).

### Influences on life expectancy

Given the causal links between LTL and multiple conditions − both in risk-increasing and risk-reducing directions − a relevant unresolved question is whether LTL has a net impact on life expectancy^35-37^. Using public health modelling methods previously described that draw on cause-specific mortality rates from the general population (**Methods**), we estimated that at age 40 years men with >1-SD shorter compared to ≥1-SD longer telomeres than the population mean had 2.47 (95% CI: 1.99-2.96) years lower life expectancy (**Figure 4A**); the corresponding estimates for women were very similar. These estimated differences were sustained to age 65 years, and gradually declined thereafter. Excess cardiovascular deaths accounted for 13% and 9% and cancer deaths 5% and 4% of the survival difference in men and women respectively, with most of the remainder due to other causes (**Figure 4B**). Broadly similar results were observed in sensitivity analyses that involved different modelling assumptions (**Supplementary Figure 10**).

**Figure 4.**
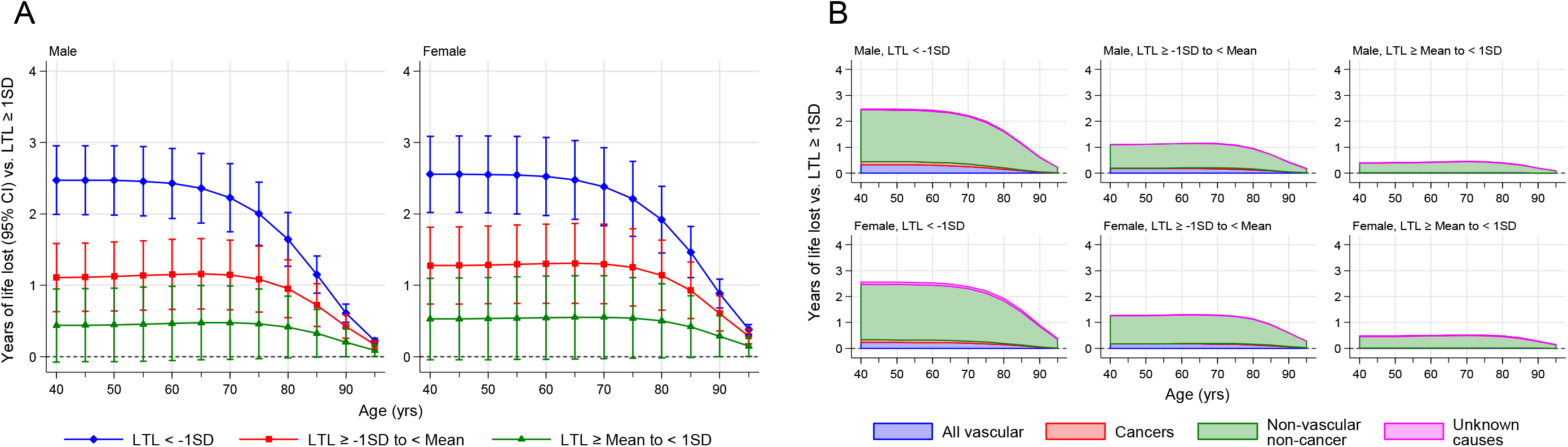
Years of life lost using UK 2015 mortality rates. Years of life lost were estimated for four standardised LTL groups: group 1 (<-1SD), group 2 (−1 to <0 SD), group 3 (0 to <1SD), and group 4 (≥1SD) from 40-95 years of age. Group 4 was used as the reference group. Data is shown for males and females separately. This was performed for all-cause mortality (A) and disease specific mortality (B).

## Discussion

This study elucidates the polygenic basis and biomedical consequences of LTL variation. In the most powerful genomic study so far, we reveal many novel candidate genes, highlight the complex regulation of LTL, and identify a role for pyrimidine metabolism. Using wide-angle analyses, we provide insight into the causal relevance of LTL to biological traits and diseases across multiple body systems, comparing genetic and observational associations *in the same set of participants*. There is much interest in shorter TL as a target for pharmacological and other interventions^38,39^. Two findings from our analyses provide insight into this issue. First, for coronary artery disease and most other conditions causally linked with shorter LTL, we found continuous linear associations, i.e. no threshold above which LTL stops being associated with risk indicating that any benefits could accrue across the range of TL. On the other hand, the observation that longer LTL is causally associated with risk of several cancers − possibly because longer telomeres allow more cell divisions and clonal expansion after first hit cancer mutations, thereby increasing the likelihood of second hit mutations that drive oncogenic transformation^40^ − highlights the complexity of TL as a therapeutic target.

Our results suggest that, despite LTL’s directionally opposing associations with different diseases, shorter LTL at age 40 years is on average associated with about 2.5 years lower life expectancy. For comparison, estimated reductions in life expectancy from long-term cigarette smoking or having diabetes in mid-life are about 10 years and 6 years, respectively^41,42^. Overall, our study provides a major resource for understanding the relevance of LTL to many complex diseases and traits.

## Data Availability

Source data is accessible via application to UK Biobank. Summary statistics of the GWAS are available on request to the corresponding authors.

## Online Methods

### Leucocyte telomere length measurements

The measurement of leucocyte telomere length (LTL) in UK Biobank (UKB) participants and the extensive quality checks and adjustment for technical factors are detailed elsewhere^12^. For the analyses presented in this paper, we included all participants with LTL measured from a UKB baseline sample where there was no mismatch in self-reported and genetic sex (n=472,174: data-freeze December 2020). LTL values were log-transformed and Z-standardised for all analyses.

### Genome-wide association study (GWAS)

We used imputed genotypes available in UKB^2^ for the GWAS. To assure quality, we restricted the analysis to variants with a minor allele frequency (MAF) ≥ 0.1% (where imputation accuracy is greatest) and an INFO score ≥ 0.3. We tested 19.4 million variants using the BOLT-LMM package, adjusting for age, gender, array and the first 10 principal components (PCs). The analysis was run separately for chromosome 23, where males were coded as 0/2.

### Conditional association analyses

To identify independently associated variants within loci, we adopted a two-stage approach to conditional analyses. We first used the summary statistics for variants meeting a threshold of P<1×10^−6^ from the GWAS to perform joint-conditional analysis using GCTA (Version 1.25.2, see **Supplementary Methods**). We set a genome-wide significance threshold at P<8.31×10^−9^ which has been suggested as an appropriate threshold for GWAS studies incorporating lower frequency and rare variants (MAF>0.1%)^43^. All variants with P<8.31×10^−9^ were then taken forward to stage two. In the second stage we performed exact joint modelling using BOLT-LMM where we adjusted for all other variants from stage one, age, sex, genotype array, and the first 10 PCs in the model. All variants emerging from this analysis with P<8.31×10^−9^ were considered to be conditionally independent at the genome-wide significance level.

### Variance explained by the genetic variants

To estimate the variance explained by all conditionally independent genome-wide significant variants, we extracted them from the imputed genetic data, scored by allele dosage. We only included participants that had both autosome and X-chromosome data. To account for familial correlation we randomly excluded one participant from each related pair, where a pair were related if the kinship coefficient was K>0.088 estimated using genetic relatedness^2^. A linear regression, adjusted for age, sex, array and the first 10 genetic PCs was run to estimate the model R^2^. A second model including all genetic variants was then run to estimate the full model R^2^ with the difference in model R^2^ used to determine the variance explained by the genetic variants.

To determine variants that pass a false discovery rate (FDR),^44^ we estimated the P-value equivalent to a q-value<0.01 as FDR_P<3.9×10^−5^. All variants from the GWAS with P<1×10^−4^ were tested using GCTA (**Supplementary Methods**) to identify conditionally independent variants that pass our FDR_P threshold. These were then clumped using PLINK to include only independent variants not in linkage disequilibrium (LD, R^2^<0.01). The remaining variants were then extracted and modelled as above to estimate the variance explained by the FDR set.

### Identification of potential causal variants

To identify putative causal variants allowing for multiple putative causal variants within a locus, we performed a shotgun stochastic search with FINEMAP V1.4^45,46^. For each locus, we calculated the posterior probability of causal configurations and report the most probable set. First, we defined a region to contain all variants within a 1 Mb window centred on each sentinel SNP. We identified the top causal variant for each region and identified all regions harboured within multiple sentinel GWAS loci. Initially we specified there to be only one causal variant. We then grouped the regions in multi-lead-SNP GWAS loci by locus (containing *k* lead SNPs within the 1Mb region). Then we allowed for a maximum of *i* causal variants (*i=k+3*). If the maximum posterior probability (PP_*icvar*_) for having *i* causal variants in the region was ≤95%, we selected the causal configuration and then generated credible sets. If PP_*icvar*_>95%, we further allowed for a maximum of *j* causal variants (*j=i+3*) and selected the causal configuration that has the largest PP_*icvar*_ closest to 95%. However, if PP_*icvar*_ was very low, the single causal configuration was selected (**Supplementary Table 14**).

### Identification of likely causal genes

To identify potential causal genes within the associated loci, we identified genes with known roles in telomere regulation (candidate genes) and used information from variant annotation in eQTL colocalisation analyses. Functional annotation for all variants identified within the 95% credible sets produced from fine-mapping was collected using VEP^47^ (**Supplementary Methods**).

To investigate whether the variants included within the 95% credible sets for each locus identified using FINEMAP share a common causal variant with eQTL signals, we conducted colocalisation analyses using COLOC^48^. Transcriptomic data were obtained from GTEx.v7 for genes with q-value <0.5 for all 48 tissues^49^. The COLOC method uses an approximate Bayes factor with both GWAS and eQTL summary statistics and regional LD structure to estimate posterior probabilities for five scenarios (PP0, PP1, PP2, PP3, and PP4). A high PP4 indicates evidence of a shared single causal variant. For each of the GWAS signals, we defined a 1 Mb region centred on the sentinel variant to test for colocalisation using the COLOC R package (https://cran.r-project.org/web/packages/coloc/vignettes/vignette.html). We defined *strong* evidence of colocalisation as PP3+PP4≥0.99 and PP4/PP3≥5 and *suggestive* evidence as PP3+PP4≥0.90 and PP4/PP3≥3, as previously described^4,50^.

Genes were prioritised on strength of evidence in the following order: biological candidate > high impact annotation > moderate impact annotation > strong evidence of colocalisation > suggestive evidence of colocalisation. Where expression of multiple genes was associated with our causal variants, we prioritised candidacy based on the number of tissues with evidence. To run downstream pathway analysis and gene-based tests where it was not possible to prioritise a gene at a locus, we substituted the nearest gene to the most significantly associated causal variant. Conversely, where it was not possible to prioritise a single gene from several with evidence, multiple genes were taken forward.

### Pathway analysis

We tested our list of prioritised or nearest genes for statistical overrepresentation testing (Fisher’s exact test) in PANTHER^51^. Pathways within the GO biological process complete annotation set were considered to be significantly over-represented at FDR q-value<0.05.

### Gene-based tests

From the prioritised genes identified in the GWAS loci, we removed non-coding RNAs, pseudogenes and poorly annotated novel transcripts. We then extracted rare and ultra-rare variants (MAF<0.1%) within exon boundaries of these genes from the UKB exome sequencing data^52^. Protein-altering variants were scored as predicted high-confidence loss-of-function and ultra-rare missense variants based on annotation obtained from VEP using the VEP LOFTEE plugin (**Supplementary Methods**)^47,53^. For each participant, the gene-specific score was obtained by aggregating the variant scores, capped at 1 (**Supplementary Methods**). We tested the association between the gene-specific scores and LTL using linear regression implemented in R v.4.0.0 adjusting for age and sex. To support this we ran single variant analyses of the rare variants using PLINK v1.9, also adjusting for age and sex.

### Genetic instruments for Mendelian randomisation analysis

Starting with the 193 sentinel variants located on the autosomes, we removed correlated variants from loci with more than one conditionally independent variant by removing those with R^2^>0.01 using PLINK clumping with LD based on the same randomly selected UKB sample as for the conditional association analysis. We removed the *HBB* locus due to potential technical artefacts (**Supplementary Information**). To remove potentially pleiotropic loci, we investigated the remaining 147 variants for association with multiple traits and phenotypes using previously curated data^54^. For each variant we derived the number of associations within different biological domains and defined evidence of pleiotropy as associations within at least 3 different domains. This led to the selection of 131 conditionally independent, uncorrelated and “non-pleiotropic” genome-wide significant instruments that we used for all MR analysis (**Supplementary Table 1**).

### Mendelian Randomisation

With our genetic instruments for LTL we performed single-sample univariable MR using two-sample methods that have been shown to be robust in large scale biobanks^55^. We used: (i) the inverse-variance weighted method for LTL based on all 131 independent and uncorrelated variants associated with LTL^56^; (ii) MR-Egger regression to estimate unmeasured pleiotropy^57^; (iii) weighted median estimator to assess the robustness to extreme SNP-outcome associations^58^ and (iv) a contamination-mixture method to explore potential presence of multiple pathways^59^. To account for between-variant heterogeneity, we used multiplicative random-effects models in all analyses and quantified heterogeneity using the I-squared statistic from the MendelianRandomization package v. 0.5.0 (https://CRAN.R-project.org/package=MendelianRandomization).

### Analysis of biomedical traits

To assess the influence of LTL on biomedical traits we were partly guided by previous reports (**Supplementary Table 12**) in our prioritisation of 93 biomedical traits, focusing only on continuous and binary outcomes. Continuous traits were first winsorized at the 0.5% and 99.5% percentile values to account for potentially influential outliers. After checking the distribution of the winsorized traits using histograms, natural log-transformations were applied to non-normally distributed traits where appropriate. All continuous biomedical traits were then scaled to the Z-standardised normal distribution. To account for familial correlation, we randomly excluded one from each related pair, where a pair were related if the kinship coefficient was K>0.088.

We used MR to investigate causal associations of LTL with biomedical traits. To estimate the genetic associations for each of our 131 genetic instruments with each biomedical outcome, we performed logistic regression for binary traits, and linear regression for continuous traits, adjusting for age, sex, array and the first five genetic PCs using SNPTEST^60^. We then used MR to investigate causal associations of LTL with biomedical traits and ran MR sensitivity analyses (**Supplementary Figure 11**).

Observational analyses were conducted to investigate the association between LTL and biomedical traits. The Z-standardised LTL was used as the predictor of interest to provide effect size estimates for a 1 standard deviation increase in LTL. Continuous traits were assessed using linear regression models, while logistic regression models were used for binary traits. All regression models were adjusted for age, sex, ethnic group (defined by UKB as: Asian, Black, Chinese, Mixed, Other and White) and white blood cell (WBC) count as proposed elsewhere^12^. To correct for measurement error and within-person variability in LTL over time, observational associations of LTL with traits and diseases were corrected for the observed regression-dilution ratio of 0.65, as detailed elsewhere^12^. Observational associations relate to *usual* LTL unless otherwise specified. The magnitude of association was estimated using a partial R^2^, calculated as the difference between the full model R^2^ and the model R^2^ leaving LTL out.

To assess non-linear associations between LTL and the traits, a quadratic term (the squared value of the LTL) was included in separate models on top of LTL, age, sex, ethnicity and WBC. We further assessed non-linear associations of LTL with various traits by fitting fractional polynomial models (**Supplementary Figure 12**) adjusted for age, WBC, sex and ethnic group. The best fitting fractional polynomial model, selected using P<0.05 as evidence for selecting more complex non-linear functions, was used to plot the continuous shape of association relative to the reference value of 0 ^61^. In further supplementary analyses we calculated adjusted hazard ratios by deciles of LTL and plotted them against the mean standardised LTL within deciles.

### Analysis of disease outcomes

We identified 123 diseases (**Supplementary Table 8**) using a slightly modified version of the strategy reported previously^4^ (**Supplementary Methods**). The selection of diseases aimed to balance the needs for clinical relevance (e.g., avoiding overlapping outcomes i.e. CAD and MI), detail (to cover diseases with different physiopathology) and statistical power. We conducted power calculations due to the large differences in disease prevalence using the ‘powerLogisticCon’ function from the R package powerMediation^62^. These power calculations show (**Supplementary Figure 13**) that all outcomes have at least 60% power to detect an odds ratio (OR) of 1.1 at the 5% level of significance. Around 75% of our disease outcomes, based on prevalence, had >99% power to detect an OR of 1.1, with 60% of our outcomes having >99% power to detect an OR of 1.05. To account for familial correlation we randomly excluded one participant from each related pair, where a pair were related if the kinship coefficient was K>0.088.

Using a combination of prevalent and incident diseases (**Supplementary Methods**) we estimated the genetic associations with each disease outcome using logistic regression. We then performed MR using these estimates as for the biomedical traits described above. For the observational associations, time-to-event analyses were conducted between z-standardised LTL and incident disease using Cox proportional hazards models, stratified by sex and ethnicity and adjusted for age and WBC. For this analysis, participants with prevalent disease at baseline were excluded. In order to test the proportional hazards assumption we fit an interaction term between LTL and time. For any deviations from proportional hazards (time interaction P<0.05), we estimated the hazard ratios (HRs) at baseline and at 10 years via linear combination. We performed these analyses using the survival (https://CRAN.R-project.org/package=survival) and greg (https://CRAN.R-project.org/package=Greg) packages in R. Estimated log(HRs) and corresponding confidence intervals were adjusted using the regression dilution ratio of 0.65^12^.

To investigate reasons for any discrepancies between MR and observational results, we performed MR analysis using only incident disease outcomes, and observational analyses using logistic regression with incident data and with prevalent data. Shapes of associations were assessed using fractional polynomials^63^ with Cox regression models adjusted for age, WBC, and stratified by sex and ethnic group. The best fitting model was selected in the same way as for the biomedical trait analysis.

### LTL and longevity

Details of the methods used to estimate differences in life expectancy have been previously described^64^ with further specific information regarding the modelling for LTL provided in **Supplementary Methods**. Briefly, estimates of cumulative survival from 40 years of age onwards among 4 groups of z-standardised measured LTL: group 1 (<-1SD), group 2 (−1 to <0 SD), group 3 (0 to <1SD), and group 4 (≥1SD), the reference group, were calculated by applying HRs for cause-specific mortality calculated from the UKB study (specific to age-at-risk and stratified by sex) to population mortality rates for the UK and European Union (EU) in 2015 (by sex and 5-year age groups). Calculations were performed giving specific consideration to interpreting estimated differences in life expectancy between group 1 (i.e. shorter telomeres) and group 4 (i.e. longer telomeres) from age 40 years. Analyses involved Stata version 14.0 (StataCorp), 2-sided P values, and used a significance level of P < 0.05.

## Acknowledgements

This research has been conducted using the UK Biobank Resource under Application Number 6077 and was funded by the UK Medical Research Council (MRC), Biotechnology and Biological Sciences Research Council and British Heart Foundation (BHF) through MRC grant MR/M012816/1. C.P.N is funded by the BHF (SP/16/4/32697). V.C., C.A.B., C.M., V.B., Q.W., R.B., C.P.N. and N.J.S. are supported by the National Institute for Health Research (NIHR) Leicester Cardiovascular Biomedical Research Centre (BRC-1215-20010). Cambridge University investigators are supported by the B.H.F (RG/13/13/30194; RG/18/13/33946), Health Data Research UK, NIHR Cambridge Biomedical Research Centre (BRC-1215-20014), NIHR Blood and Transplant Research Unit in Donor Health and Genomics (NIHR BTRU-2014-10024) and MRC (MR/L003120/1). J.D. holds a BHF Personal Professorship and NIHR Senior Investigator Award. A.M.W. and E.A. received support from the EU/EFPIA Innovative Medicines Initiative Joint Undertaking BigData@Heart (11607). Parsa Akbari, Thomas Bolton and Matthew Arnold made computational and biostatistical contributions to this work.

## Author Contributions

M.D., C.S, M.P., S.Sh., D.E.N. and V.C. generated and curated the data. Q.W., T.J., V.C., A.S.B. and C.P.N. performed the GWAS analyses. Q.W., S.E.H., X.W., A.V.K., V.C. and C.P.N. performed the rare variant analyses. Q.W., P.S.B., J.E., and V.C. conducted downstream annotations. C.M., V.B., P.S.B., V.C. and C.P.N. performed biomedical trait analyses. E.A., S.St., S.K., E.D.A. and C.P.N. performed disease analyses. S.K. and A.M.W. performed life expectancy analyses. V.C., C.P.N., Q.W., E.A., C.M., S.K., S.St., V.B., W.H.O., E.D.A., A.M.W., A.S.B., J.R.T., J.N.D. and N.J.S. prepared the manuscript and all authors revised it. V.C., C.P.N., J.R.T., J.N.D. and N.J.S. (Principal investigator) secured funding and oversaw the project.

## Competing Interests Declaration

The authors declare no competing interests

Supplementary Information is available for this paper.

Correspondence and requests for materials should be addressed to Veryan Codd or Nilesh J. Samani

