## Supplementary Figure 3 for "Polygenic basis and biomedical consequences of telomere length variation"

Supplementary Figure 3. regional association plots for GWAS sentinels.

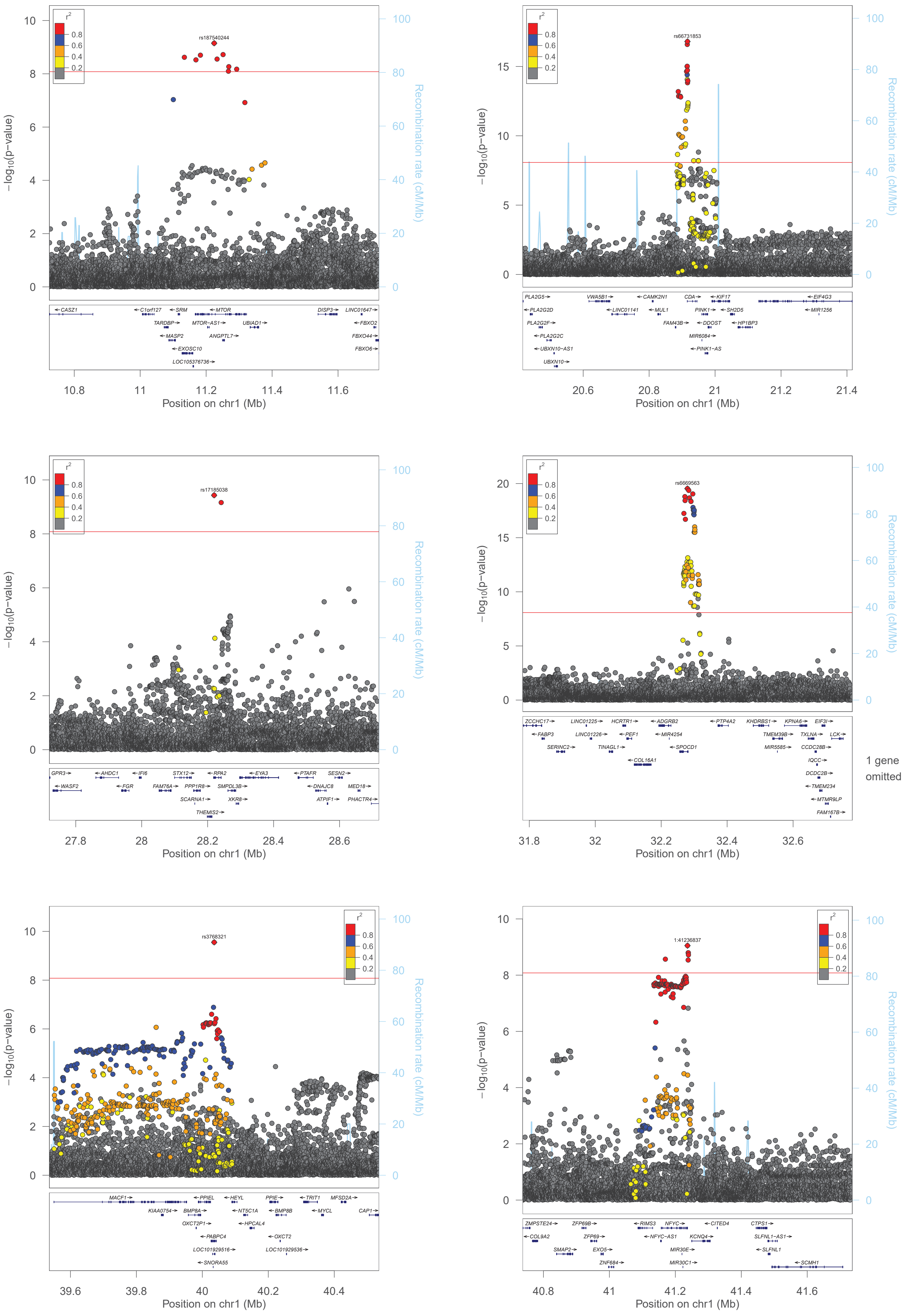

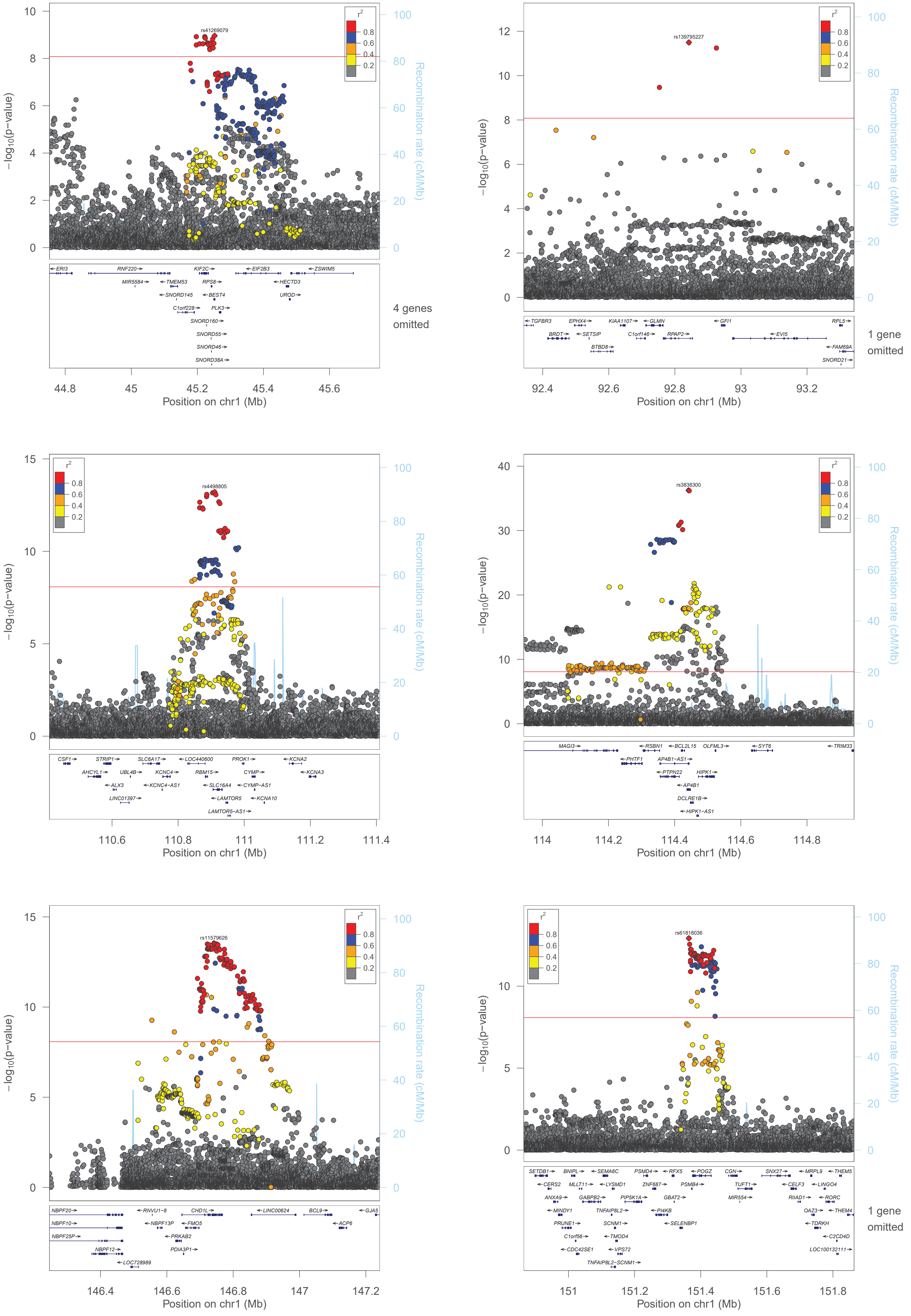

−log10(p−value)
Recombination rate (cM/Mb)
r2
0.8
0.6
0.4
0.2
rs41269079
ERI3
RNF220
KIF2C
EIF2B3
ZSWIM5
MIR5584
TMEM53
RPS8
HECTD3
SNORD145
BEST4
UROD
C1orf228
PLK3
SNORD160
SNORD55
SNORD46
SNORD38A
4 genes omitted
Position on chr1 (Mb)
rs139795227
TGFBR3
EPHX4
KIAA1107
GLMN
GFI1
RPL5
BRDT
SETSIP
C1orf146
RPAP2
EVI5
BTBD8
FAM69A
SNORD21
1 gene omitted
rs4498805
CSF1
STRIP1
SLC6A17
LOC440600
PROK1
KCNA2
AHCYL1
UBL4B
KCNC4
RBM15
CYMP
KCNA3
ALX3
KCNC4−AS1
SLC16A4
CYMP−AS1
LINC01397
LAMTOR5
KCNA10
LAMTOR5−AS1
rs3838300
MAGI3
RSBN1
BCL2L15
OLFML3
SYT6
TRIM33
PHTF1
AP4B1−AS1
PTPN22
HIPK1
AP4B1
DCLRE1B
HIPK1−AS1
rs11579626
NBPF20
RNVU1−8
CHD1L
LINC00624
BCL9
GJA5
NBPF10
NBPF13P
FMO5
ACP6
NBPF25P
PRKAB2
NBPF12
PDIA3P1
LOC728989
rs61818036
SETDB1
BNIPL
SEMA6C
PSMD4
RFX5
POGZ
CGN
SNX27
MRPL9
THEM5
CERS2
MLLT11
LYSMD1
ZNF687
PSMB4
TUFT1
CELF3
LINGO4
ANXA9
GABPB2
PIP5K1A
GBAT2
MIR554
RIIAD1
RORC
MINDY1
TNFAIP8L2
PI4KB
OAZ3
THEM4
PRUNE1
SCNM1
SELENBP1
TDRKH
C1orf56
TMOD4
C2CD4D
CDC42SE1
VPS72
LOC100132111
TNFAIP8L2−SCNM1
1 gene omitted

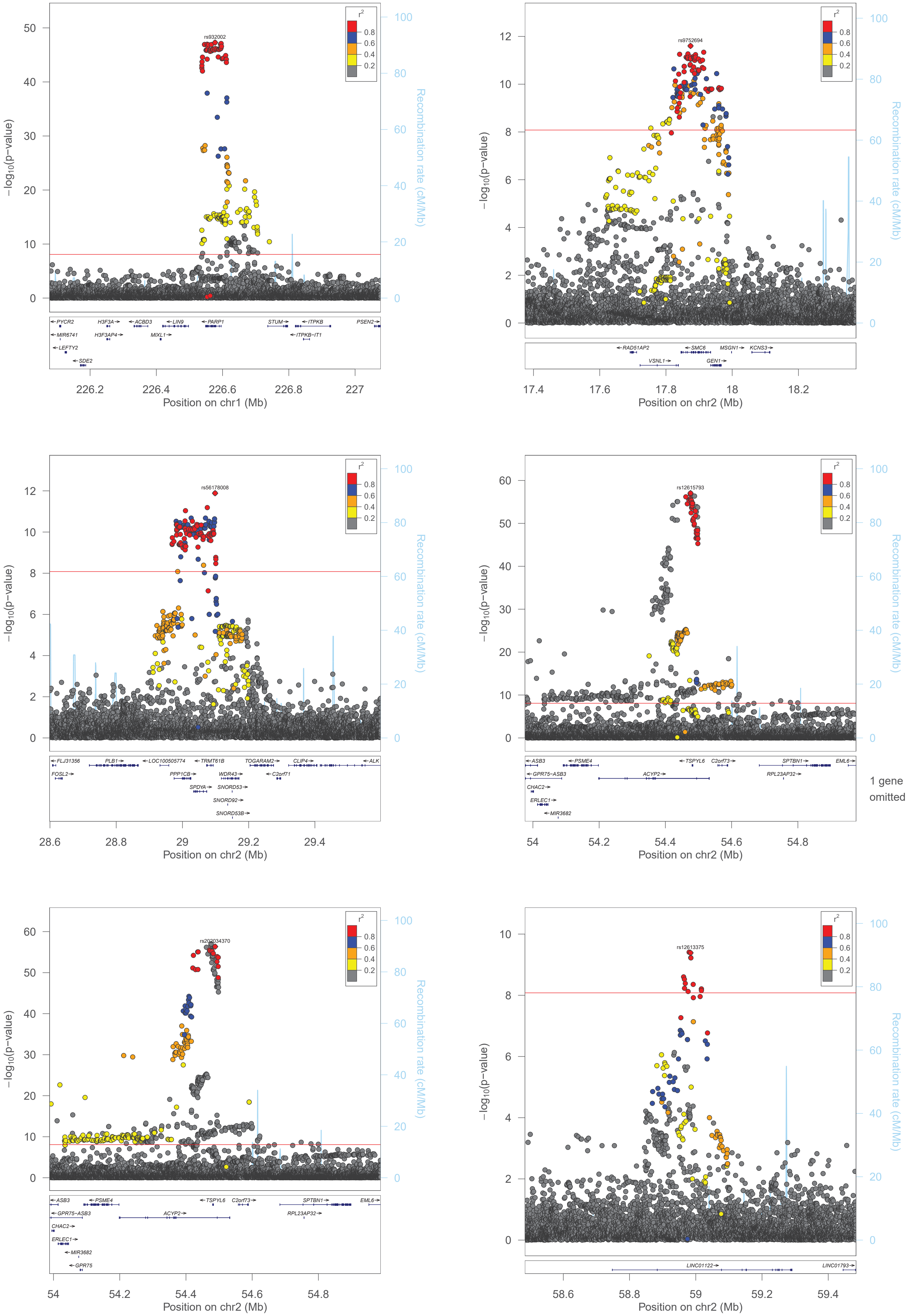
rs932002
rs9752694
rs56178008
rs12615793
rs202034370
rs12613375
r2
0.8
0.6
0.4
0.2
−log10(p−value)
Recombination rate (cM/Mb)
Position on chr1 (Mb)
Position on chr2 (Mb)
PYCR2
H3F3A
ACBD3
LIN9
PARP1
STUM
ITPKB
PSEN2
MIR6741
H3F3AP4
MIXL1
ITPKB−IT1
LEFTY2
SDE2
RAD51AP2
SMC6
MSGN1
KCNS3
VSNL1
GEN1
FLJ31356
PLB1
LOC100505774
TRMT61B
TOGARAM2
CLIP4
ALK
FOSL2
PPP1CB
WDR43
C2orf71
SPDYA
SNORD53
SNORD92
SNORD53B
ASB3
PSME4
TSPYL6
C2orf73
SPTBN1
EML6
GPR75−ASB3
ACYP2
RPL23AP32
CHAC2
ERLEC1
MIR3682
GPR75
1 gene omitted
LINC01122
LINC01793

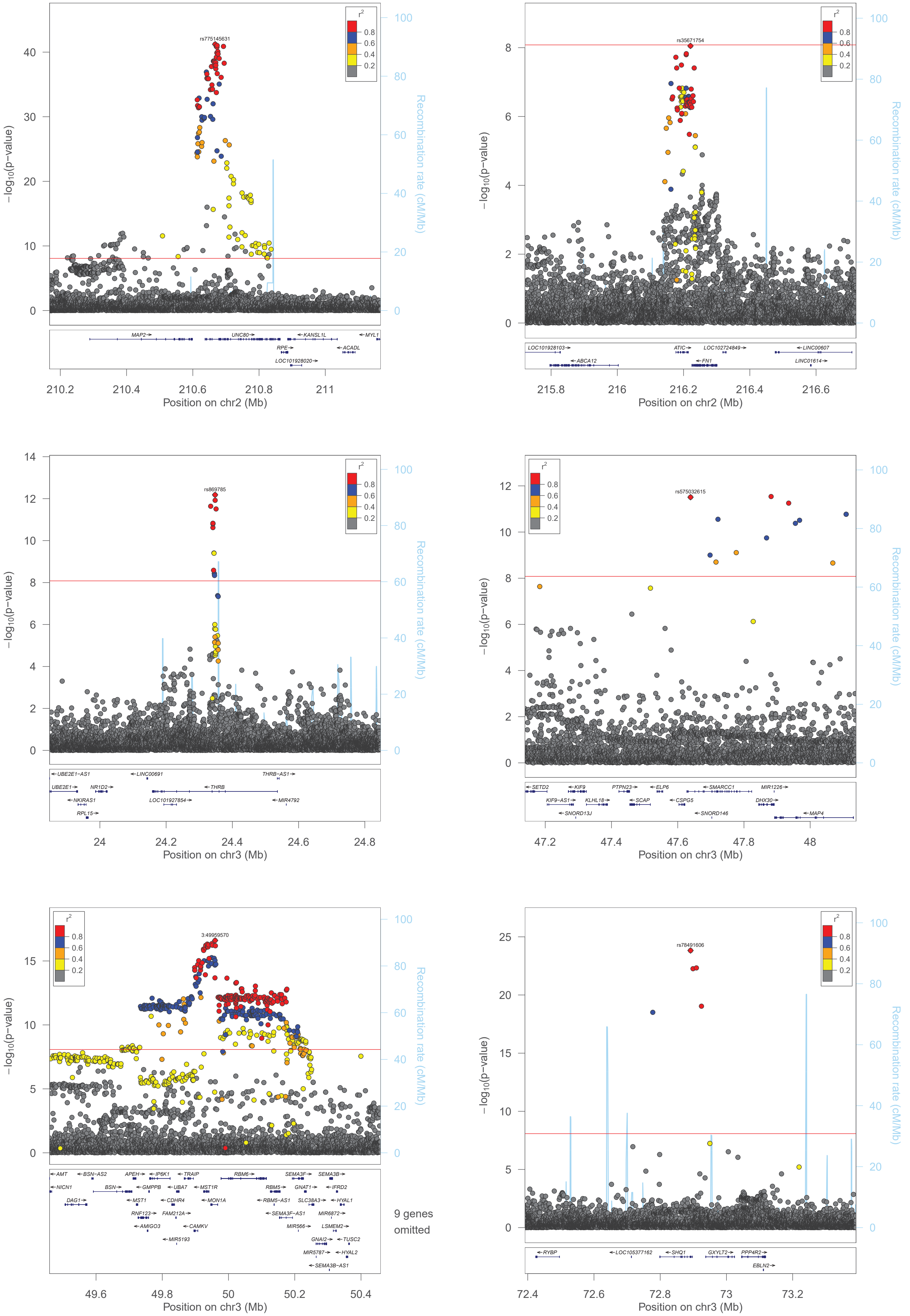

−log10(p−value)
Recombination rate (cM/Mb)
r2
0.8
0.6
0.4
0.2
rs775145631
MAP2→
UNC80→
←KANSL1L
←MYL1
RPE→
←ACADL
LOC101928020→
Position on chr2 (Mb)
rs35671754
LOC101928103→
←ABCA12
ATIC→
←FN1
LOC102724849→
←LINC00607
LINC01614→
Position on chr2 (Mb)
rs869785
←UBE2E1−AS1
←LINC00691
THRB−AS1→
UBE2E1→
NR1D2→
←THRB
←NKIRAS1
LOC101927854→
←MIR4792
RPL15→
Position on chr3 (Mb)
rs575032615
←SETD2
←KIF9
PTPN23→
←ELP6
←SMARCC1
MIR1226→
KIF9−AS1→
KLHL18→
←SCAP
←CSPG5
DHX30→
←SNORD13J
←SNORD146
←MAP4
Position on chr3 (Mb)
3:49959570
←AMT
←BSN−AS2
APEH→
←IP6K1
←TRAIP
RBM6→
SEMA3F→
SEMA3B→
←NICN1
BSN→
←GMPPB
←UBA7
←MST1R
RBM5→
GNAT1→
←IFRD2
DAG1→
←MST1
←CDHR4
←MON1A
←RBM5−AS1
SLC38A3→
←HYAL1
RNF123→
FAM212A→
←SEMA3F−AS1
MIR6872→
←AMIGO3
←CAMKV
MIR566→
LSMEM2→
←MIR5193
GNAI2→
←TUSC2
MIR5787→
←HYAL2
←SEMA3B−AS1
9 genes omitted
Position on chr3 (Mb)
rs78491606
←RYBP
←LOC105377162
←SHQ1
GXYLT2→
PPP4R2→
EBLN2→
Position on chr3 (Mb)

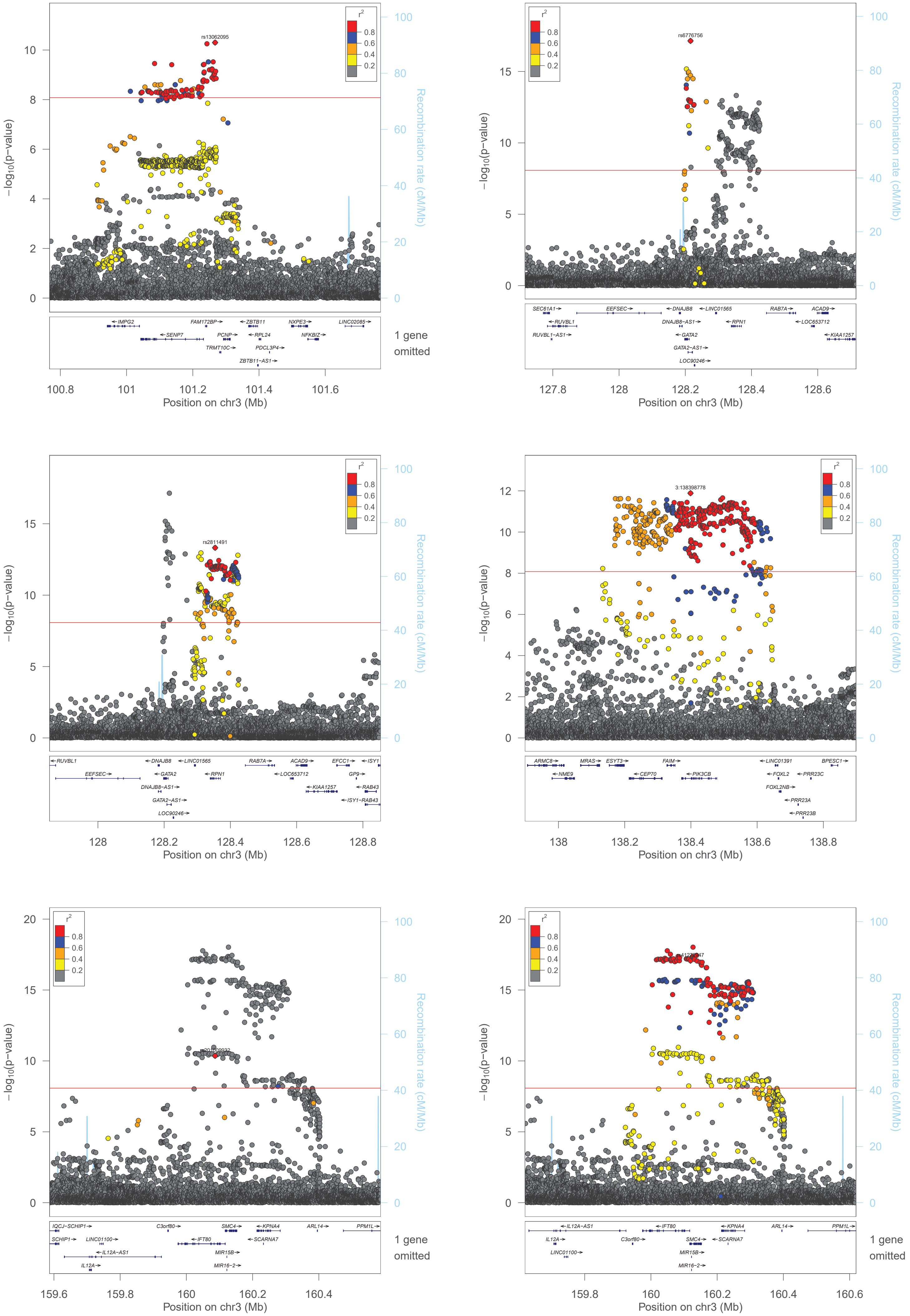
rs13062095
rs6776756
rs2811491
3:138398778
−log10(p−value)
Recombination rate (cM/Mb)
r2
0.8
0.6
0.4
0.2
Position on chr3 (Mb)
1 gene omitted
IMPG2
FAM172BP
ZBTB11
NXPE3
LINC02085
SENP7
PCNP
RPL24
NFKBIZ
TRMT10C
PDCL3P4
ZBTB11-AS1
SEC61A1
EEFSEC
DNAJB8
LINC01565
RAB7A
ACAD9
RUVBL1
DNAJB8-AS1
RPN1
LOC653712
RUVBL1-AS1
GATA2
KIAA1257
GATA2-AS1
LOC90246
EFCC1
ISY1
GP9
RAB43
ISY1-RAB43
ARMC8
MRAS
ESYT3
FAIM
LINC01391
BPESC1
NME9
CEP70
PIK3CB
FOXL2
PRR23C
FOXL2NB
PRR23A
PRR23B
IQCJ-SCHIP1
C3orf80
SMC4
KPNA4
ARL14
PPM1L
SCHIP1
LINC01100
IFT80
SCARNA7
IL12A-AS1
MIR15B
IL12A
MIR16-2

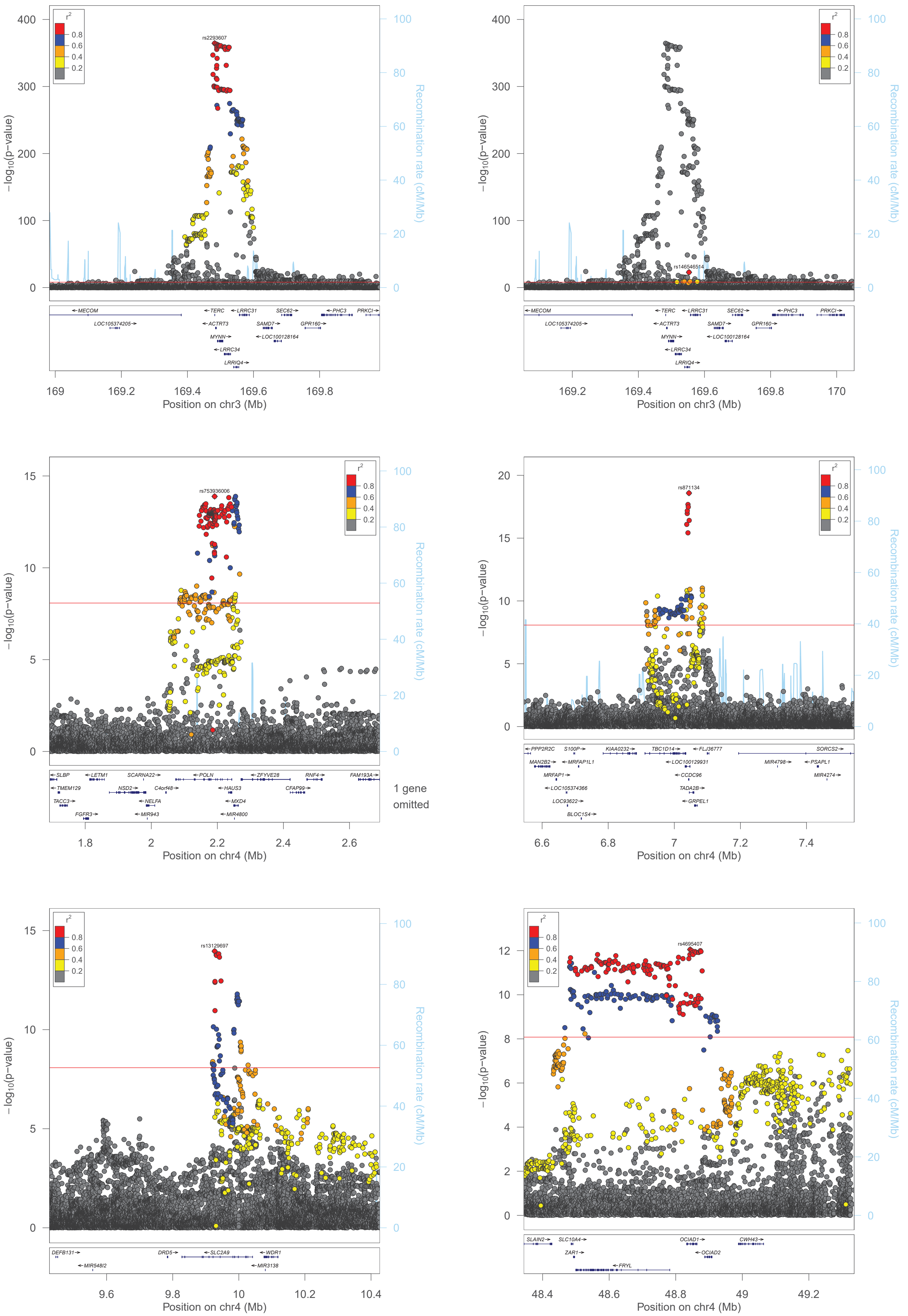

r2
0.8
0.6
0.4
0.2
rs2293607
rs146546514
rs753936006
rs871134
rs13129697
rs4695407
−log10(p−value)
Recombination rate (cM/Mb)
Position on chr3 (Mb)
Position on chr4 (Mb)
MECOM
LOC105374205
TERC
ACTRT3
MYNN
LRRC34
LRRIQ4
LRRC31
SEC62
SAMD7
LOC100128164
GPR160
PHC3
PRKCI
SLBP
TMEM129
TACC3
FGFR3
LETM1
NSD2
SCARNA22
C4orf48
NELFA
MIR943
POLN
HAUS3
MXD4
MIR4800
ZFYVE28
CFAP99
RNF4
FAM193A
1 gene omitted
PPP2R2C
MAN2B2
MRFAP1
LOC105374366
LOC93622
BLOC1S4
S100P
MRFAP1L1
KIAA0232
TBC1D14
LOC100129931
CCDC96
TADA2B
GRPEL1
FLJ36777
MIR4798
SORCS2
PSAPL1
MIR4274
DEFB131
MIR548I2
DRD5
SLC2A9
WDR1
MIR3138
SLAIN2
SLC10A4
ZAR1
FRYL
OCIAD1
OCIAD2
CWH43

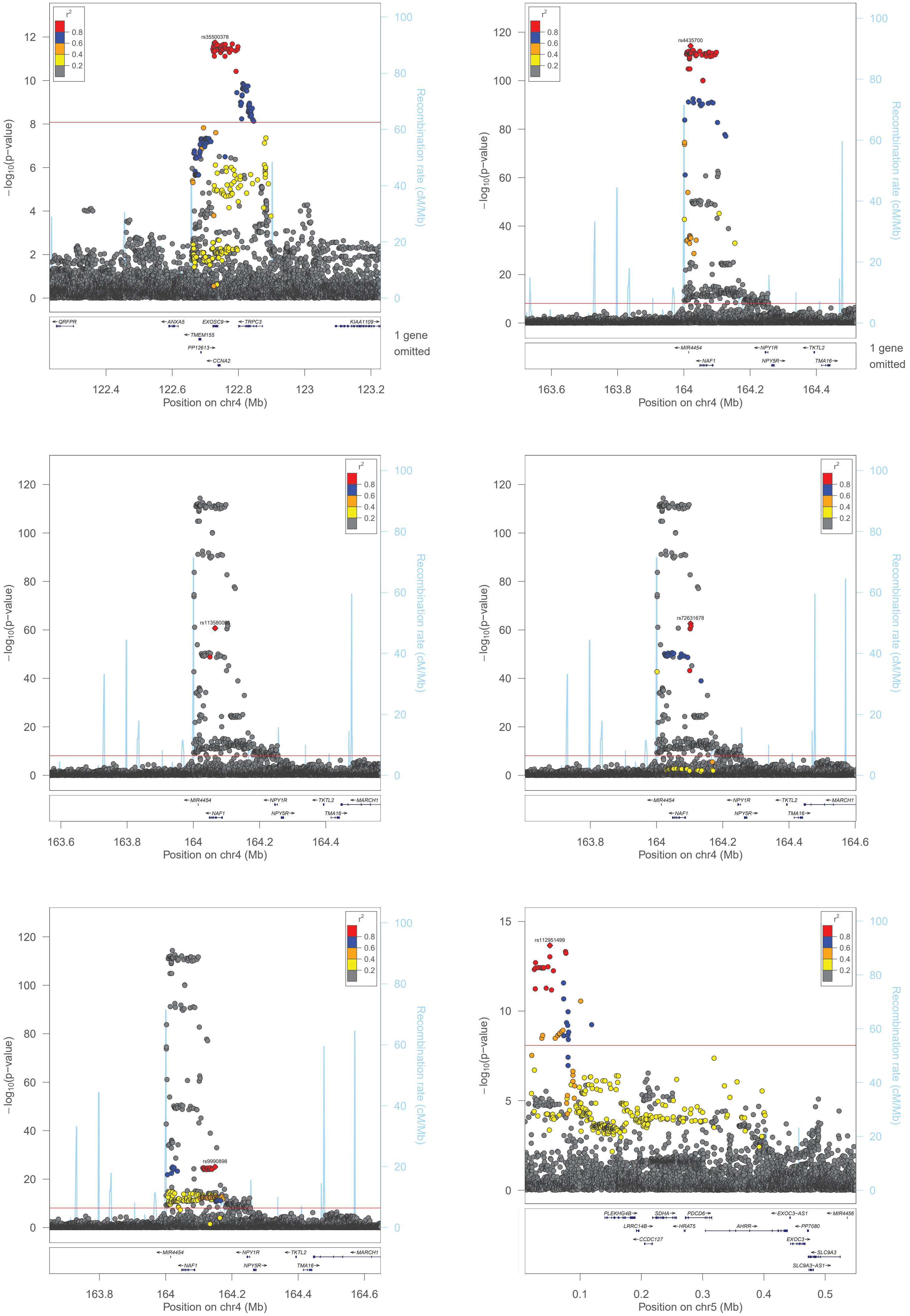

r2
0.8
0.6
0.4
0.2
rs35500378
−log10(p−value)
Recombination rate (cM/Mb)
QRFPR
ANXA5
EXOSC9
TRPC3
KIAA1109
TMEM155
PP12613
CCNA2
1 gene omitted
Position on chr4 (Mb)
rs4435700
MIR4454
NAF1
NPY1R
NPY5R
TKTL2
TMA16
1 gene omitted
Position on chr4 (Mb)
rs113580085
MIR4454
NAF1
NPY1R
NPY5R
TKTL2
TMA16
MARCH1
Position on chr4 (Mb)
rs72631678
Position on chr4 (Mb)
rs9990898
Position on chr4 (Mb)
rs112951499
PLEKHG4B
SDHA
PDCD6
EXOC3−AS1
MIR4456
LRRC14B
HRAT5
AHRR
PP7080
CCDC127
EXOC3
SLC9A3
SLC9A3−AS1
Position on chr5 (Mb)

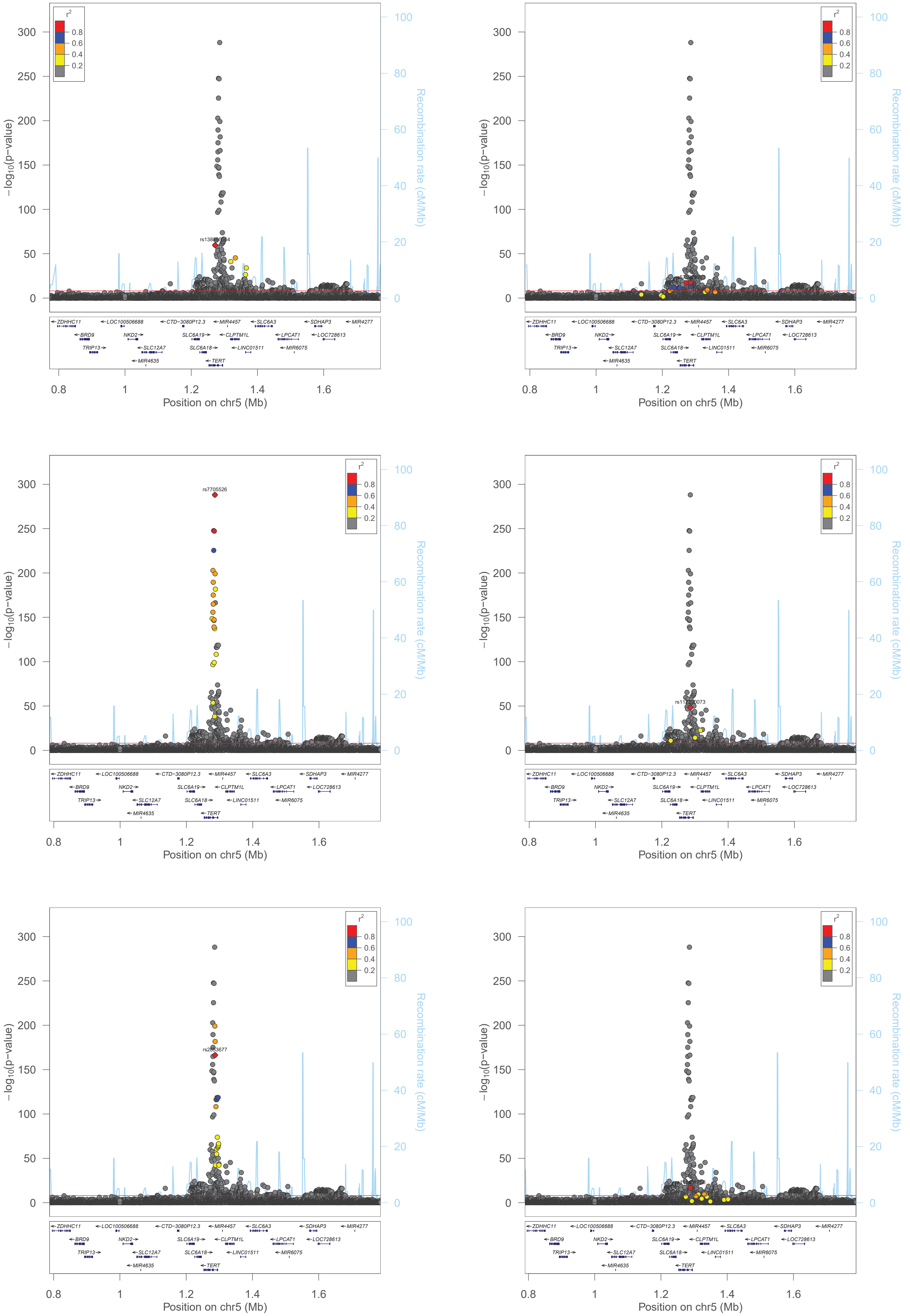

r²
0.8
0.6
0.4
0.2
−log10(p−value)
Recombination rate (cM/Mb)
Position on chr5 (Mb)
rs7705526
ZDHHC11
LOC100506688
CTD−3080P12.3
MIR4457
SLC6A3
SDHAP3
MIR4277
BRD9
NKD2
SLC6A19
CLPTM1L
LPCAT1
LOC728613
TRIP13
SLC12A7
SLC6A18
LINC01511
MIR6075
MIR4635
TERT

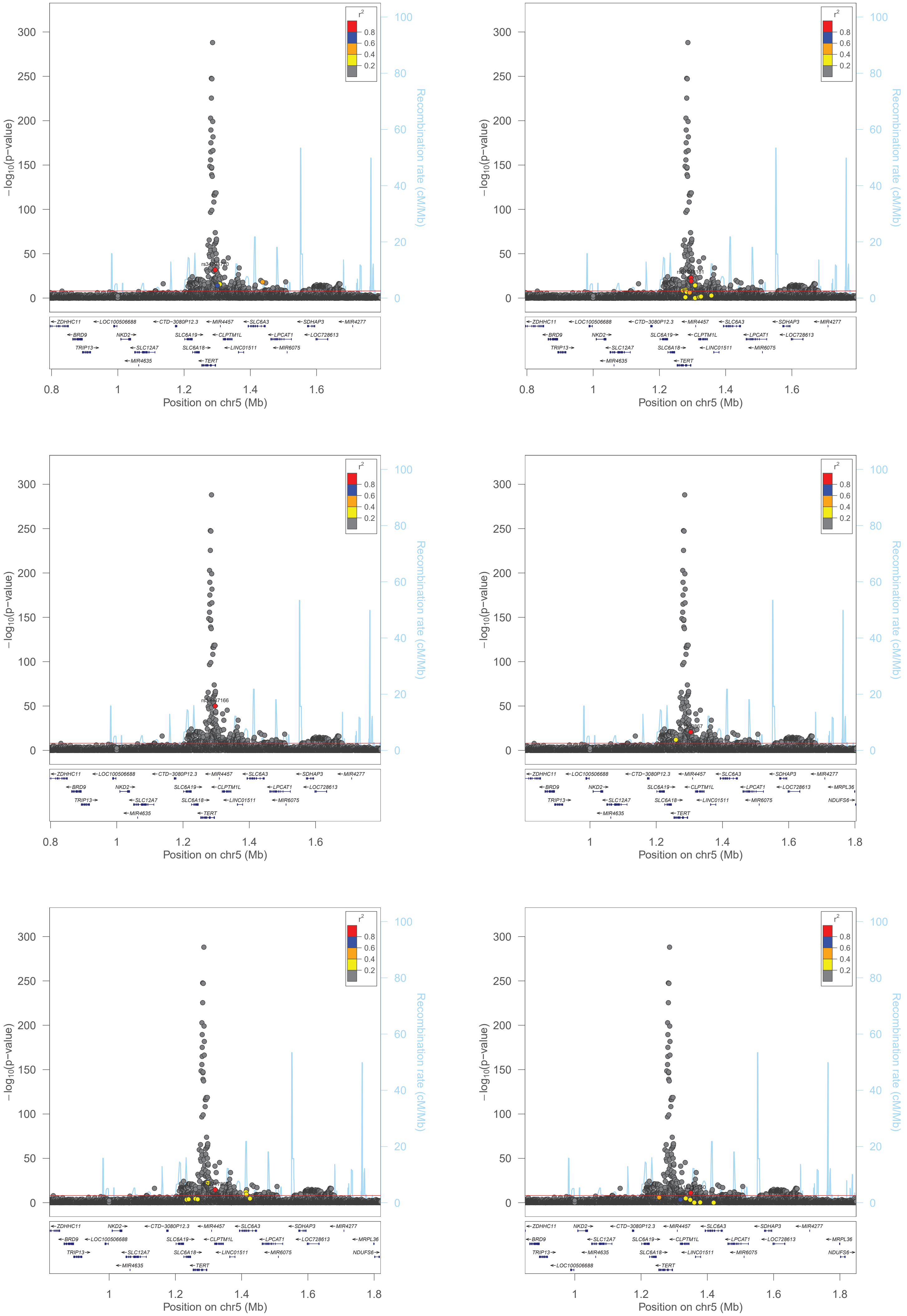

−log10(p−value)
Recombination rate (cM/Mb)
Position on chr5 (Mb)
r2
0.8
0.6
0.4
0.2
ZDHHC11
LOC100506688
CTD−3080P12.3
MIR4457
SLC6A3
SDHAP3
MIR4277
BRD9
NKD2
SLC6A19
CLPTM1L
LPCAT1
LOC728613
TRIP13
SLC12A7
SLC6A18
LINC01511
MIR6075
MIR4635
TERT
MRPL36
NDUFS6

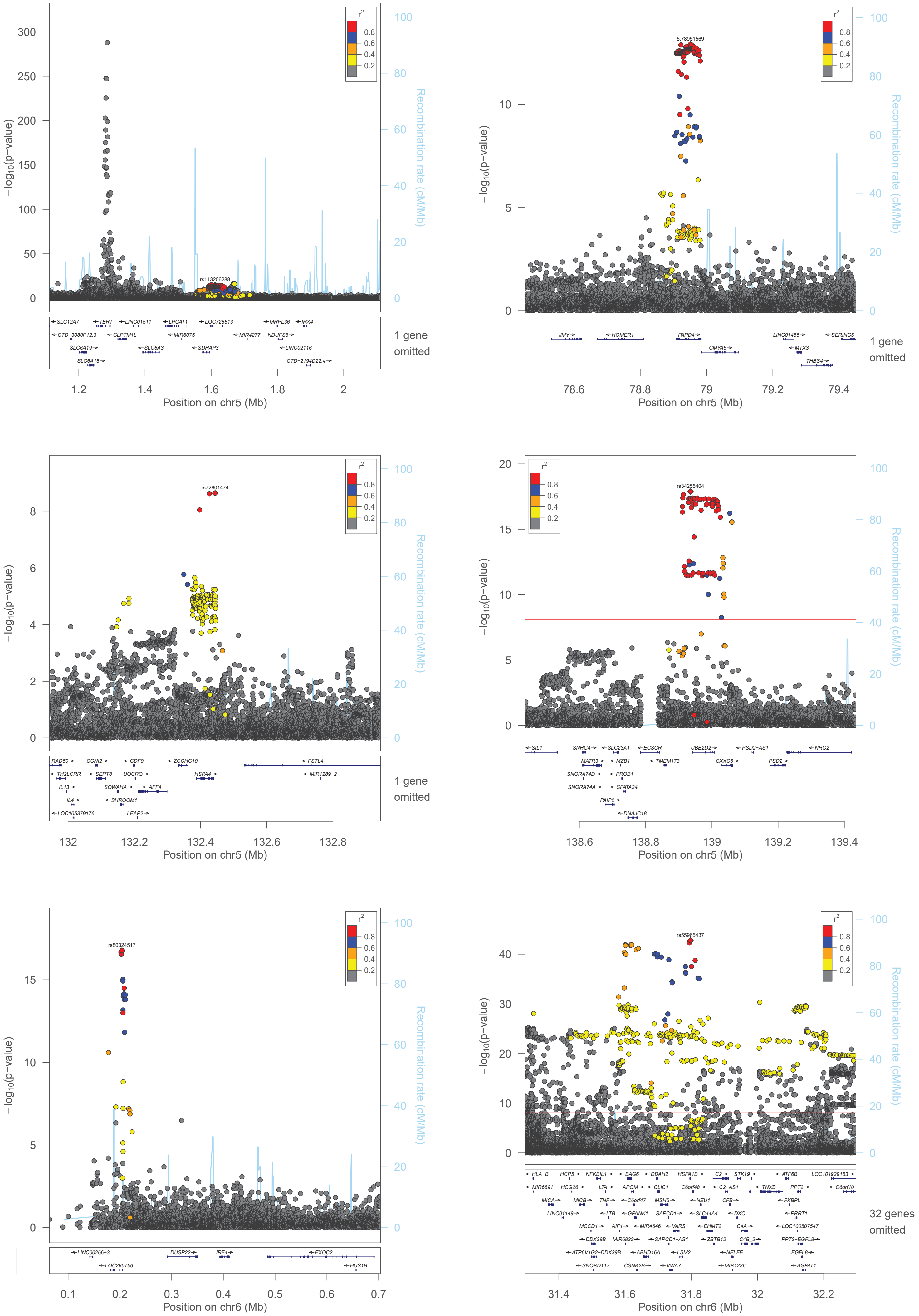

−log10(p−value)
Recombination rate (cM/Mb)
r2
rs113206288
1 gene omitted
Position on chr5 (Mb)
5:78951569
1 gene omitted
Position on chr5 (Mb)
rs72801474
1 gene omitted
Position on chr5 (Mb)
rs34255404
Position on chr5 (Mb)
rs80324517
Position on chr6 (Mb)
rs55965437
32 genes omitted
Position on chr6 (Mb)

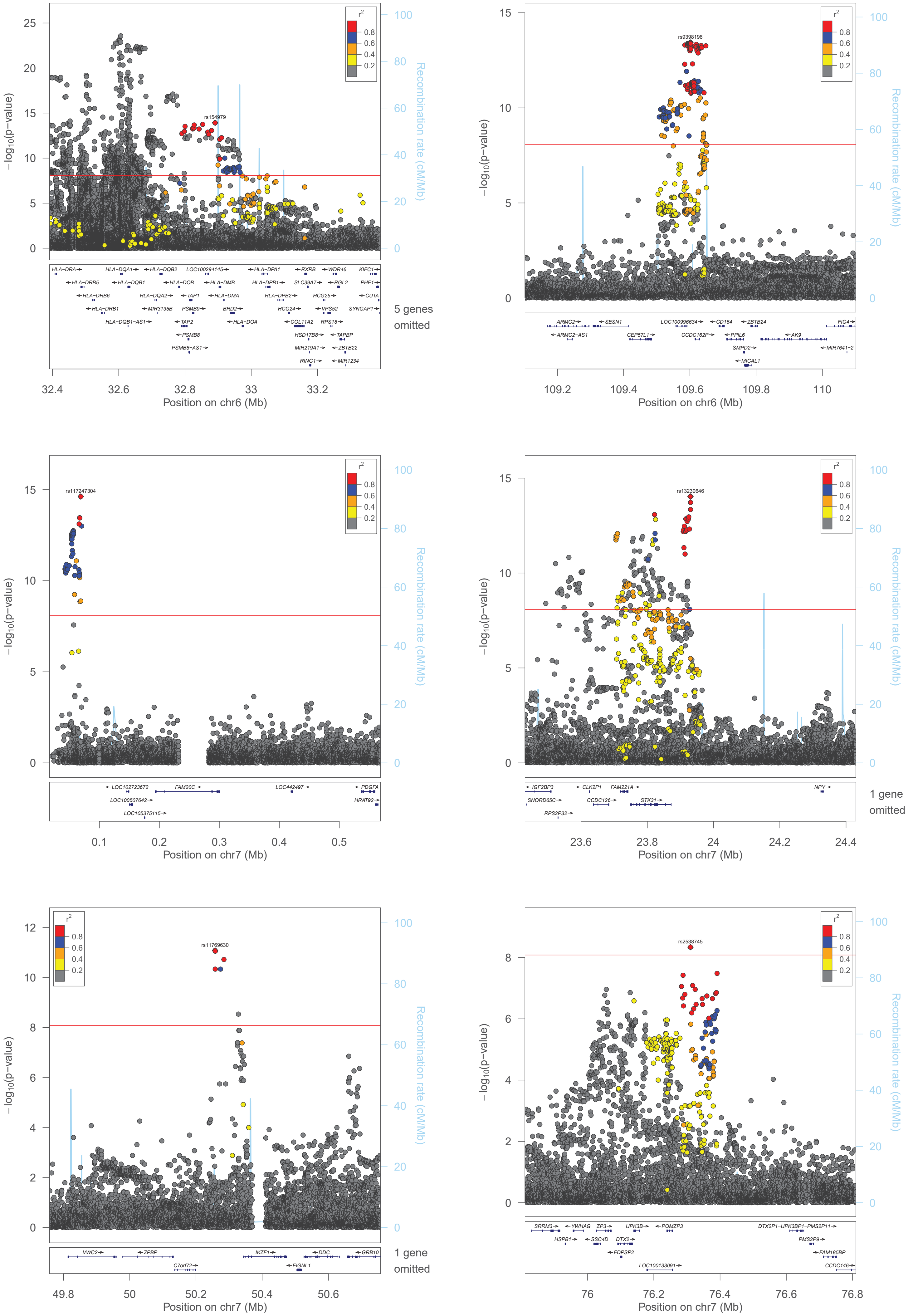

−log10(p−value)
Recombination rate (cM/Mb)
r2
0.8
0.6
0.4
0.2
rs154979
5 genes omitted
Position on chr6 (Mb)
rs9398196
Position on chr6 (Mb)
rs117247304
Position on chr7 (Mb)
rs13230646
1 gene omitted
Position on chr7 (Mb)
rs11769630
1 gene omitted
Position on chr7 (Mb)
rs2538745
Position on chr7 (Mb)

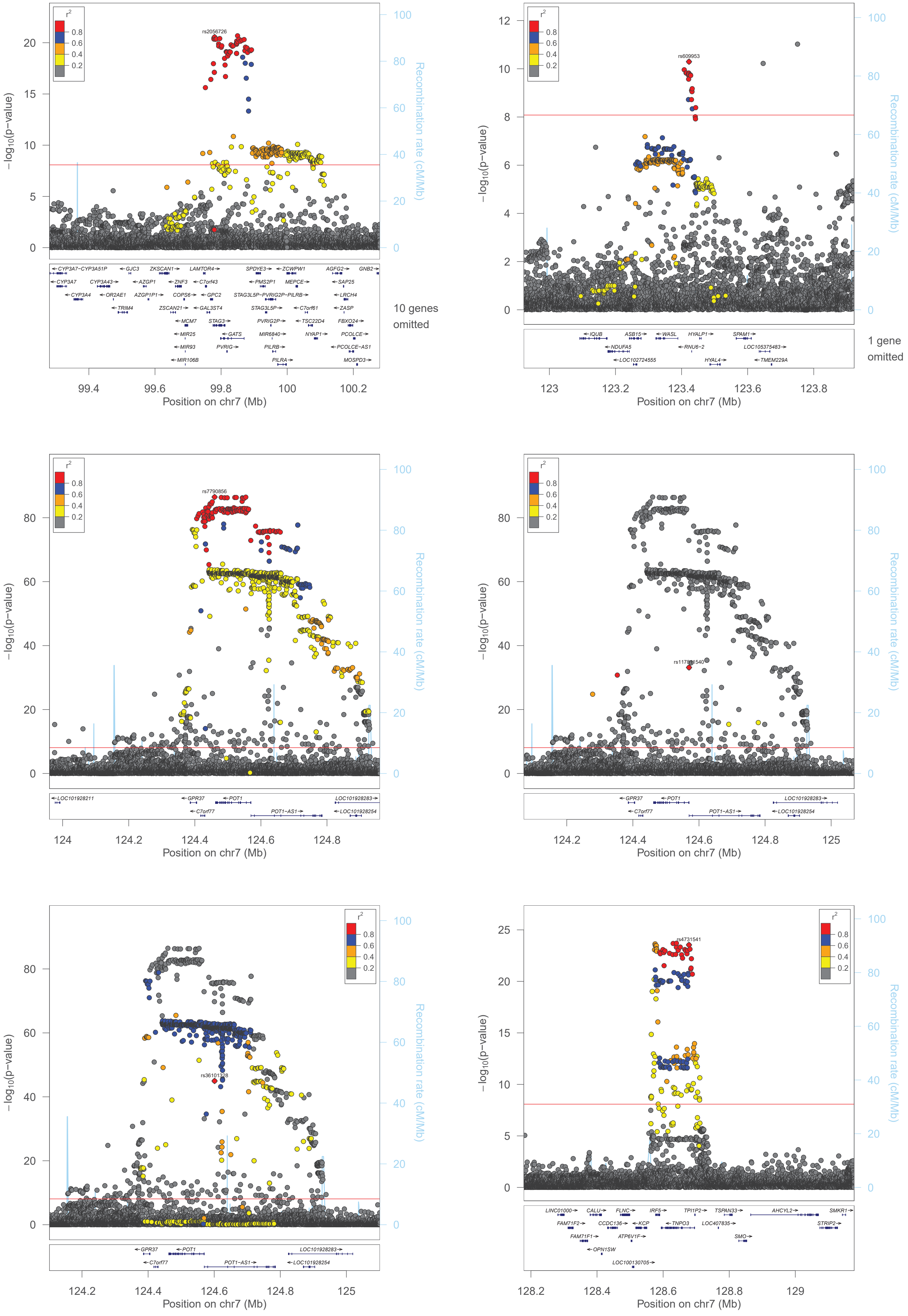

r²
0.8
0.6
0.4
0.2
rs2056726
−log10(p−value)
Recombination rate (cM/Mb)
10 genes omitted
Position on chr7 (Mb)
rs609953
1 gene omitted
rs7790856
rs117811540
rs36101328
rs4731541
CYP3A7−CYP3A51P
GJC3
ZKSCAN1
LAMTOR4
SPDYE3
ZCWPW1
AGFG2
GNB2
CYP3A7
CYP3A43
AZGP1
ZNF3
C7orf43
PMS2P1
MEPCE
SAP25
CYP3A4
OR2AE1
AZGP1P1
COPS6
GPC2
STAG3L5P−PVRIG2P−PILRB
LRCH4
TRIM4
ZSCAN21
GAL3ST4
STAG3L5P
C7orf61
ZASP
MCM7
STAG3
PVRIG2P
TSC22D4
FBXO24
MIR25
GATS
MIR6840
NYAP1
PCOLCE
MIR93
PVRIG
PILRB
PCOLCE−AS1
MIR106B
PILRA
MOSPD3
IQUB
ASB15
WASL
HYALP1
SPAM1
NDUFA5
RNU6−2
LOC105375483
LOC102724555
HYAL4
TMEM229A
LOC101928211
GPR37
POT1
LOC101928283
C7orf77
POT1−AS1
LOC101928254
LINC01000
CALU
FLNC
IRF5
TPI1P2
TSPAN33
AHCYL2
SMKR1
FAM71F2
CCDC136
KCP
TNPO3
LOC407835
STRIP2
FAM71F1
ATP6V1F
SMO
OPN1SW
LOC100130705

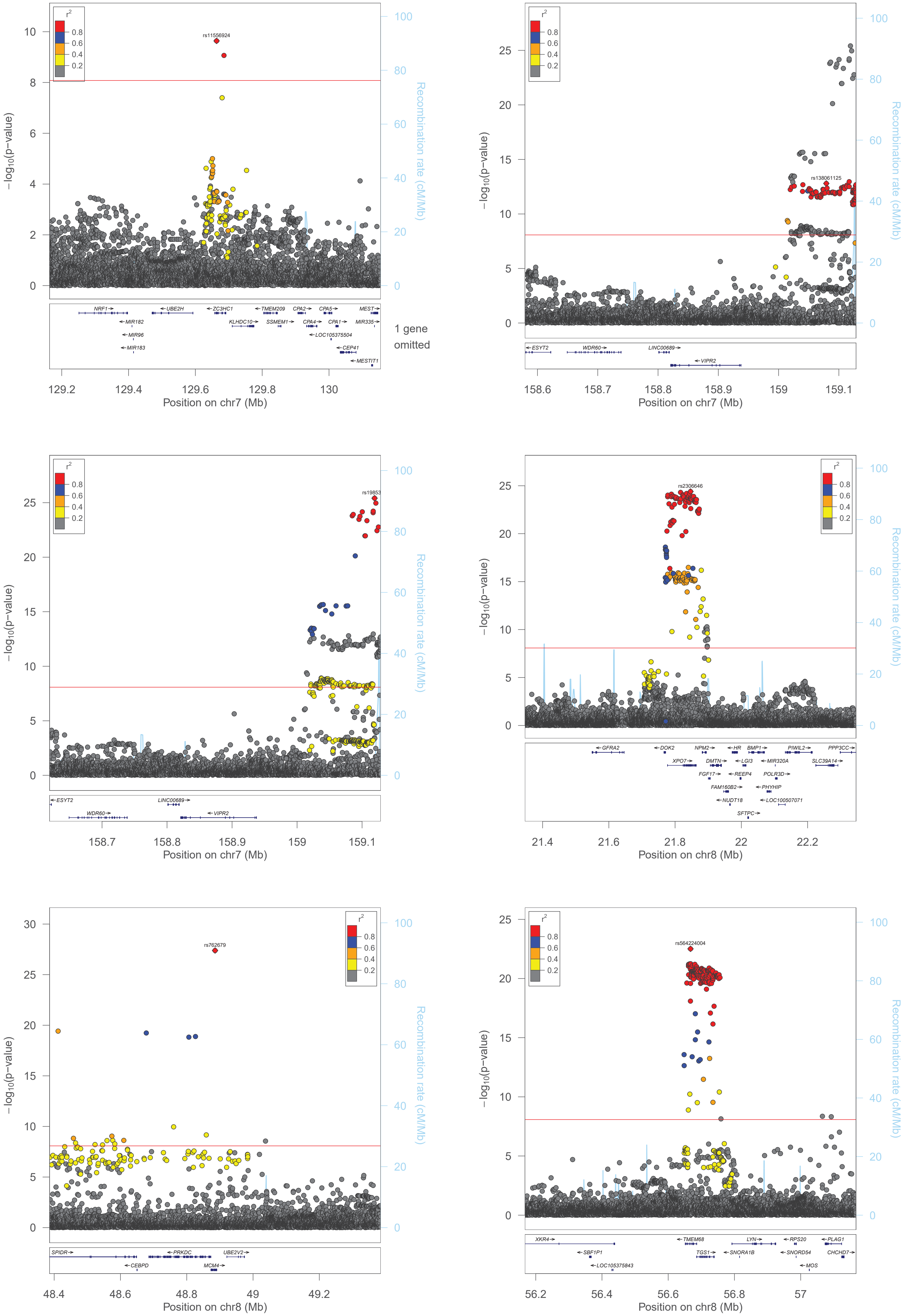

r2
0.8
0.6
0.4
0.2
rs11556924
−log10(p−value)
Recombination rate (cM/Mb)
NRF1
UBE2H
ZC3HC1
TMEM209
CPA2
CPA5
MEST
MIR182
KLHDC10
SSMEM1
CPA4
CPA1
MIR335
MIR96
LOC105375504
MIR183
CEP41
MESTIT1
1 gene omitted
Position on chr7 (Mb)
rs138061125
ESYT2
WDR60
LINC00689
VIPR2
Position on chr7 (Mb)
rs19853
ESYT2
LINC00689
WDR60
VIPR2
Position on chr7 (Mb)
rs2306646
GFRA2
DOK2
NPM2
HR
BMP1
PIWIL2
PPP3CC
XPO7
DMTN
LGI3
MIR320A
SLC39A14
FGF17
REEP4
POLR3D
FAM160B2
PHYHIP
NUDT18
LOC100507071
SFTPC
Position on chr8 (Mb)
rs762679
SPIDR
PRKDC
UBE2V2
CEBPD
MCM4
Position on chr8 (Mb)
rs564224004
XKR4
TMEM68
LYN
RPS20
PLAG1
SBF1P1
TGS1
SNORA1B
SNORD54
CHCHD7
LOC105375843
MOS
Position on chr8 (Mb)

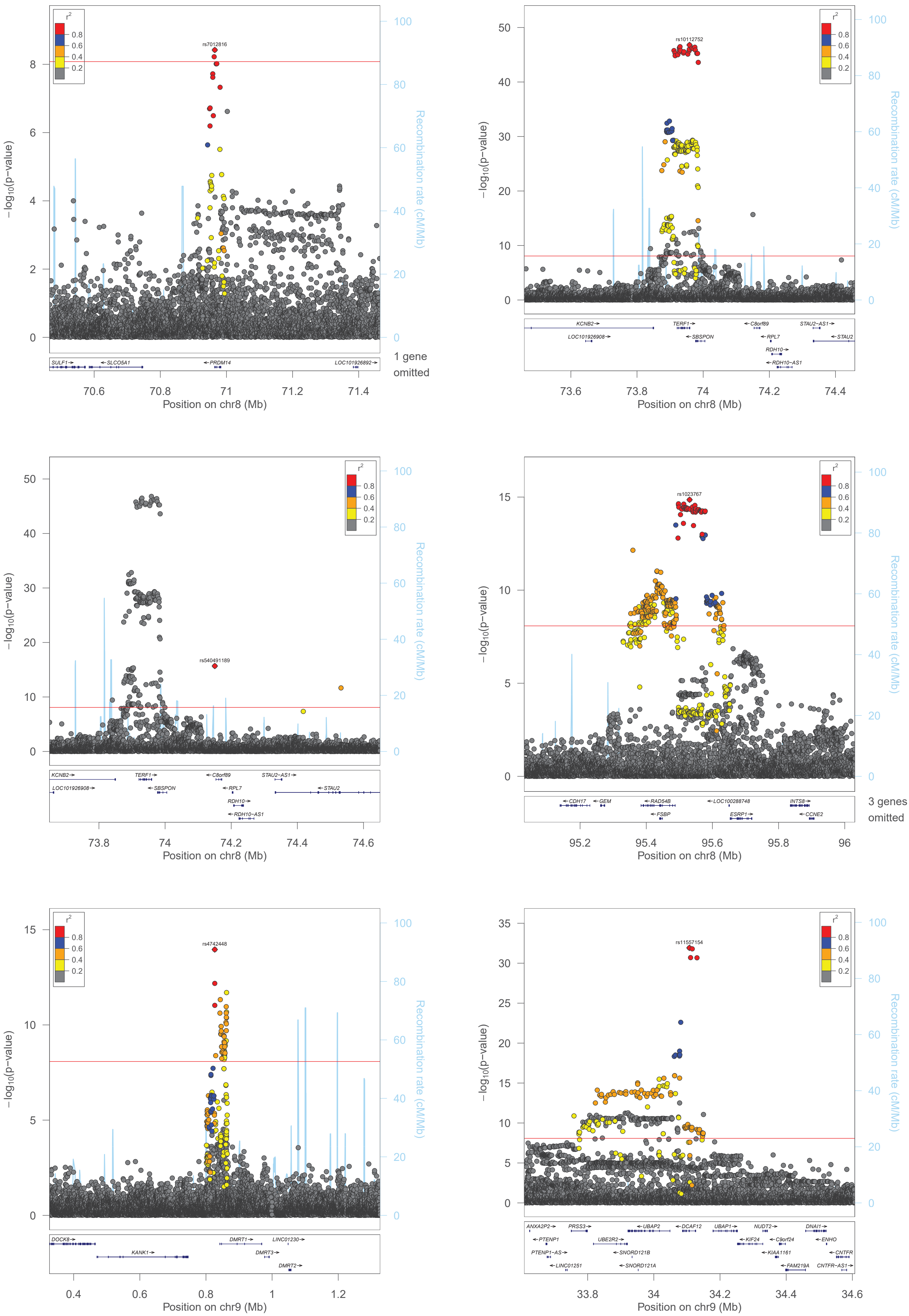

r²
0.8
0.6
0.4
0.2
−log10(p−value)
Recombination rate (cM/Mb)
rs7012816
SULF1→
←SLCO5A1
←PRDM14
LOC101926892→
1 gene omitted
Position on chr8 (Mb)
rs10112752
KCNB2→
LOC101926908→
TERF1→
←SBSPON
←C8orf89
←RPL7
RDH10→
←RDH10−AS1
STAU2−AS1→
←STAU2
rs540491189
rs1023767
←CDH17
←GEM
←RAD54B
←FSBP
←LOC100288748
ESRP1→
INTS8→
←CCNE2
3 genes omitted
rs4742448
DOCK8→
KANK1→
DMRT1→
LINC01230→
DMRT3→
DMRT2→
Position on chr9 (Mb)
rs11557154
ANXA2P2→
←PTENP1
PTENP1−AS→
←LINC01251
PRSS3→
UBE2R2→
←SNORD121B
←SNORD121A
←UBAP2
←DCAF12
UBAP1→
←KIF24
NUDT2→
←C9orf24
←KIAA1161
←FAM219A
DNAI1→
←ENHO
←CNTFR
CNTFR−AS1→

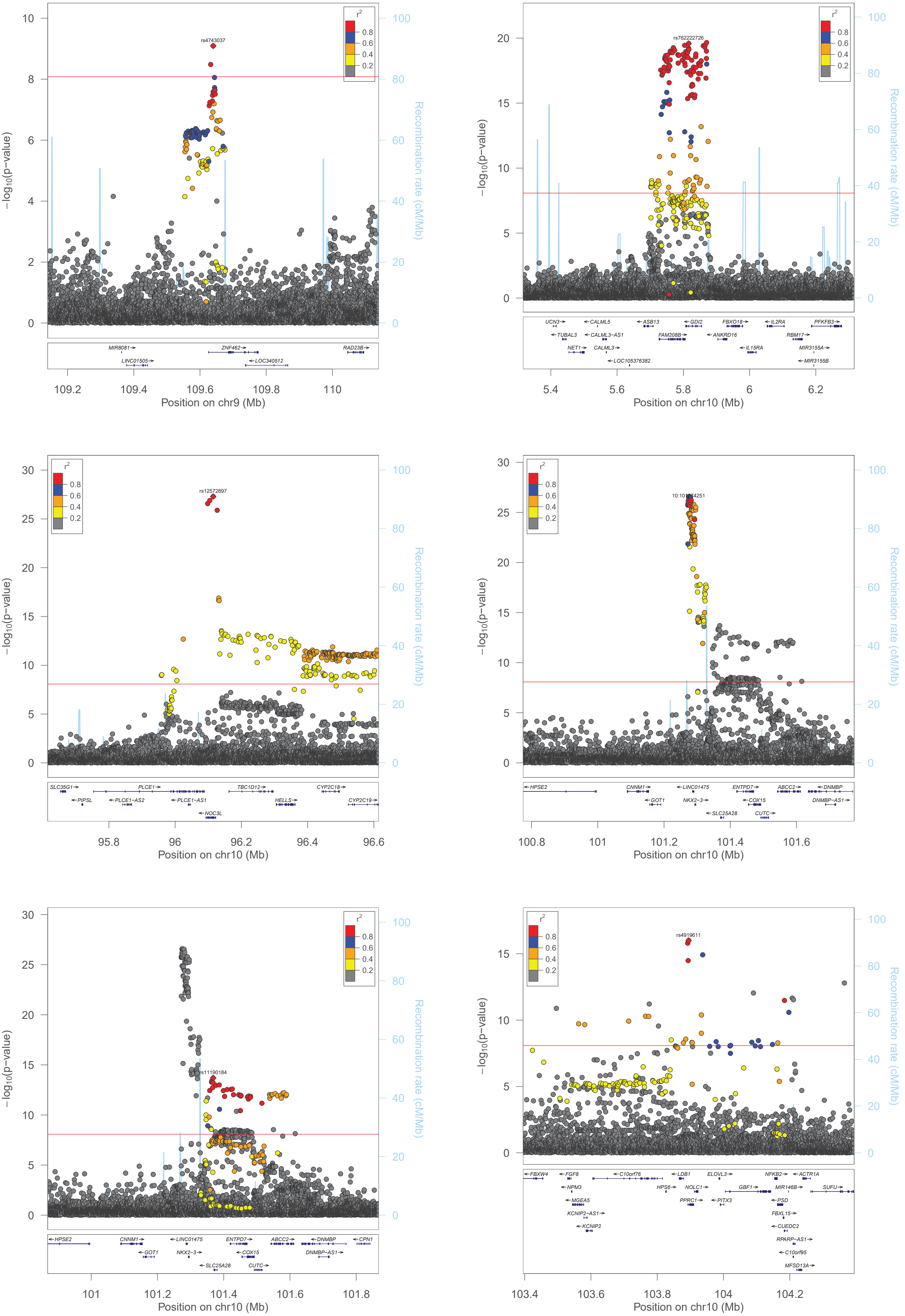

−log10(p−value)
Recombination rate (cM/Mb)
r2
0.8
0.6
0.4
0.2
rs4743037
MIR8081
LINC01505
ZNF462
LOC340512
RAD23B
Position on chr9 (Mb)
rs762222726
UCN3
TUBAL3
NET1
CALML5
CALML3−AS1
CALML3
LOC105376382
ASB13
FAM208B
GDI2
ANKRD16
FBXO18
IL15RA
IL2RA
RBM17
PFKFB3
MIR3155A
MIR3155B
Position on chr10 (Mb)
rs12572897
SLC35G1
PIPSL
PLCE1
PLCE1−AS2
PLCE1−AS1
NOC3L
TBC1D12
HELLS
CYP2C18
CYP2C19
Position on chr10 (Mb)
HPSE2
CNNM1
GOT1
LINC01475
NKX2−3
SLC25A28
ENTPD7
COX15
CUTC
ABCC2
DNMBP
DNMBP−AS1
Position on chr10 (Mb)
rs11190184
HPSE2
CNNM1
GOT1
LINC01475
NKX2−3
SLC25A28
ENTPD7
COX15
CUTC
ABCC2
DNMBP
DNMBP−AS1
CPN1
Position on chr10 (Mb)
rs4919611
FBXW4
FGF8
NPM3
MGEA5
KCNIP2−AS1
KCNIP2
C10orf76
HPS6
LDB1
NOLC1
PPRC1
ELOVL3
PITX3
GBF1
NFKB2
PSD
FBXL15
CUEDC2
RPARP−AS1
C10orf95
MFSD13A
MIR146B
ACTR1A
SUFU
Position on chr10 (Mb)

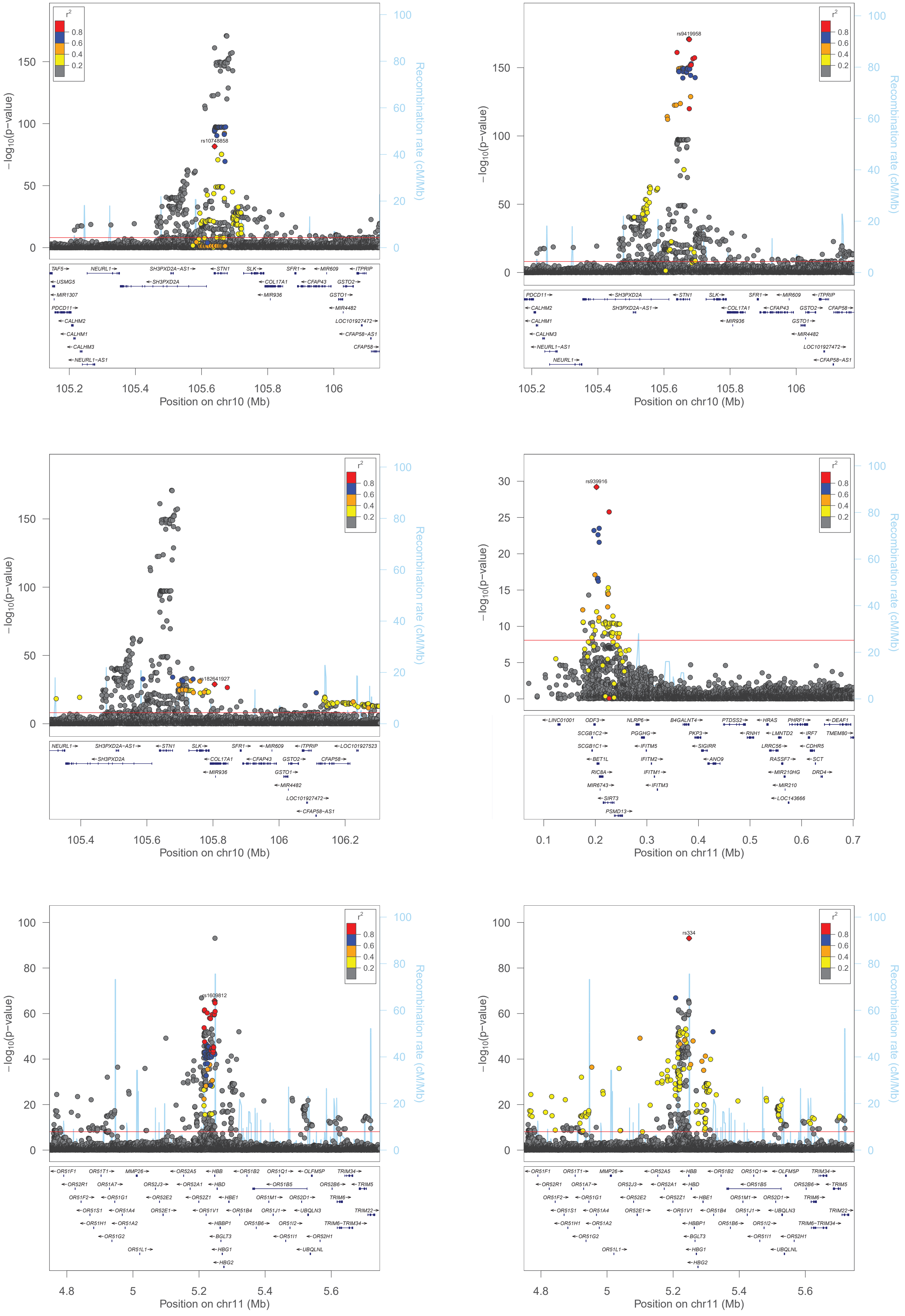

−log10(p−value)
Recombination rate (cM/Mb)
r2
0.8
0.6
0.4
0.2
rs10748858
Position on chr10 (Mb)
TAF5 NEURL1 SH3PXD2A−AS1 STN1 SLK SFR1 MIR609 ITPRIP
USMG5 SH3PXD2A COL17A1 CFAP43 GSTO2
MIR1307 MIR936 GSTO1
PDCD11 MIR4482
CALHM2 LOC101927472
CALHM1 CFAP58−AS1
CALHM3 CFAP58
NEURL1−AS1
rs9419958
Position on chr10 (Mb)
PDCD11 SH3PXD2A STN1 SLK SFR1 MIR609 ITPRIP
CALHM2 SH3PXD2A−AS1 COL17A1 CFAP43 GSTO2 CFAP58
CALHM1 MIR936 GSTO1
CALHM3 MIR4482
NEURL1−AS1 LOC101927472
NEURL1 CFAP58−AS1
rs182641927
Position on chr10 (Mb)
NEURL1 SH3PXD2A−AS1 STN1 SLK SFR1 MIR609 ITPRIP LOC101927523
SH3PXD2A COL17A1 CFAP43 GSTO2 CFAP58
MIR936 GSTO1
MIR4482
LOC101927472
CFAP58−AS1
rs939916
Position on chr11 (Mb)
LINC01001 ODF3 NLRP6 B4GALNT4 PTDSS2 HRAS PHRF1 DEAF1
SCGB1C2 PGGHG PKP3 RNH1 LMNTD2 IRF7 TMEM80
SCGB1C1 IFITM5 SIGIRR LRRC56 CDHR5
BET1L IFITM2 ANO9 RASSF7 SCT
RIC8A IFITM1 MIR210HG DRD4
MIR6743 IFITM3 MIR210
SIRT3 LOC143666
PSMD13
rs1609812
Position on chr11 (Mb)
rs334
Position on chr11 (Mb)
OR51F1 OR51T1 MMP26 OR52A5 HBB OR51B2 OR51Q1 OLFM5P TRIM34
OR52R1 OR51A7 OR52J3 OR52A1 HBD OR51B5 OR52B6 TRIM5
OR51F2 OR51G1 OR52E2 OR52Z1 HBE1 OR51M1 OR52D1 TRIM6
OR51S1 OR51A4 OR52E1 OR51V1 OR51B4 OR51J1 UBQLN3 TRIM22
OR51H1 OR51A2 HBBP1 OR51B6 OR51I2 TRIM6−TRIM34
OR51G2 BGLT3 OR51I1 OR52H1
OR51L1 HBG1 UBQLNL
HBG2

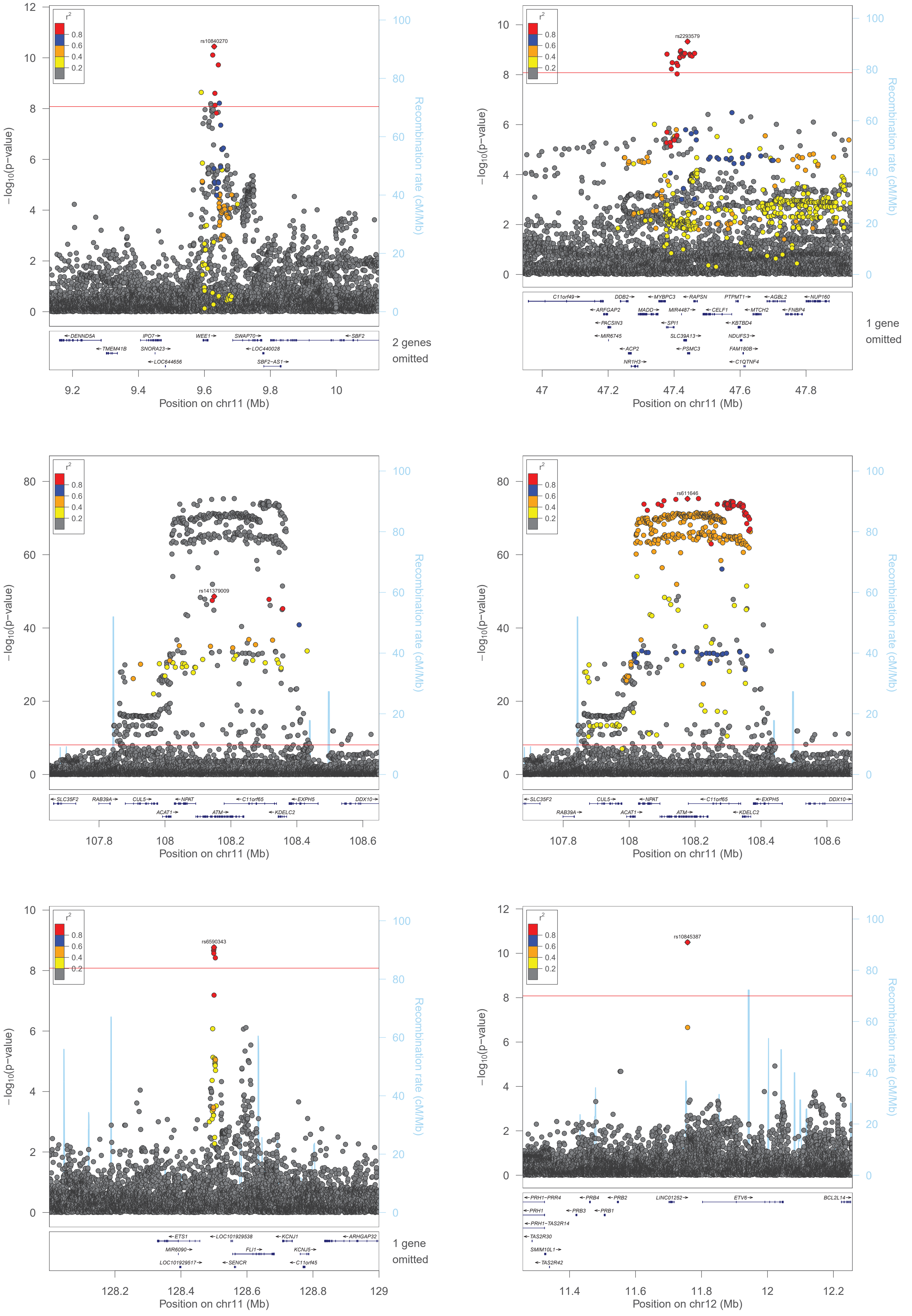

rs10840270
rs2293579
rs141379009
rs611646
rs6590343
rs10845387
r2
0.8
0.6
0.4
0.2
−log10(p−value)
Recombination rate (cM/Mb)
Position on chr11 (Mb)
Position on chr12 (Mb)
2 genes omitted
1 gene omitted
1 gene omitted

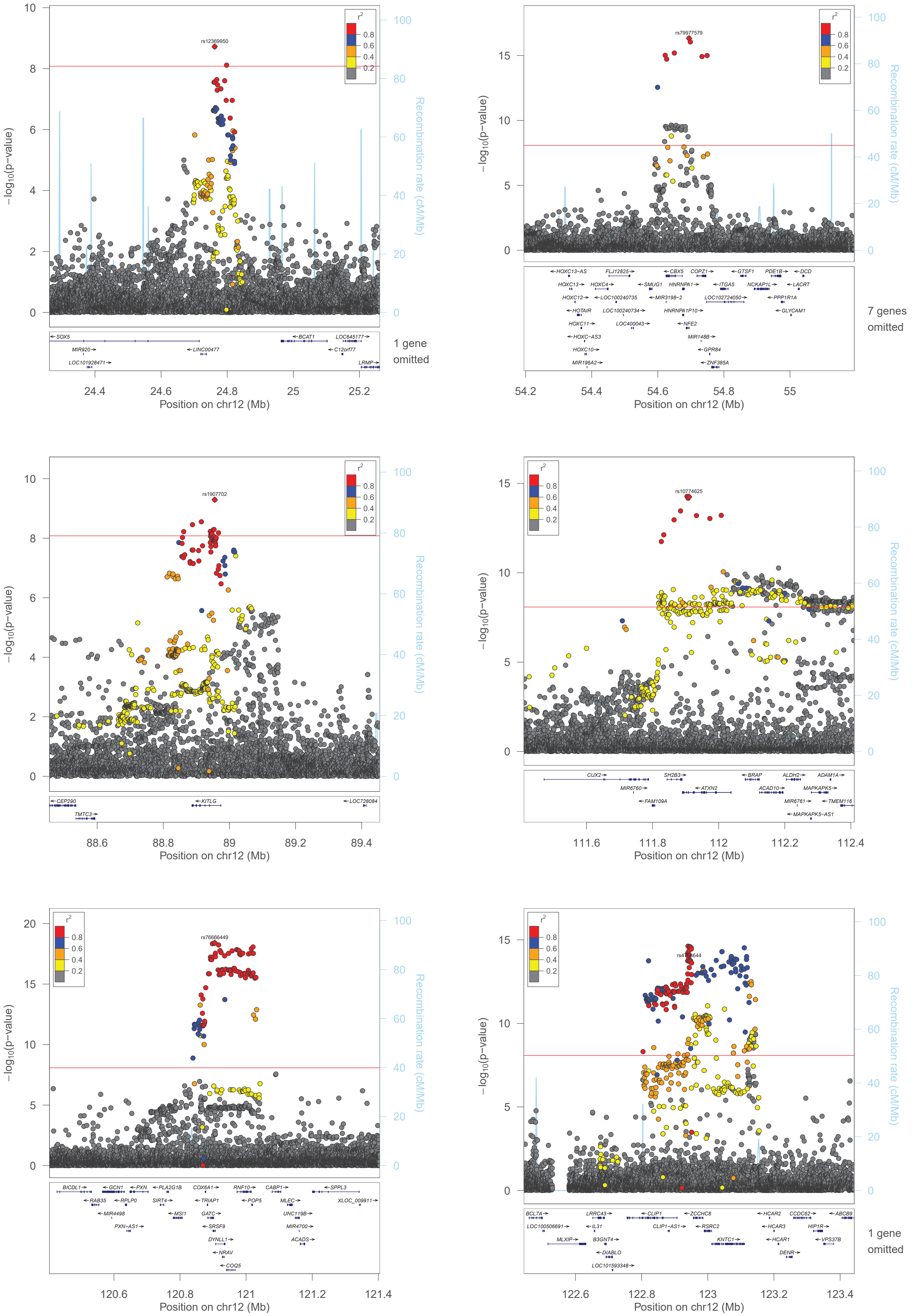

−log10(p−value)
Recombination rate (cM/Mb)
r2
0.8
0.6
0.4
0.2
rs12369950
SOX5
BCAT1
LOC645177
MIR920
LINC00477
C12orf77
LOC101928471
LRMP
1 gene omitted
Position on chr12 (Mb)
rs79977579
HOXC13−AS
FLJ12825
CBX5
COPZ1
GTSF1
PDE1B
DCD
HOXC13
HOXC4
SMUG1
HNRNPA1
ITGA5
NCKAP1L
LACRT
HOXC12
LOC100240735
MIR3198−2
LOC102724050
PPP1R1A
HOTAIR
LOC100240734
HNRNPA1P10
GLYCAM1
HOXC11
LOC400043
NFE2
HOXC−AS3
MIR148B
HOXC10
GPR84
MIR196A2
ZNF385A
7 genes omitted
rs1907702
CEP290
KITLG
LOC728084
TMTC3
rs10774625
CUX2
SH2B3
BRAP
ALDH2
ADAM1A
MIR6760
ATXN2
ACAD10
MAPKAPK5
FAM109A
MIR6761
TMEM116
MAPKAPK5−AS1
rs76666449
BICDL1
GCN1
PXN
PLA2G1B
COX6A1
RNF10
CABP1
SPPL3
RAB35
RPLP0
SIRT4
TRIAP1
POP5
MLEC
XLOC_009911
MIR4498
MSI1
GATC
UNC119B
PXN−AS1
SRSF9
MIR4700
DYNLL1
ACADS
NRAV
COQ5
BCL7A
LRRC43
CLIP1
ZCCHC8
HCAR2
CCDC62
ABCB9
LOC100506691
IL31
CLIP1−AS1
RSRC2
HCAR3
HIP1R
MLXIP
B3GNT4
KNTC1
HCAR1
VPS37B
DIABLO
DENR
LOC101593348
1 gene omitted

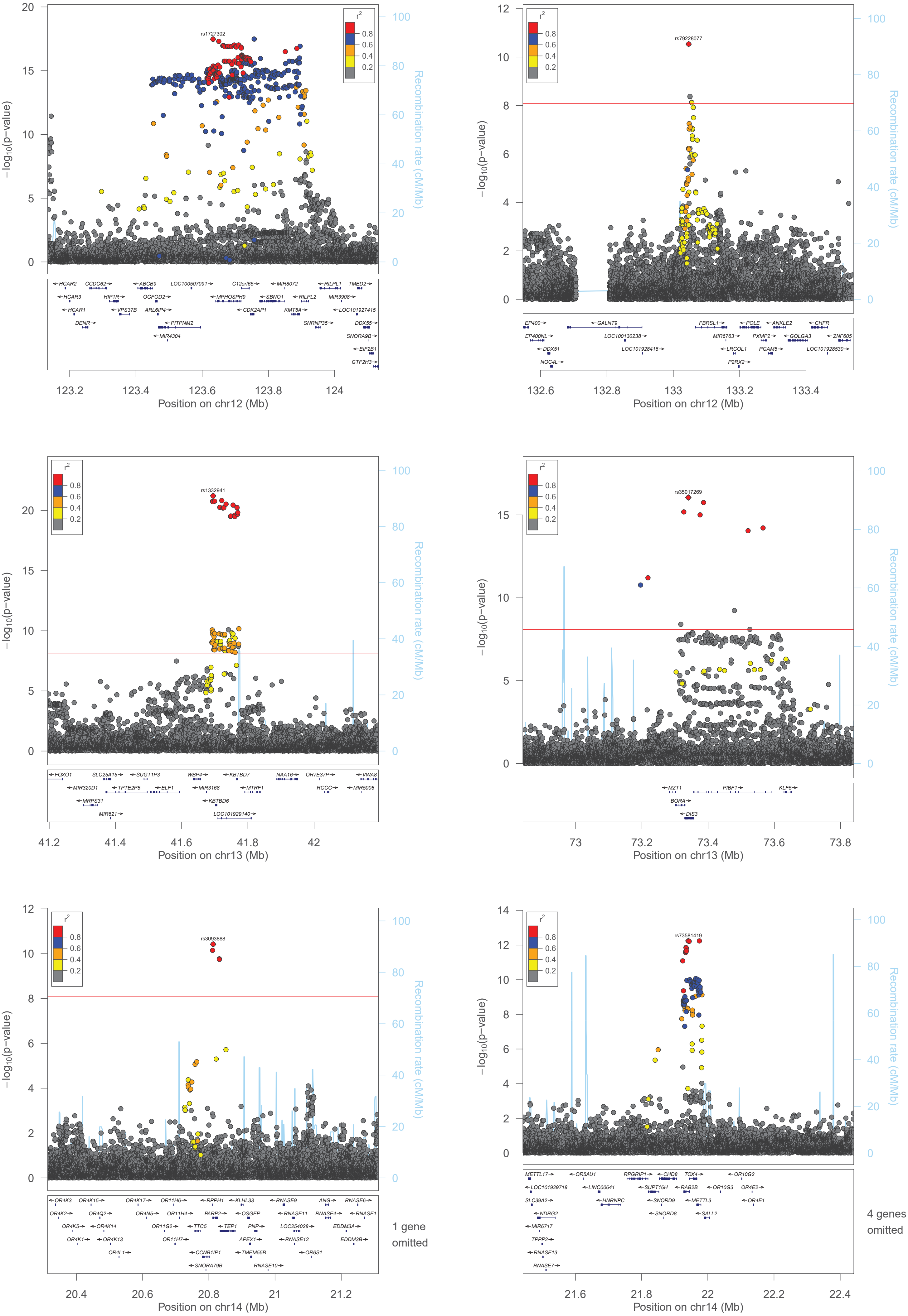

rs1727302
−log10(p−value)
Recombination rate (cM/Mb)
Position on chr12 (Mb)
rs79228077
Position on chr12 (Mb)
rs1332941
Position on chr13 (Mb)
rs35017269
Position on chr13 (Mb)
rs3093888
1 gene omitted
Position on chr14 (Mb)
rs73581419
4 genes omitted
Position on chr14 (Mb)

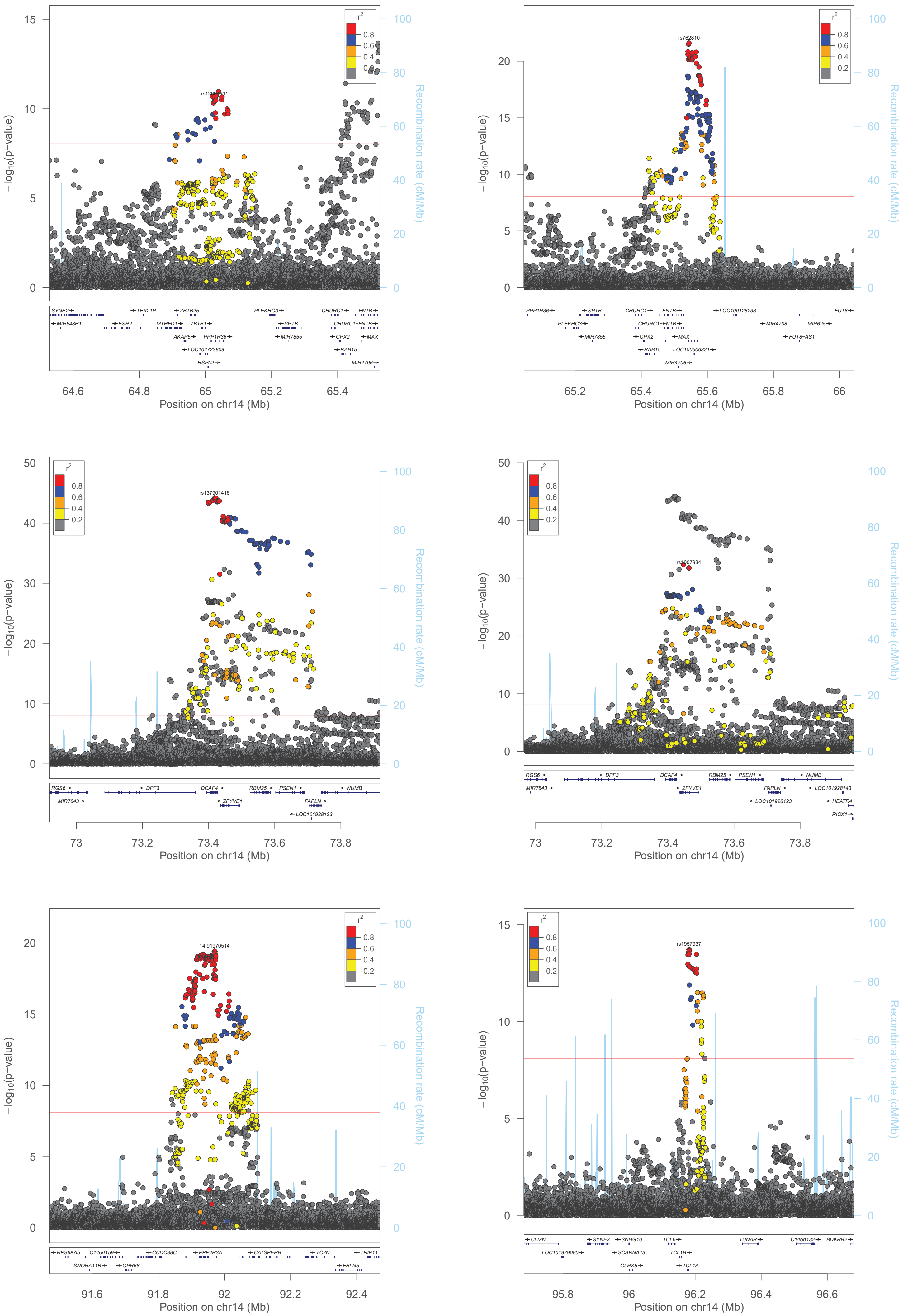

rs12586411
rs762810
rs137901416
rs1007934
14:91970514
rs1957937
−log10(p−value)
Recombination rate (cM/Mb)
Position on chr14 (Mb)
r2
0.8
0.6
0.4
0.2

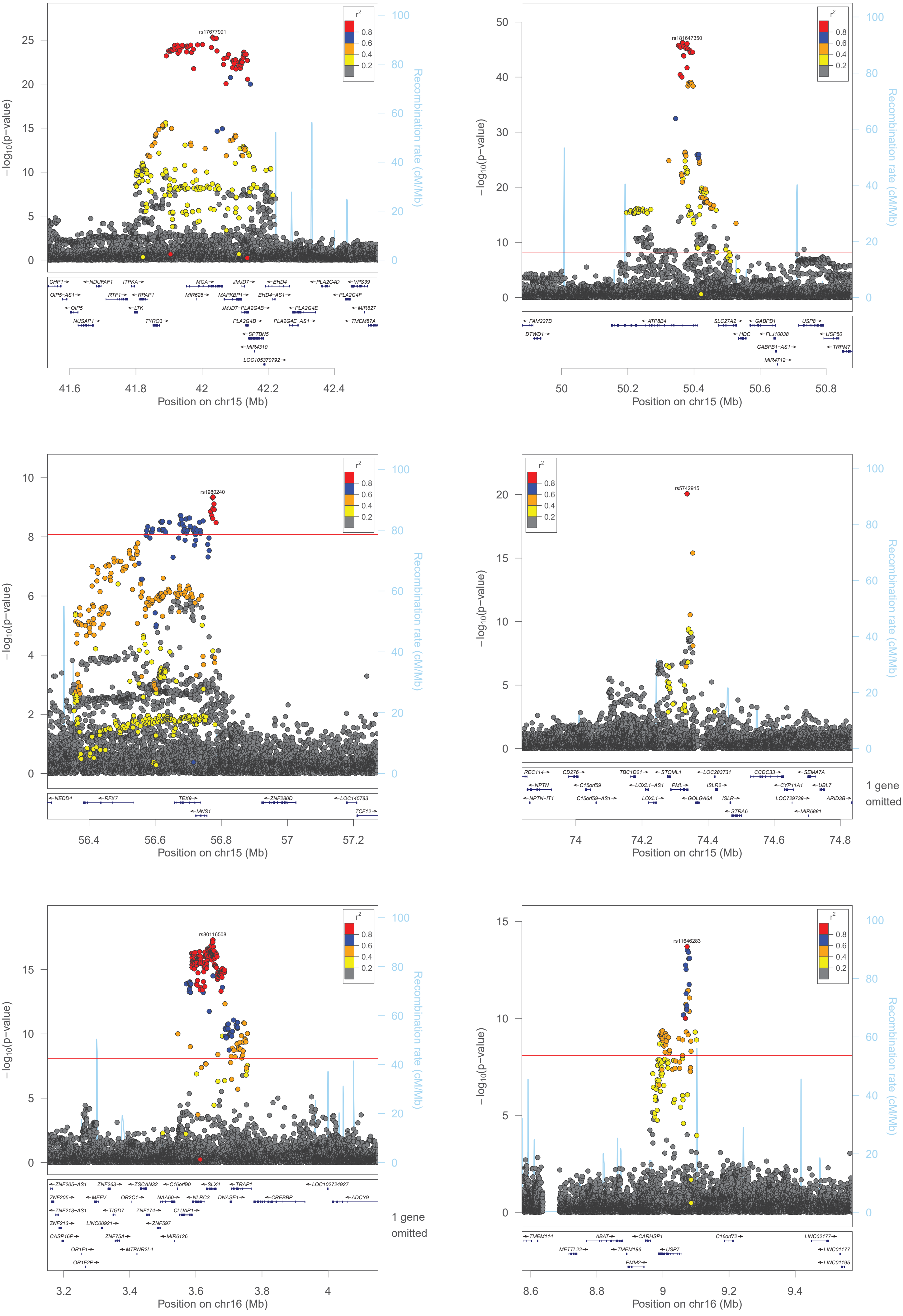

rs17677991
−log10(p−value)
Recombination rate (cM/Mb)
Position on chr15 (Mb)
rs181647350
Position on chr15 (Mb)
rs1980240
Position on chr15 (Mb)
rs5742915
1 gene omitted
Position on chr15 (Mb)
rs80116508
1 gene omitted
Position on chr16 (Mb)
rs11646283
Position on chr16 (Mb)

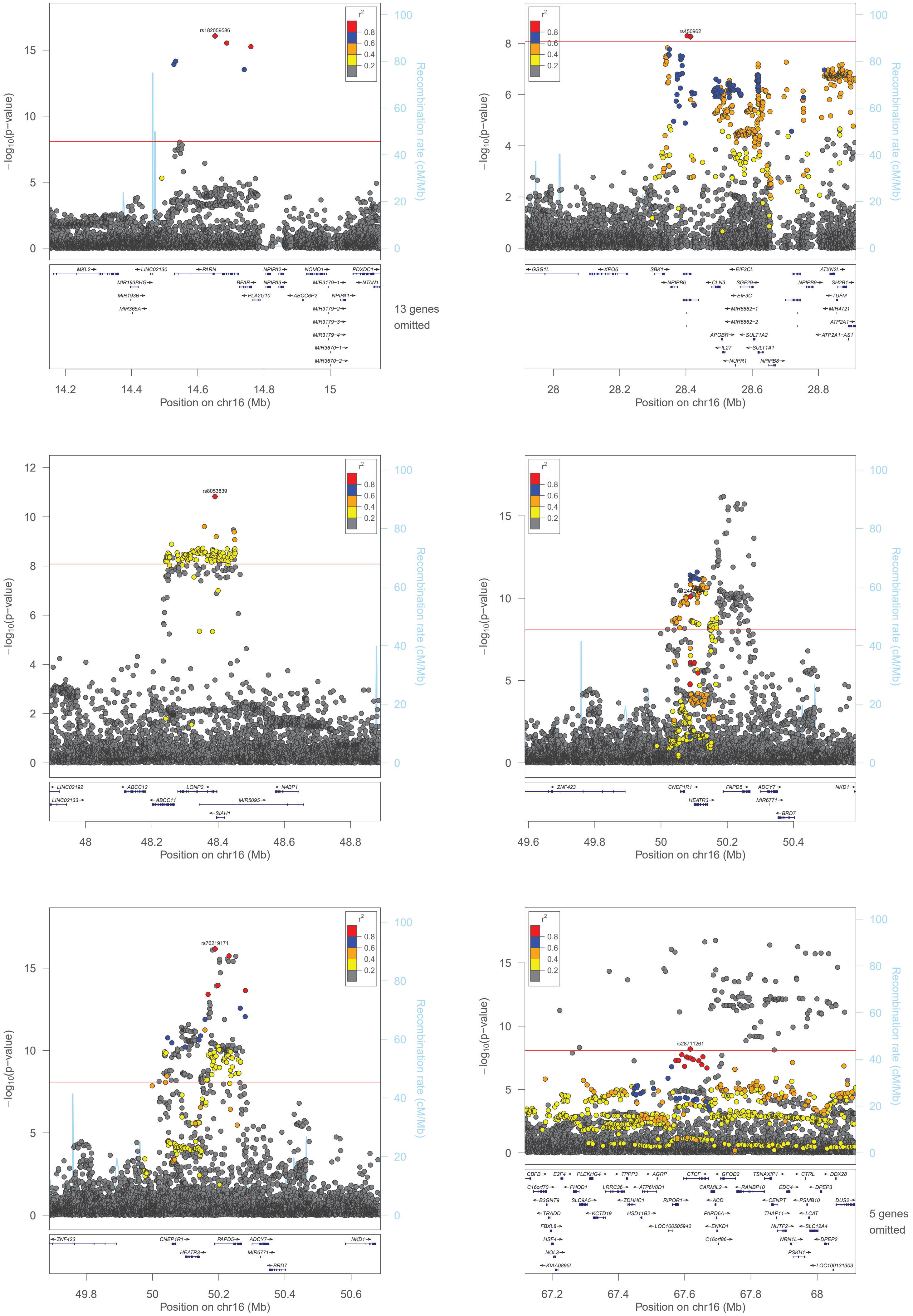

rs182059586
r2
0.8
0.6
0.4
0.2
−log10(p−value)
Recombination rate (cM/Mb)
MKL2
LINC02130
PARN
NPIPA2
NOMO1
PDXDC1
MIR193BHG
BFAR
NPIPA3
MIR3179−1
NTAN1
MIR193B
PLA2G10
ABCC6P2
NPIPA1
MIR365A
MIR3179−2
MIR3179−3
MIR3179−4
MIR3670−1
MIR3670−2
13 genes omitted
Position on chr16 (Mb)
rs450962
GSG1L
XPO6
SBK1
EIF3CL
ATXN2L
NPIPB6
CLN3
SGF29
NPIPB9
SH2B1
EIF3C
TUFM
MIR6862−1
MIR4721
MIR6862−2
ATP2A1
APOBR
SULT1A2
ATP2A1−AS1
IL27
SULT1A1
NUPR1
NPIPB8
Position on chr16 (Mb)
rs8053839
LINC02192
ABCC12
LONP2
N4BP1
LINC02133
ABCC11
MIR5095
SIAH1
Position on chr16 (Mb)
ZNF423
CNEP1R1
PAPD5
ADCY7
NKD1
HEATR3
MIR6771
BRD7
Position on chr16 (Mb)
rs76219171
ZNF423
CNEP1R1
PAPD5
ADCY7
NKD1
HEATR3
MIR6771
BRD7
Position on chr16 (Mb)
rs28711261
CBFB
E2F4
PLEKHG4
TPPP3
AGRP
CTCF
GFOD2
TSNAXIP1
CTRL
DDX28
C16orf70
FHOD1
LRRC36
ATP6V0D1
CARMIL2
RANBP10
EDC4
DPEP3
B3GNT9
SLC9A5
ZDHHC1
RIPOR1
ACD
CENPT
PSMB10
DUS2
TRADD
KCTD19
HSD11B2
PARD6A
THAP11
LCAT
FBXL8
LOC100505942
ENKD1
NUTF2
SLC12A4
HSF4
C16orf86
NRN1L
DPEP2
NOL3
PSKH1
KIAA0895L
LOC100131303
5 genes omitted
Position on chr16 (Mb)

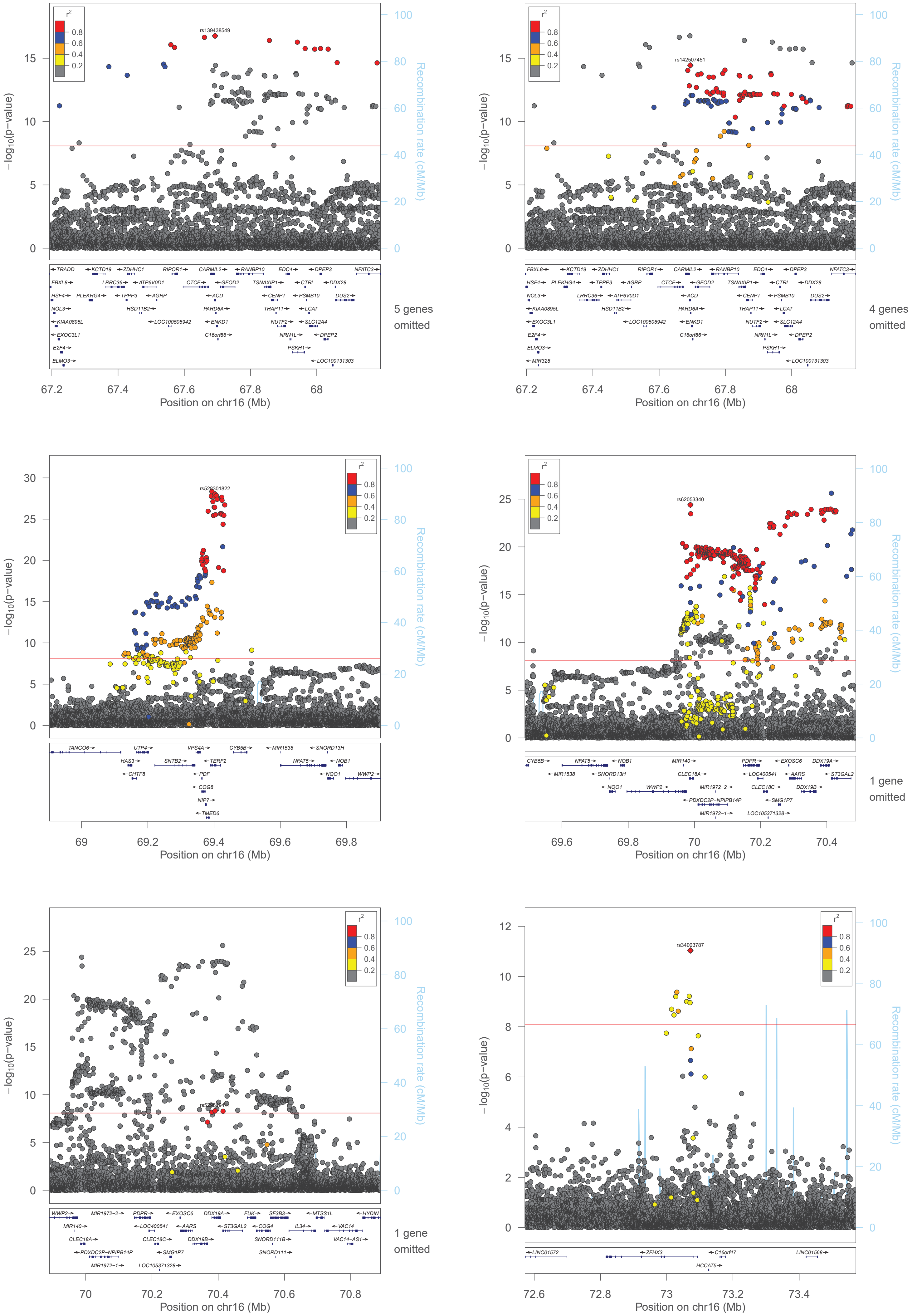
rs139438549
rs142507451
rs528301822
rs62053340
rs34003787
r2
0.8
0.6
0.4
0.2
−log10(p−value)
Recombination rate (cM/Mb)
Position on chr16 (Mb)
5 genes omitted
4 genes omitted
1 gene omitted
1 gene omitted

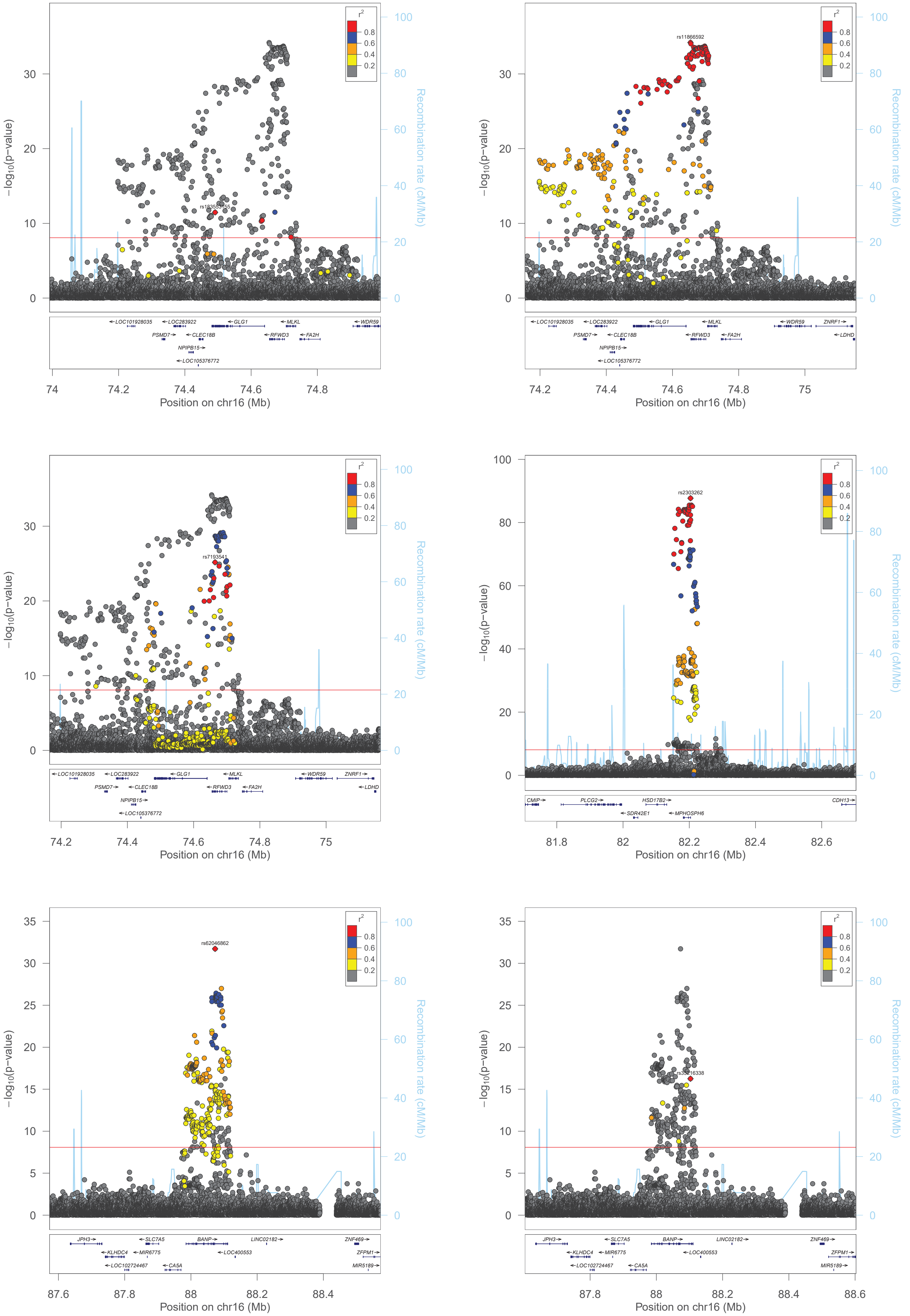
−log10(p−value)
Recombination rate (cM/Mb)
Position on chr16 (Mb)
r2
0.8
0.6
0.4
0.2
rs11866592
rs7193541
rs2303262
rs62046862
LOC101928035
LOC283922
GLG1
MLKL
WDR59
ZNRF1
PSMD7
CLEC18B
RFWD3
FA2H
LDHD
NPIPB15
LOC105376772
CMIP
PLCG2
HSD17B2
CDH13
SDR42E1
MPHOSPH6
JPH3
SLC7A5
BANP
LINC02182
ZNF469
KLHDC4
MIR6775
LOC400553
ZFPM1
LOC102724467
CA5A
MIR5189

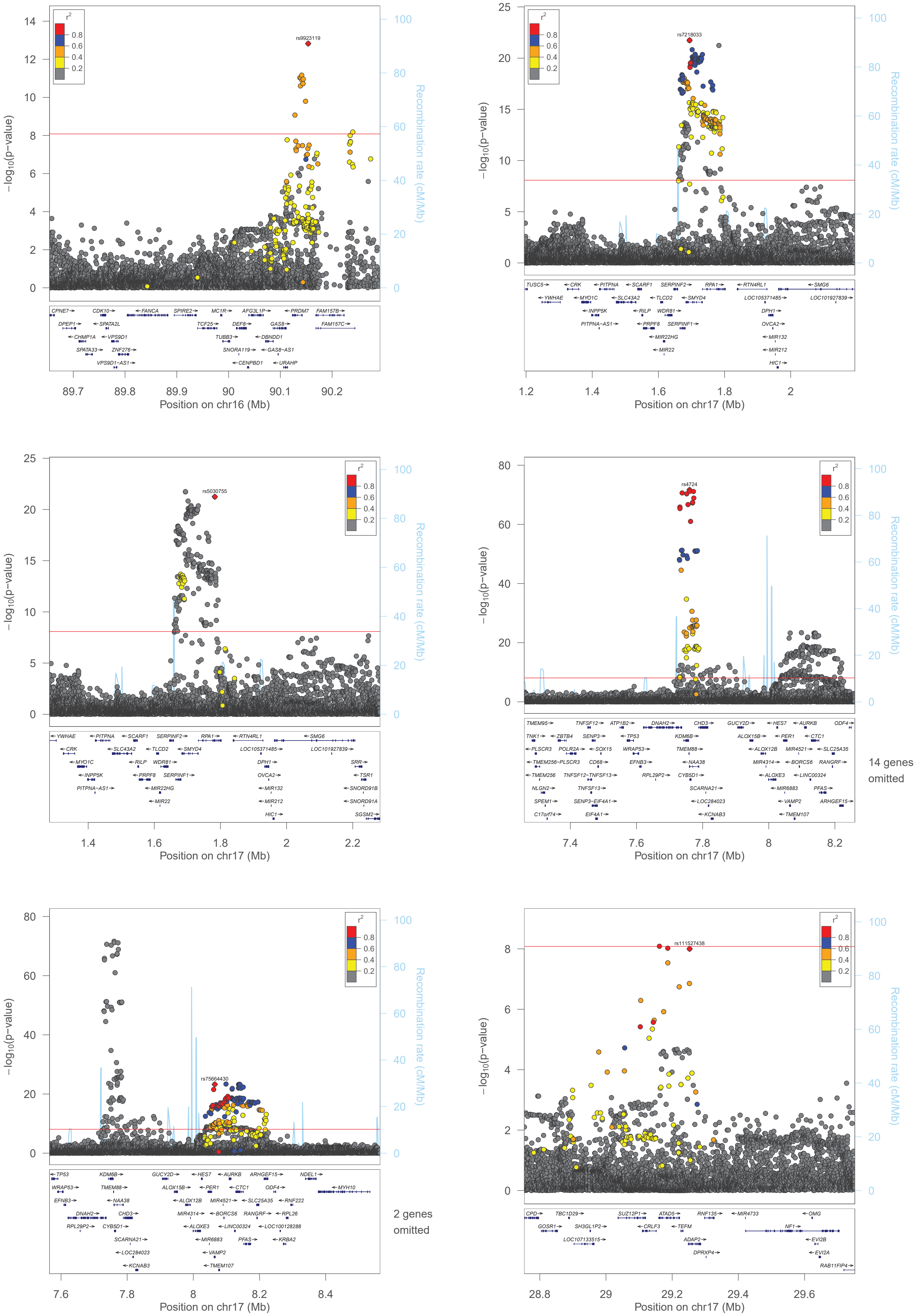

rs9923119
−log10(p−value)
Recombination rate (cM/Mb)
Position on chr16 (Mb)
rs7218033
Position on chr17 (Mb)
rs5030755
Position on chr17 (Mb)
rs4724
14 genes omitted
Position on chr17 (Mb)
rs75664430
2 genes omitted
Position on chr17 (Mb)
rs111527438
Position on chr17 (Mb)

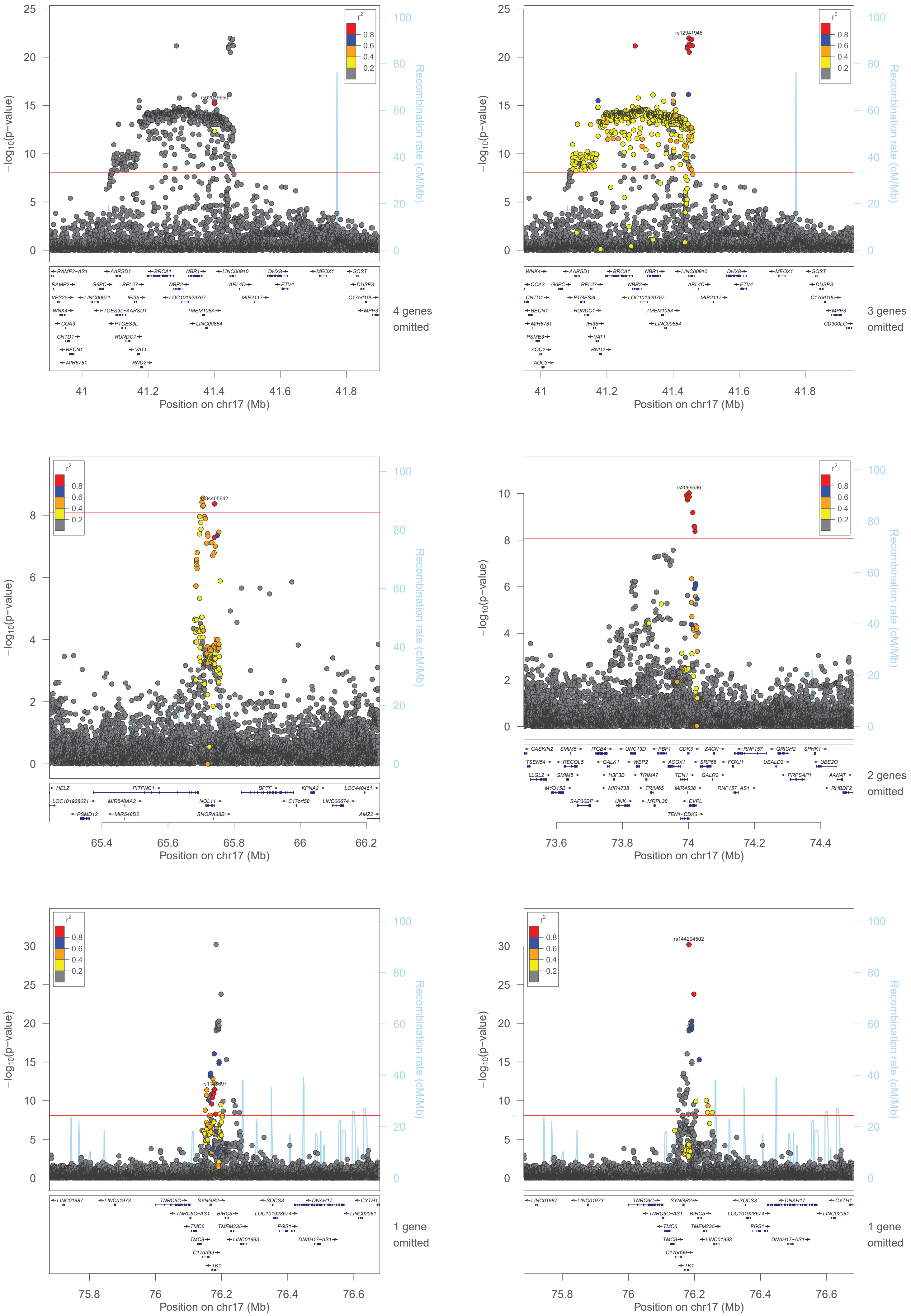

−log10(p−value)
Recombination rate (cM/Mb)
Position on chr17 (Mb)
r2
0.8
0.6
0.4
0.2
rs62079650
rs12941945
rs34405642
rs2069536
rs144204502
4 genes omitted
3 genes omitted
2 genes omitted
1 gene omitted
1 gene omitted

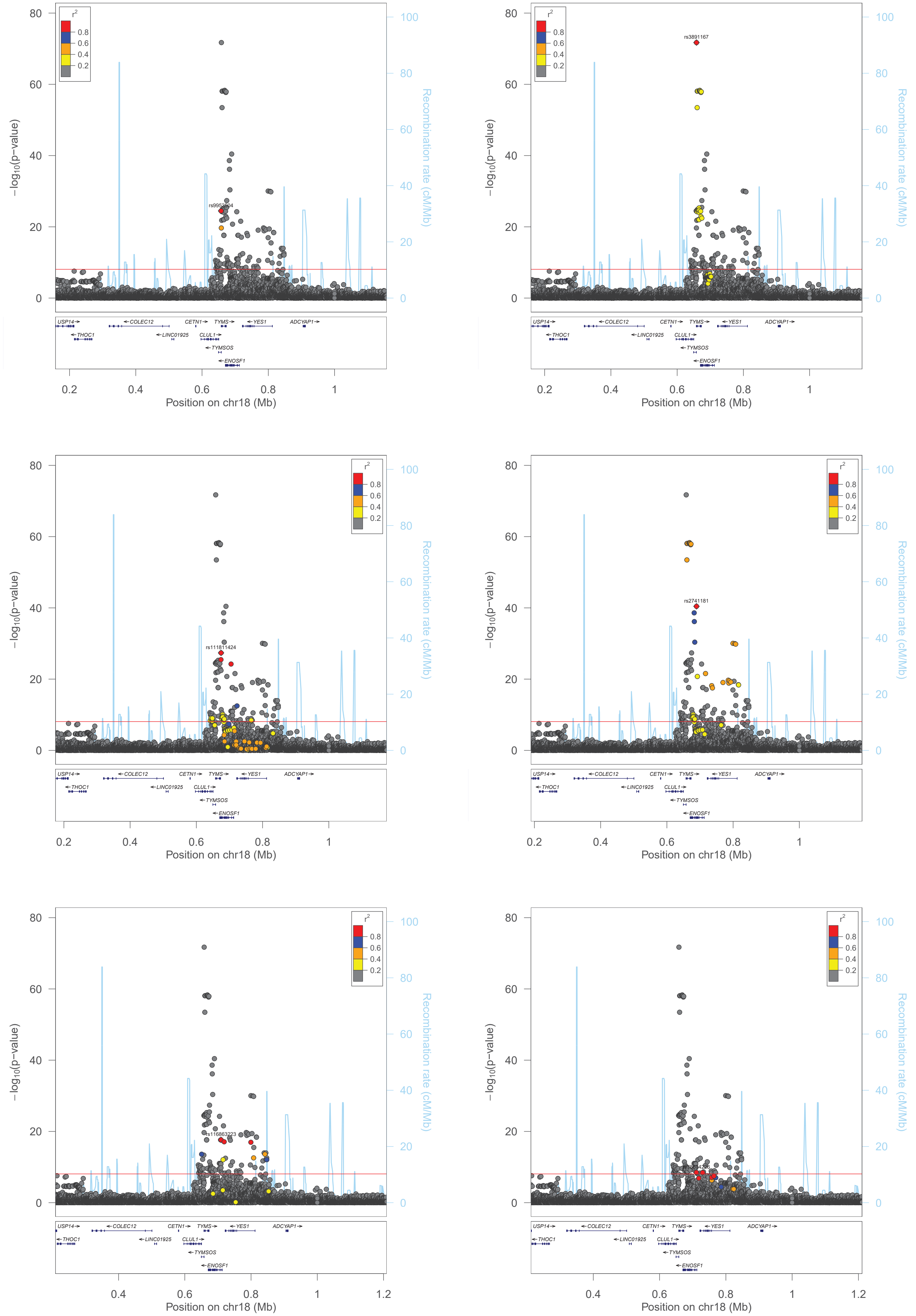

r2
0.8
0.6
0.4
0.2
−log10(p-value)
Recombination rate (cM/Mb)
Position on chr18 (Mb)
rs9952504
rs3891167
rs111811424
rs2741181
rs116863223
USP14
THOC1
COLEC12
LINC01925
CETN1
CLUL1
TYMS
TYMSOS
ENOSF1
YES1
ADCYAP1

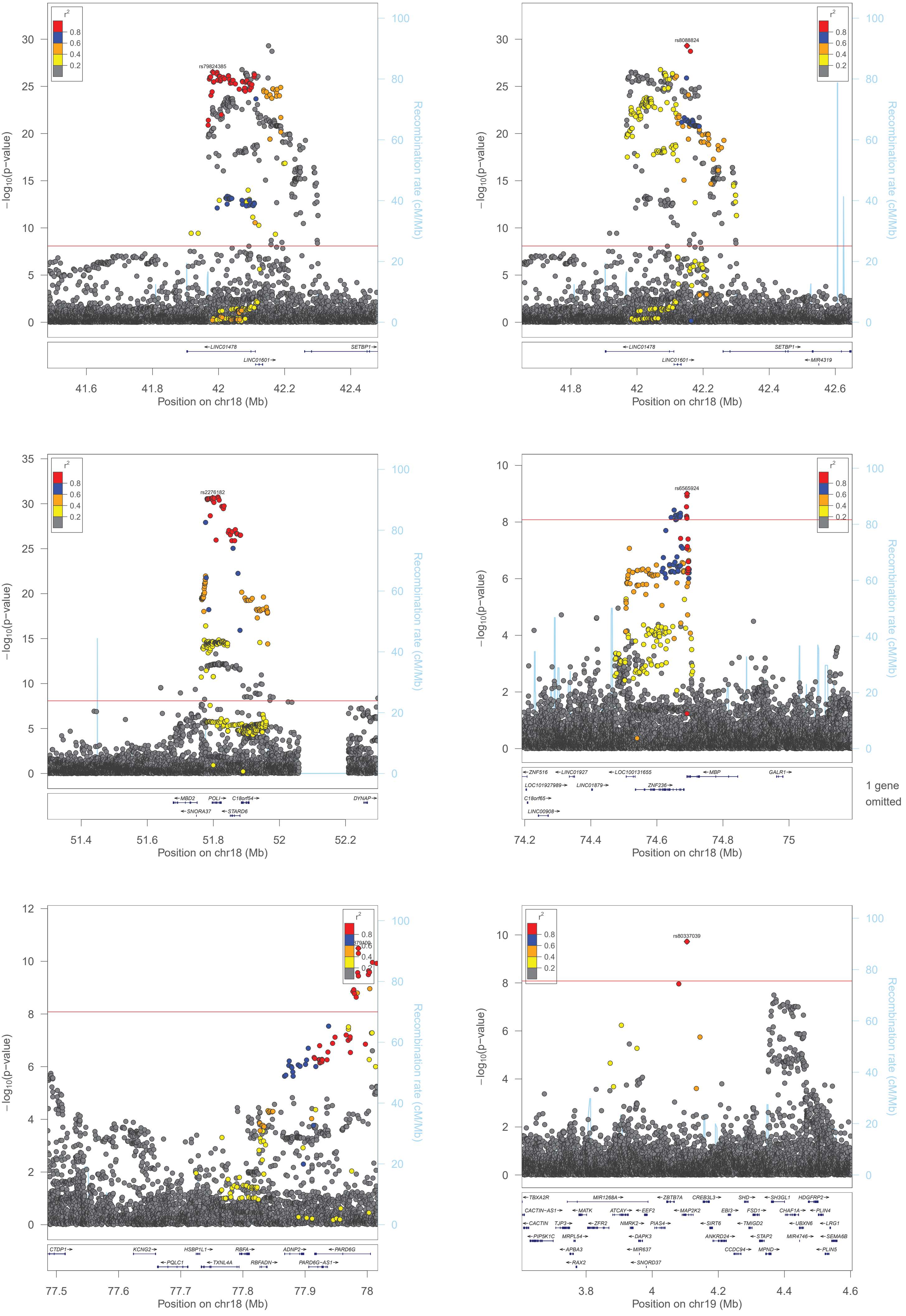

r2
0.8
0.6
0.4
0.2
rs79824385
rs8088824
rs2276182
rs6565924
rs80337039
−log10(p−value)
Recombination rate (cM/Mb)
Position on chr18 (Mb)
Position on chr19 (Mb)
LINC01478
LINC01601
SETBP1
MIR4319
MBD2
POLI
C18orf54
DYNAP
SNORA37
STARD6
ZNF516
LINC01927
LOC100131655
MBP
GALR1
LOC101927989
LINC01879
ZNF236
C18orf65
LINC00908
1 gene omitted
CTDP1
KCNG2
HSBP1L1
RBFA
ADNP2
PARD6G
PQLC1
TXNL4A
RBFADN
PARD6G−AS1
TBXA2R
MIR1268A
ZBTB7A
CREB3L3
SHD
SH3GL1
HDGFRP2
CACTIN−AS1
MATK
ATCAY
EEF2
MAP2K2
EBI3
FSD1
CHAF1A
PLIN4
CACTIN
TJP3
ZFR2
NMRK2
PIAS4
SIRT6
TMIGD2
UBXN6
LRG1
PIP5K1C
MRPL54
DAPK3
ANKRD24
STAP2
MIR4746
SEMA6B
APBA3
MIR637
CCDC94
MPND
PLIN5
RAX2
SNORD37

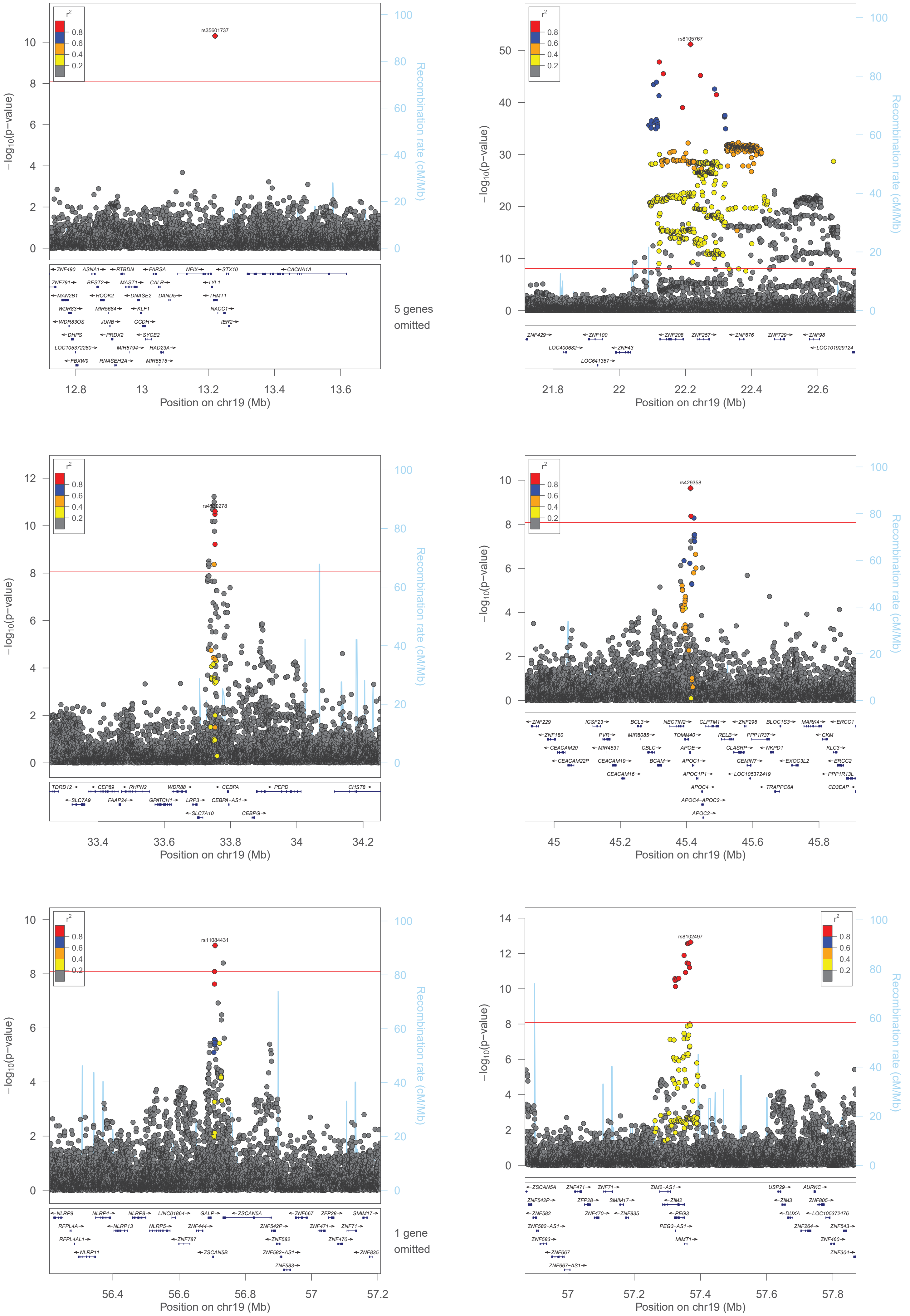

rs35601737
rs8105767
rs4420278
rs429358
rs11084431
rs8102497
r2
0.8
0.6
0.4
0.2
−log10(p−value)
Recombination rate (cM/Mb)
Position on chr19 (Mb)
5 genes omitted
1 gene omitted

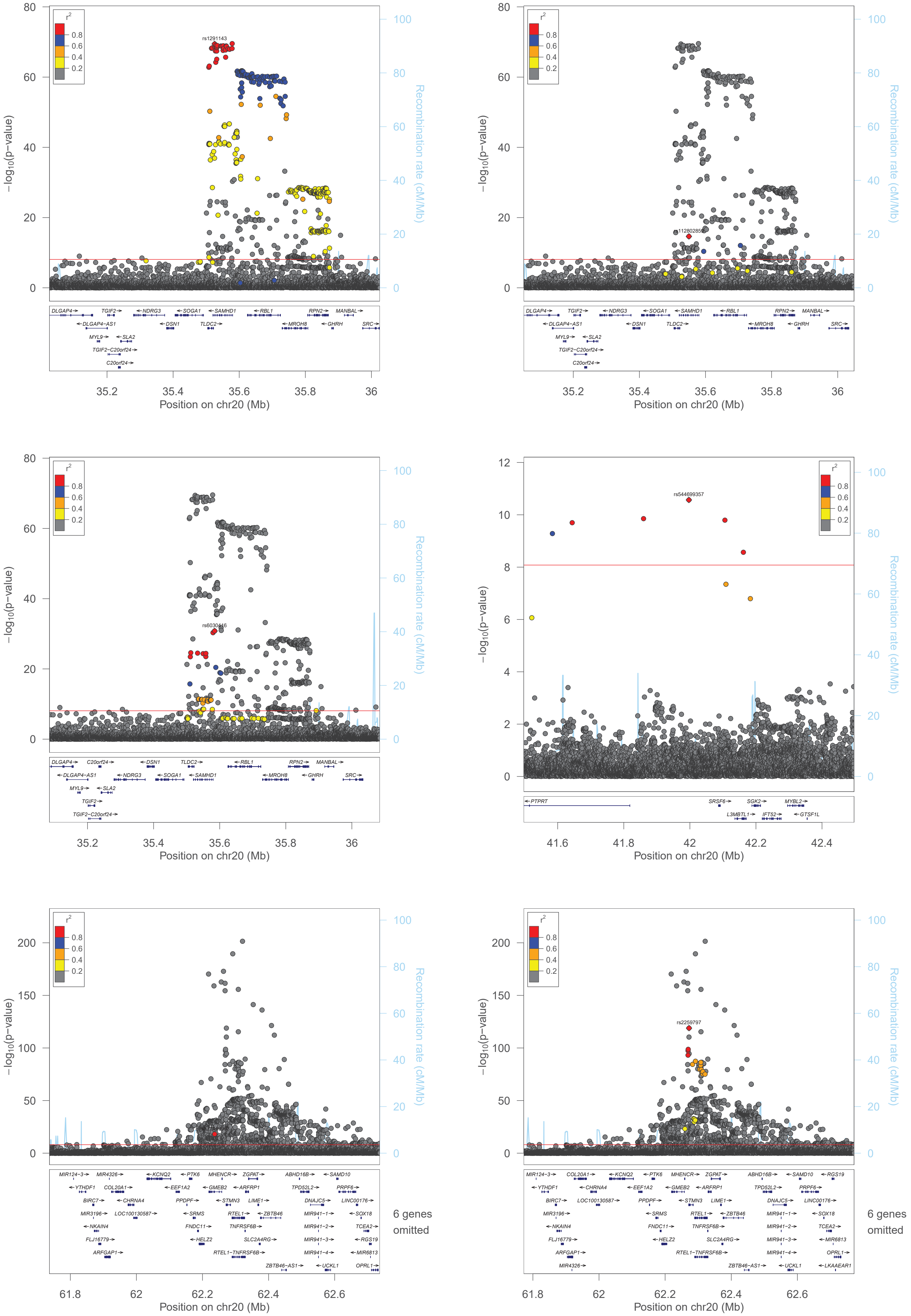

−log10(p−value)
Recombination rate (cM/Mb)
Position on chr20 (Mb)
r2
0.8
0.6
0.4
0.2
rs1291143
rs544699357
rs2259797
6 genes omitted

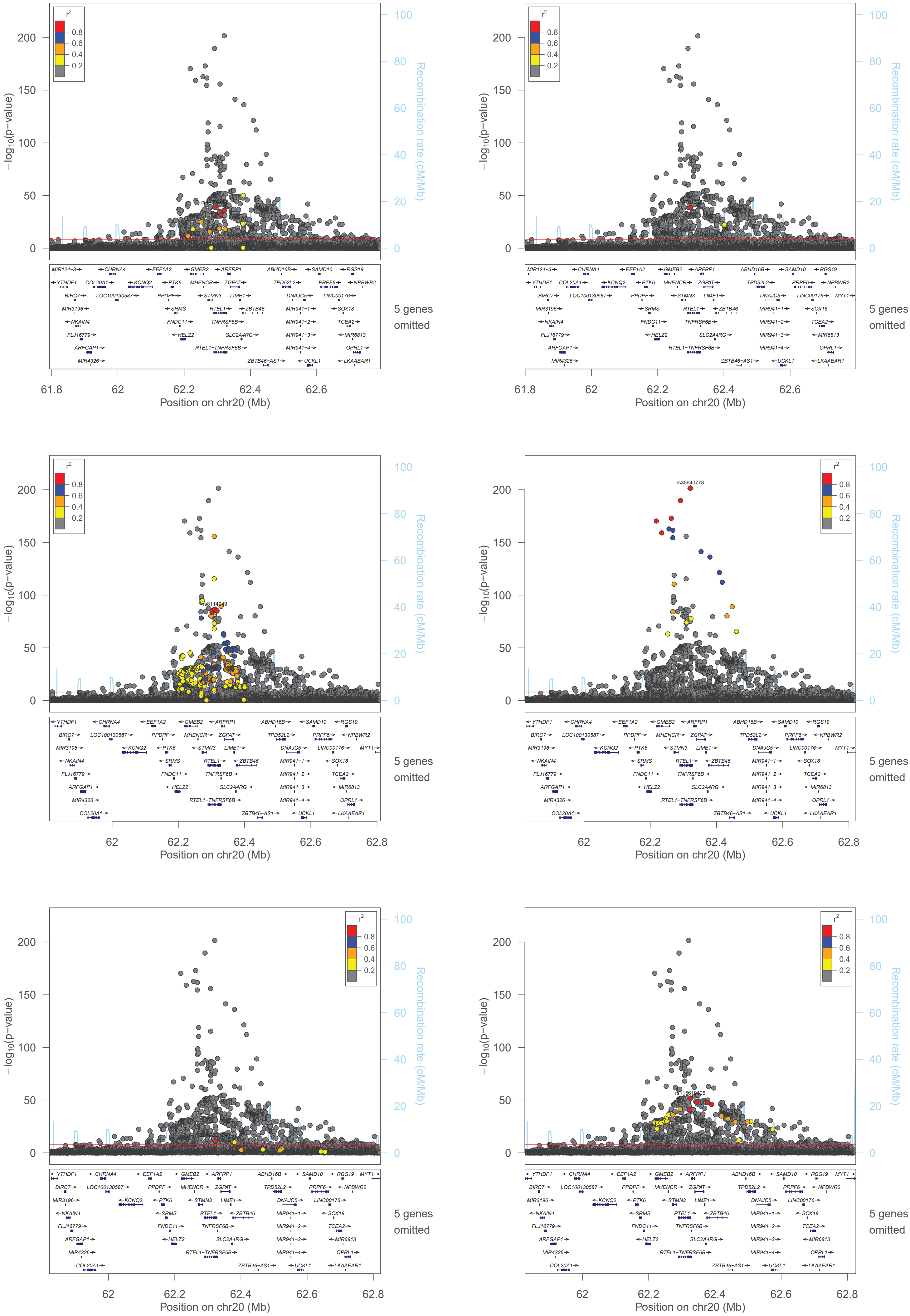

−log10(p−value)
Recombination rate (cM/Mb)
Position on chr20 (Mb)
r2
0.8
0.6
0.4
0.2
5 genes omitted
rs8114049
rs35640778

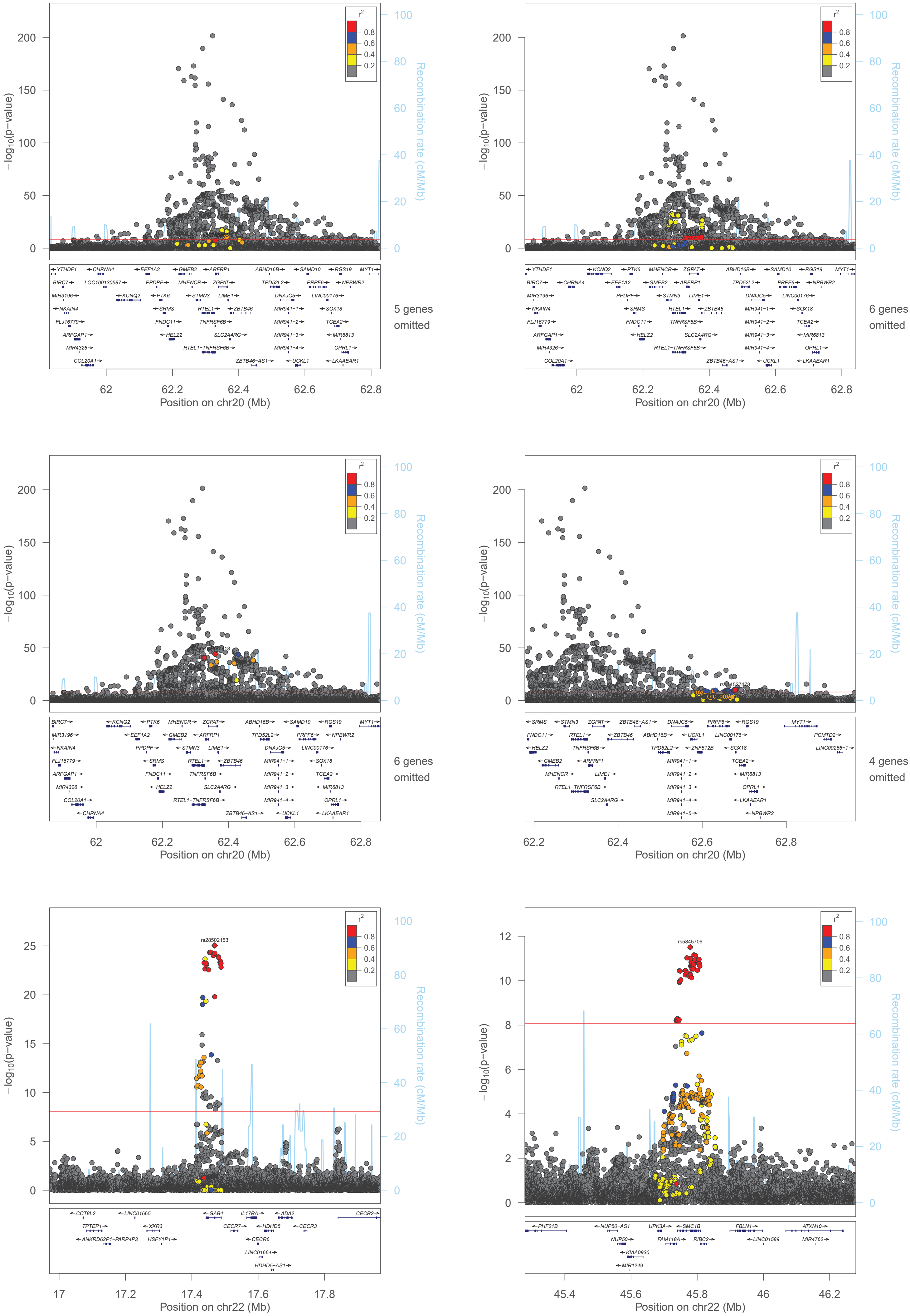

−log10(p-value)
Recombination rate (cM/Mb)
r2
0.8
0.6
0.4
0.2
5 genes omitted
6 genes omitted
6 genes omitted
4 genes omitted
Position on chr20 (Mb)
Position on chr20 (Mb)
Position on chr20 (Mb)
Position on chr20 (Mb)
rs28502153
rs5845706
Position on chr22 (Mb)
Position on chr22 (Mb)

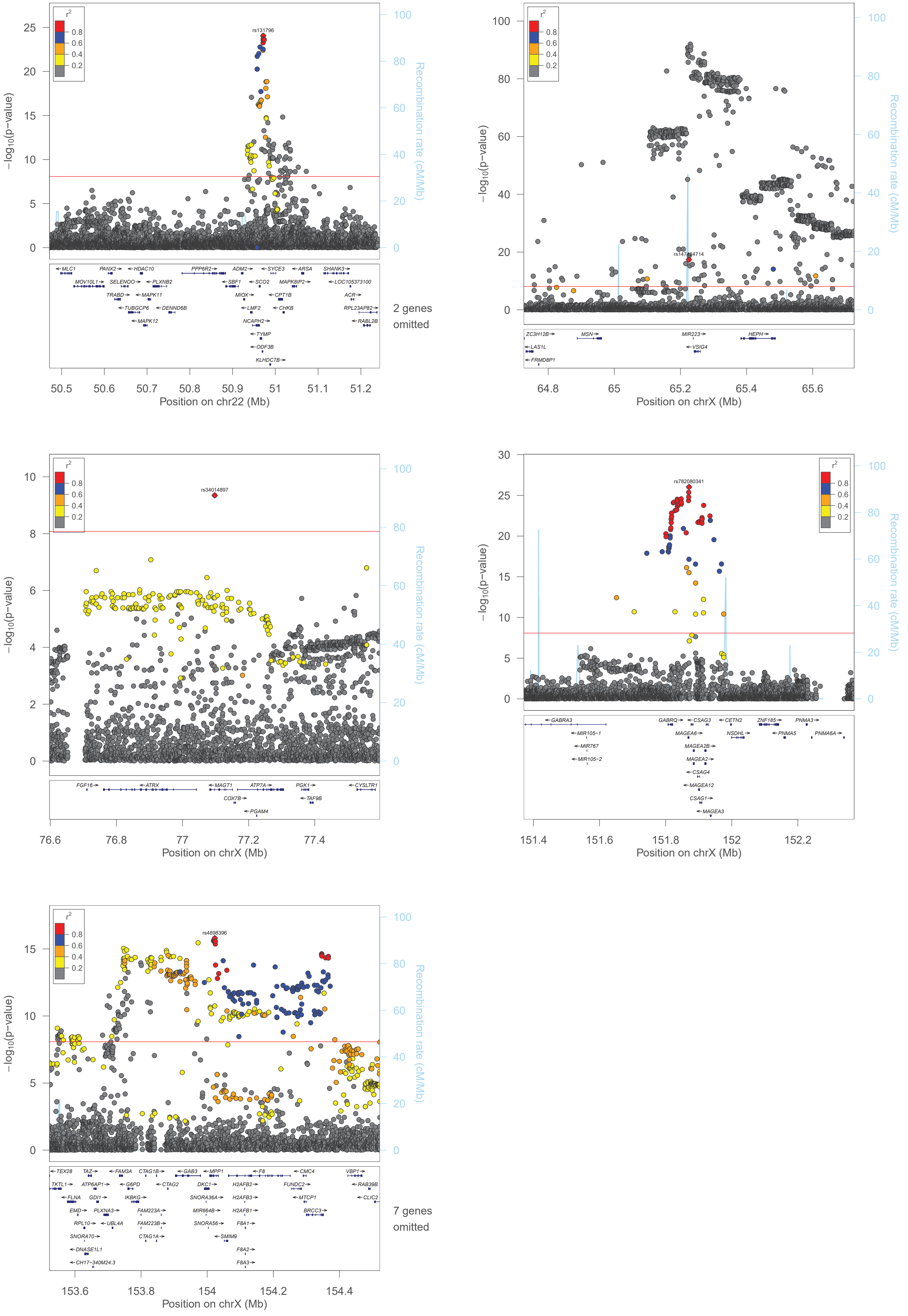

r2
0.8
0.6
0.4
0.2
−log10(p−value)
Recombination rate (cM/Mb)
rs131796
MLC1
PANX2
HDAC10
PPP6R2
ADM2
SYCE3
ARSA
SHANK3
MOV10L1
SELENOO
PLXNB2
SBF1
SCO2
MAPK8IP2
LOC105373100
TRABD
MAPK11
MIOX
CPT1B
ACR
TUBGCP6
DENND6B
LMF2
CHKB
RPL23AP82
MAPK12
NCAPH2
RABL2B
TYMP
ODF3B
KLHDC7B
2 genes omitted
Position on chr22 (Mb)
rs147424714
ZC3H12B
MSN
MIR223
HEPH
LAS1L
VSIG4
FRMD8P1
Position on chrX (Mb)
rs34014897
FGF16
ATRX
MAGT1
ATP7A
PGK1
CYSLTR1
COX7B
TAF9B
PGAM4
Position on chrX (Mb)
rs782080341
GABRA3
GABRQ
CSAG3
CETN2
ZNF185
PNMA3
MIR105−1
MAGEA6
NSDHL
PNMA5
PNMA6A
MIR767
MAGEA2B
MIR105−2
MAGEA2
CSAG4
MAGEA12
CSAG1
MAGEA3
Position on chrX (Mb)
rs4898396
TEX28
TAZ
FAM3A
CTAG1B
GAB3
MPP1
F8
CMC4
VBP1
TKTL1
ATP6AP1
G6PD
CTAG2
DKC1
H2AFB2
FUNDC2
RAB39B
FLNA
GDI1
IKBKG
SNORA36A
H2AFB3
MTCP1
CLIC2
EMD
PLXNA3
FAM223A
MIR664B
H2AFB1
BRCC3
RPL10
UBL4A
FAM223B
SNORA56
F8A1
SNORA70
CTAG1A
SMIM9
DNASE1L1
F8A2
CH17−340M24.3
F8A3
7 genes omitted
Position on chrX (Mb)
