## Supplementary Information and Figures for "Polygenic basis and biomedical consequences of telomere length variation"

V. Codd et al.

### Contents

| | |
|---|---|
| Investigation of rs334 and rs1609812 | 2 |
| Identifying likely-causal genes within conditionally independent GWS loci | 3 |
| Analysis of rare and ultra-rare variants in gene based tests | 4 |
| Association of LTL with biomedical traits | 5 |
| Investigating disease association non-concordance. | 6 |
| Supplementary methods | 7 |
| Supplementary references | 13 |
| Supplementary figures | 14 |

**Investigation of rs334 and rs1609812**

The Hgbu and Hgbd primers used for the multiplex qPCR telomere length (TL) measurement[12] bind within the HBB gene, where rs334 is located. Aligning the primer sequences to the genomic sequence with annotation of known SNPs using the UCSC genome browser (**Supplementary Figure 3**) reveals that rs334 is located 2bp from the 5' end of the reverse primer (Hgbd). Although the 5' end of primer sequences is less critical for primer binding, these primers also carry a large GC rich non-binding tail. Reduction of the primer binding capacity from 18bp to 16bp, when there is already a large tail, could, therefore, reduce efficiency of the reaction, leading to underestimation of the single copy repeat number calculated (S). Underestimating (S) would subsequently inflate the T/S ratio used as the leucocyte TL (LTL) measurement. The association we observed of the rs334 minor allele with longer LTL is consistent with this hypothesis.

An additional sentinel variant, rs1609812, was identified in the same locus as rs334. To investigate this association further, we excluded rs334 and any participant with a diagnosis of thalassaemia, as other thalassaemia mutations in this region are located within the primer sequences (**Supplementary Figure 3**), and re-tested the association of rs1609812 with LTL. The original association (beta=0.043, $P=4.88 \times 10^{-53}$) is unaffected by rs334 carrier removal (beta=0.044, $P=5.32 \times 10^{-54}$) using the data prior to conditional analyses.

To investigate associations of rs1609812 with LTL further (particularly given the contrast between the strong association observed in our study and the lack of evidence from prior publications), we examined two separate large datasets, neither of which showed significant associations (P=0.04 and P=0.01 in Li et al., 2020[4] and Dorajoo et al., 2019[5] respectively). Both previous studies used different single copy genes in the TL assay (36B4 and albumin). Although we have no direct evidence that rs1609812 or correlated variants influence the S measurement, the lack of supportive evidence from studies using a different gene to calculate S values suggests it may be an artefactual association. As such, our results related to this locus should be interpreted with caution.

**Identifying likely-causal genes within conditionally independent GWAS loci**

To identify potential causal variants at each locus, we performed fine-mapping using a two-step approach in FINEMAP that allowed for more than one causal SNP, annotating the variants in the 95% credible sets to prioritise those with likely functional impact at loci that contained more than one independent sentinel (**Methods, Supplementary Table 2**). We then used the credible sets (a set of SNPs that have a 95% probability of containing the casual SNP, **Supplementary Table 2**) to identify the genes that are most likely influenced by the LTL associated variants. We first functionally annotated the credible set SNPs to identify those variants that are most likely to have an impact on gene function (**Methods**). In total, 72 variants, including 24 identified as being the most likely causal variant within a locus, were annotated as having protein-altering consequences (**Supplementary Table 3**). These were distributed across 44 loci, with the majority highlighting a single gene per locus. One exception is the RTEL1 locus, where four likely-causal SNPs are all located within both *RTEL1* and the *RTEL1-TNFRSF6B* readthrough transcript. However, these are most likely to only have a functional consequence within RTEL1 as the *RTEL1-TNFRSF6B* transcript is annotated as likely undergoing nonsense mediated decay and therefore not translated. Several genes contained more than one coding variant, most notably *SLX4* which contained seven, although only one was annotated as being damaging across multiple prediction tools (**Supplementary Table 3**).

To identify variants acting through altered gene expression, we searched for evidence of a shared causal variant influencing both LTL and gene expression using eQTL data from 48 tissues from the GTEx Consortium (**Methods**, **Supplementary Table 4**). We found evidence of colocalising eQTLs at 93 (68%) of the loci (**Supplementary Tables 4** and **5**). For several genes, there was strong evidence of a colocalised eQTL across a number of different tissues. Among the most striking were *RPA1*, *TEN1*, *POLI* and *AC011498*.6, which showed evidence of strong, ubiquitous colocalisation (**Supplementary Table 4**, **Figure 2A**). There were also tissue specific signals for *IRF5, MPHOSPH6, AC007292.1, AC145285.2, LONP2, MCM4, CDK3, PPP1CB* and *TRMT61B*. *PPP1CB* and *TRMT61B* are within the same locus and show similar levels of colocalisation across different tissues. *AC011498*.6 and *AC007292*.1 are also within the same locus, as are *TEN1* and *CDK3*, with the strength of evidence pointing more towards the gene in the ubiquitous grouping (*TEN1* and *AC011498*.6).

Although many components of SHELTERIN and telomerase were within our conditionally independent GWS loci, not all were found. The two remaining SHELTERIN components *TINF2* and *TERF2IP* along with two further core telomerase genes *NOP10* and *GAR1* are within loci identified at <1%FDR (rs34354104, rs6564260, rs2615358 and rs73839158 respectively, **Supplementary Table 2**), Variants around *TINF2* have been associated with LTL previously, but only in a Singaporean Chinese population[5]. We also observed the previous *MOB1B* locus within the <1%FDR[4]. Although we believe there are loci genuinely associated with LTL within the FDR, this list may also contain more false-positive results than the estimated 1% due to the large number of variants being tested and the influence of LD on the FDR approach. We therefore have not undertaken further analyses on these variants at this stage and advise cautious interpretation of them.

**Analysis of rare and ultra-rare variants in gene based tests**

To understand the direction of association of genes associated with LTL in our gene based tests, we assessed the association of the contributing individual variants in single variant analyses. Where individual variants within a gene differed in direction of effect we see consistency with the effect of the gene score for variants most likely to be true LoF (truncating or frameshift variants in early exons) and where minor allele counts are >10 (where estimates of individual variant effects are likely to be more reliable, **Supplementary Table 8**).

High confidence truncating or protein-altering variants throughout RTEL1 were mostly associated with shorter LTL, consistent with data suggesting that the full length RTEL1 protein is required to facilitate telomere elongation by telomerase[29]. The direction of effect is also consistent with the known biological roles of TERF1, POT1, TERT and PARN. TERF1 and POT1 are both components of SHELTERIN and limit telomere elongation by restricting access of the telomere end to telomerase[65,66]. For both, LoF variants result in longer LTL. Reduction of TERT or PARN results in lower telomerase activity reflecting the shorter LTL effect observed (**Supplementary Table 8)**[21]. Our results suggest reduction in SAMHD1 results in longer LTL. Whilst reduction in SAMHD1 can result in telomere fragility, this has only been shown in the absence of TERF1[28]. A major role of SAMHD1 is in regulating cellular dNTP pools, especially dGTP levels. Loss of SAMHD1 results in elevated dGTP and dGTP levels have been seen to positively correlate with both telomerase activity and TL[67,68].

Within ATM there was less consistency in the direction of effect of single variants (**Supplementary Table 8**). Truncating variants located in early exons associated with both longer and shorter LTL. However, those variants with MAC>10, where we have more confidence in the single variant level data, were consistently associated with shorter LTL, which reflects previous studies showing loss of ATM resulting in short TL[69].

Loss of CTC1 results in diseases characterised as short TL syndromes, however this requires biallelic mutation, presumably resulting in complete LoF of CTC1. Our results suggest that partial loss of CTC1 can result in moderate LTL lengthening. One possible explanation is that, like SHELTERIN, the CST complex limits telomerase activity at the telomere end and loss results in more accessibility and therefore lengthening[14].

A further 141 genes were nominally significant ($p<0.05$) and included both candidate genes (*TP53*, *PML*, *WRAP53*, *ACD*), one gene involved in nucleotide metabolism (*TYMS*) and *SMC1B*, a component of the cohesin complex that protects from telomere shortening during meiotic cell division[70].

**Association of LTL with biomedical traits**

Longer usual LTL was associated with a higher peak expiratory flow rate; there was a similar (but non-significant) association with genetically-determined LTL, suggesting longer telomeres enhance respiratory function (**Supplementary Table 5**). A related issue of considerable interest is whether LTL is a determinant of physical fitness. We found associations of usual LTL with different measures of physical activity and with grip strength but no compelling genetic evidence to support causal relationships (**Supplementary Table 5**). Similarly, in women we found associations of longer usual LTL with several traits related to reproductive health, including age of first and last birth and age at menopause, but none was associated with genetically-determined LTL (**Supplementary Table 5**). With respect to traits that may act as proxies for neurological/cognitive function, we observed an association between longer usual LTL and higher age at completion of education, with genetic evidence supportive of a causal association (**Figure 3A**). By contrast, longer usual LTL was associated with prolonged snap button times (a measure of reaction time).

Given interest in associations of LTL with metabolic and endocrine factors that could mediate disease relationships[71], we explored association of LTL with biochemical traits available in UK Biobank (UKB). Genetically-determined LTL was associated with albumin, apolipoprotein A, high density lipoprotein, triglycerides insulin-like growth factor 1 (IGF1), and sex hormone binding globulin (**Figure 3A**). Longer LTL was associated with lower circulating levels of these biomarkers, except for IGF1 and triglycerides.

**Investigating disease association non-concordance.**

We assessed the proportional hazards assumption for all observational models by allowing for time-varying effects. Where proportional hazards were shown to deviate (time interaction $P<0.05$), we estimated the hazard ratios at baseline and at 10 years via linear combination. This analysis showed that the interpretation of our estimates do not change substantially, within the timeframe of the UKB study, even if allowed to vary over time.

For 26 diseases, we found Bonferroni-significant associations with usual LTL but non-significant associations with genetically-determined LTL (**Supplementary Figure 9**). For some of these conditions, the observational associations are likely to reflect residual confounding. For example, heart failure and atrial fibrillation (AF) are often secondary consequences of coronary artery disease (CAD). Adjusting for CAD made the observational association with AF non-significant (OR 1.01 (0.98-1.03), p=0.63) and substantially attenuated the association with heart failure (OR 1.07 (1.03-1.10), $p=5.5x10^{-5}$). On the other hand, for other observational associations, such as with aortic stenosis, osteoporosis and chronic obstructive pulmonary disease, there was a trend towards a genetic association that was directionally consistent with the observational findings (**Supplementary Figure 9**), suggesting the need for greater power in MR analyses. Nevertheless, our power calculations suggest our MR analyses had power to detect an odds ratio of at least 1.10 for these diseases (**Supplementary Figure 13**).

For nine diseases, we found nominal-significant associations with genetically-determined LTL but no associations, or associations in the opposite direction, with usual LTL (**Figure 3B**). Discrepancies could arise from either use of different case definitions (e.g., use of prevalent disease outcomes for genetic analysis vs incident disease outcomes for observational analysis) or residual confounding or both. As an example, we focused on hypertension, an outcome *inversely* associated with shorter LTL in MR analyses, yet *positively* associated with usual LTL. When restricting the MR analysis to incident disease outcomes, the inverse MR association attenuated to the null, suggesting that a disease history was has an effect on the LTL genetic associations (**Supplementary Figure 8**). It is also possible that the change is due to a drop in statistical power. When using prevalent data, the positive observational association also attenuates to the null. After adjusting for several potential confounders, we observed an inverse association with usual LTL, in keeping with MR results (**Supplementary Table 13**).

**Supplementary methods**

**GCTA:** Starting with the most significantly associated SNPs, the model then adds SNPs iteratively in a forward stepwise manner, calculating conditional P-values for all SNPs. LD estimation within GCTA was estimated using a subset of 50,000 samples randomly selected from the UKB. Where a SNP shows evidence of collinearity with another ($R^2$>0.9), the conditional P-value is set to 1. This process is repeated until no further SNPs can be added to the model.

**VEP:** Using VEP[47] gives information on the location of SNPs with respect to genes alongside several measures of functional consequence. Overall, each variant is placed into one of four impact categories. High impact variants include those predicted to ablate or truncate transcripts, alter the reading frame or alter splicing. Moderate variants are mostly predicted to alter protein sequence through addition, loss or change in amino acids, although this class also includes loss of regulatory region variants. Low impact variants include sequence changes that are unlikely to alter function (synonymous changes, start/stop retained) and modifier variants include those that may alter expression levels. CADD scores are returned for all variants and SIFT and PolyPhen annotations are given for variants that lead to amino acid changes. To aid interpretation all results were filtered to the canonical transcript of each gene.

**Gene based score:** Variant annotation was extracted from VEP using the LOFTEE plugin[47,53] Variants were categorised into four groups and scored based on their predicted function. Firstly, ultra-rare potential inactivating variants (MAF<0.01%), i.e. those that were predicted with high confidence to truncate (STOP gain) or radically alter the protein sequence (splice site, frameshift) by the Loss-Of-Function Transcript Effect Estimator (LOFTEE) algorithm, were assigned a score of 1. This corresponds to a complete inactivation of the allele. The same score was applied to rare variants (MAF<0.1%) that were predicted to be pathogenic or likely pathogenic in ClinVar[72] (downloaded 17/12/2020), using the clinical significance annotation ("clinsig" field), where there was no conflicting annotation ('benign,' 'likely benign,' or 'uncertain significance').

We then incorporated ultra-rare variants with a predicted cryptic splice site by SpliceAI a score of 0.75, based on previous literature[73]. The SpliceAI algorithm, using default parameters, was used to estimate four delta scores, DS_AG (acceptor gain delta score), DS_AL (acceptor loss delta score), DS_DG (donor gain delta score), and DS_DL (donor loss delta score). Each of these scores has values ranging from 0 to 1. Variants with a value of any of these four scores ≥0.5 were designated as a cryptic splice site and assigned the score of 0.75.

To increase statistical power we also integrated ultra-rare missense mutations that were predicted to be damaging by all five computational algorithms used (SIFT, Polyphen2-HDIV, Polyphen2-HVAR, LRT, and MutationTaster) as previously described[74,75.] Annotation differs between these algorithms, for clarity we took the following annotations to be "damaging; damaging (SIFT) possibly or probably damaging (Polyphen2-HDIV, Polyphen2-HVAR), deleterious (LRT) and disease-causing-automatic or disease-causing (MutationTaster). As the effect of single amino acid changes are less likely to be as strong as inactivating variants, these variants were assigned a gene-specific weight that took into account the cumulative frequencies of high-confidence loss-of-function variants as compared to predicted damaging missense variants as previously described[75-77]. Given the cumulative allele frequency of all of the LOFTEE high confidence rare variants of a gene (G) as $f_L$, the cumulative allele frequency of all of the predicted damaging missense variants as $f_M$, the weight for the missense variants was estimated as:

$$\left(\frac{f_L \times (1 - f_L)}{f_M \times (1 - f_M)}\right)^{0.5}$$

For each participant an aggregated gene score was computed based on individual variant weightings with a maximum score of 1. Where missense variants were weighted 0 due to $f_L$ = 0 in genes with no high confidence LoF variants, the missense variants were re-assigned a weight of 1.

**Disease codes for health episodes statistics and death registry records:** We defined 123 outcomes based on incident electronic health records data using primary and secondary codes from the 9th and 10th revisions of the international statistical classification of diseases and related health problems (ICD-9 & 10) and the office of population censuses and surveys classification of surgical operations versions 3 and 4 (OPCS-3 & 4) from the UK office of national statistics (ONS). Precise outcome definitions are given in Supplementary Table 11. We grouped these outcomes into 10 broad categories: cancer, cardiovascular disease, digestive diseases, genitourinary diseases, immune/inflammatory diseases, infections, mental illness, musculoskeletal diseases, respiratory diseases, sensory disorders. We generated code lists using available UKB data from both self-reported data (UKB codes for the outcomes obtained from self-reported / verbal interview data gathered at the assessment centre visits) and electronic health records (ICD-9 and ICD-10 codes for use in both hospital episodes statistics (HES) and death registry data, and OPSC-3 and OPSC-4 codes to identify directly linked operations in HES).

A basic HES record is a finished consultant episode of care (the time spent under the care of one consultant). An admission, or spell, is defined as a continuous period of time spent as a patient within a trust and may include more than one episode. We defined first disease occurrence using within-hospital episodes and death records as follows:

| **Disease definition** | ICD-9/10 codes identified from HES | OPCS3/4* operation codes identified from HES | ICD-9/10 codes identified in ONS records |
|---|---|---|---|
| **First recorded hospitalisation or death with any diagnosis of disease of interest** | Primary and secondary diagnosis during a hospital episode | Operation that is directly linked to the disease of interest | Underlying and related cause of death |

*We did not differentiate between main and secondary operations

**Definition of prevalent and incident cases:** We defined the baseline date as the date of sample collection that LTL was measured in, and censored the end of follow-up in hospital health record data as 31/March/2020. This corresponds to the latest UKB release update before the COVID-19 outbreak in the UK. We allocated censor dates on the basis of the location of a participant's assessment centre. Data about deaths is also subject to censoring using the same date of 31 March 2020 used for HES records.

Prevalent cases are defined as self-reported at baseline, or a recorded hospitalisation with any (primary or secondary) diagnosis of the disease of interest before the baseline date from the following sources:

- UKB self-report codes at baseline
- Relevant codes (ICD-9/10 and OPCS3/4) in primary or secondary diagnosis during a hospital episode HES

The rationale for including secondary fields from HES and ONS to define prevalent cases is because of the inclusion of less strict self-report codes from UKB.

Incident cases are defined as the first recorded hospitalisation or death with the disease of interest given as the primary or secondary cause occurring after the UKB baseline visit. Time-to-event is defined as the post-baseline date of the first incident hospitalisation or death, or otherwise censored at the end of study follow-up on 31 March 2020.

**Public health modelling of years' life lost:** We used three pieces of information to estimate differences in life expectancy associated with shorter measured LTL at baseline, categorised into 4 pre-defined groups, henceforth "exposure groups" namely, group 1 (<-1SD), group 2 (-1 to <0 SD), group 3 (0 to <1SD), and group 4 (≥1SD) [Reference group]:

(i) age-at-risk specific hazard ratios for all-cause (and cause-specific) mortality in each exposure group versus the reference (derived from UKB);

(ii) population all-cause (and cause-specific) mortality rates (derived from the EUROSTAT database (http://ec.europa.eu/eurostat/data/database) for UK and 28 EU countries as calculated averages during the three year period 2014-2016 (i.e. 2015 midpoint); and

(iii) prevalence of exposure groups in the population (derived from UKB).

We estimated population survival curves for each exposure group, utilising estimated age-at-risk specific hazard ratios for mortality by exposure groups in UKB and routine statistics on overall population mortality rates. We estimated differences in life-expectancy as differences in areas under any two survival curves compared.

Age-at-risk specific hazard ratios for mortality by exposure groups were estimated from UKB data. Specifically, a Cox regression model stratified by sex was fitted using a dataset in which participant ages-at-risk were deterministically updated by splitting the follow up times every 5-years and recalculating an age-at-risk variable at the beginning of each 5-year interval of follow up. Interactions between baseline exposure groups and linear and quadratic terms for the age-at-risk variable were included in the model to obtain smoothed hazard ratios. Thus, for participant in stratum $s$ with exposure group indicator variable $E_{si(j)}$ (i.e. dummy variable equal to 1 if in exposure group $j$ and zero otherwise) the log hazard rate at time $t$ since baseline was modelled as:

$$\log(h_{si}(t)) = \log(h_{s0}(t)) + \sum_{j=1}^{3} \gamma_{0j} E_{si(j)} + \beta_1 agerisk_{si} + \beta_2 agerisk_{si}^2 + \sum_{j=1}^{3} \gamma_{1j} E_{si(j)} \times agerisk_{si} + \sum_{j=1}^{3} \gamma_{2j} E_{si(j)} \times agerisk_{si}^2 \qquad (1)$$

from which the age-at-risk specific hazard ratios (and 95% CIs) for mortality were obtained as linear combinations of the relevant estimated coefficients, with age-at-risk fixed at values corresponding to midpoints of 5-year age-groups from age 40 onwards (**Supplementary Figure 14**).

Population all-cause (and cause-specific) mortality rates per 100,000 were obtained in 5-year age-groups for the UK and 28 EU countries from the EUROSTAT database (http://ec.europa.eu/eurostat/data/database) as calculated averages during the three year period 2014-2016 (i.e. 2015 midpoint) (**Supplementary Figure 15**). Because the mortality rates were provided only up to age-group 80-84 years, but we desired to estimate the overall population survival curves, we used a piecewise cubic Hermite interpolation (PCHIP) method to smooth through the midpoints of 5-year age-groups and extrapolate the mortality rates to age 110 years (**Supplementary Figure 16**). Next, assuming exponential survival (i.e. constant hazard) within each 5-year age group, we estimated the age-specific survival probability as $S_a = exp(-5 \times IR_a)$ and derived the overall population survival curves from age 35 onwards as the product of the relevant age-group specific survival probabilities (**Supplementary Figure 17**).

$$p(survival|agerisk \geq 35) = \prod_{agerisk \geq 35} S_a \qquad (2)$$

In order to infer population mortality rates appropriate for the reference exposure group used in our estimation of age-specific hazard ratios (i.e. group 4 with standardised LTL ≥1-SD longer than the population mean), we used ordinal logistic regression to model the age-specific prevalence of the four exposure groups in UKB by sex and decade of recruitment (**Supplementary Figure 18**). We used the age-specific prevalence estimates for the decade commencing in the year 2000 to infer the age-specific mortality rates appropriate for our reference group $IR_{a0}$ as:[1]

$$IR_{a0} = \frac{IR_a}{p_{a0} + \sum_{j=1}^{3} p_{aj} \times RR_{aj}} \qquad (3)$$

where $IR_a$ is the population mortality rate for age group $a$, $p_{aj}$ is the age-specific prevalence of exposure group $j$, and $RR_{aj}$ is the age-specific hazard ratio in comparison of exposure group $j$ versus reference group ($j = 0$). The age-specific mortality rates in each of the non-reference exposure groups were then inferred in turn by multiplying the age-specific mortality rate for the reference group $IR_{a0}$ by the age-specific hazard ratios $RR_{aj}$ based on UKB data and equation *(2)* above used to infer the exposure group-specific population survival curves (**Supplementary Figure 19**). Finally, differences in life expectancy according to baseline exposure groups were estimated as difference in the areas under the survival curves for the reference group and each of the non-reference exposure groups in turn (**Supplementary Figure 20**). The areas under curves were calculated by numerical integration.

Monte Carlo simulation was used to calculate confidence intervals for the estimated reductions in life expectancy, taking into account uncertainty in the age-at-risk specific hazard ratios calculated from equation *(1)* above. In particular, new parameter estimates were randomly drawn from the multivariate normal distribution defined by the fitted model mean and covariance matrix, 200 times, and the above procedure repeated for each draw to calculate differences in life-expectancy for each index age of interest. Assuming asymptotic normality, the standard deviation of the 200 Monte Carlo estimates of differences in life expectancy for each index age were used to calculate 95% confidence intervals around the originally estimated value. Histograms were inspected to judge that normality assumption was reasonable.

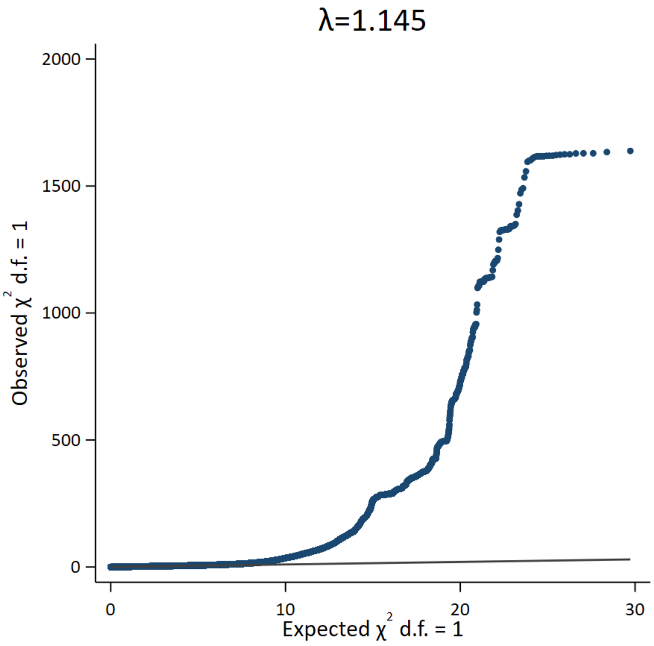


**Supplementary Figure 1. Quantile-quantile plot. Observed versus expected results from the GWAS is plotted. The genomic inflation factor (λ) is also shown.**

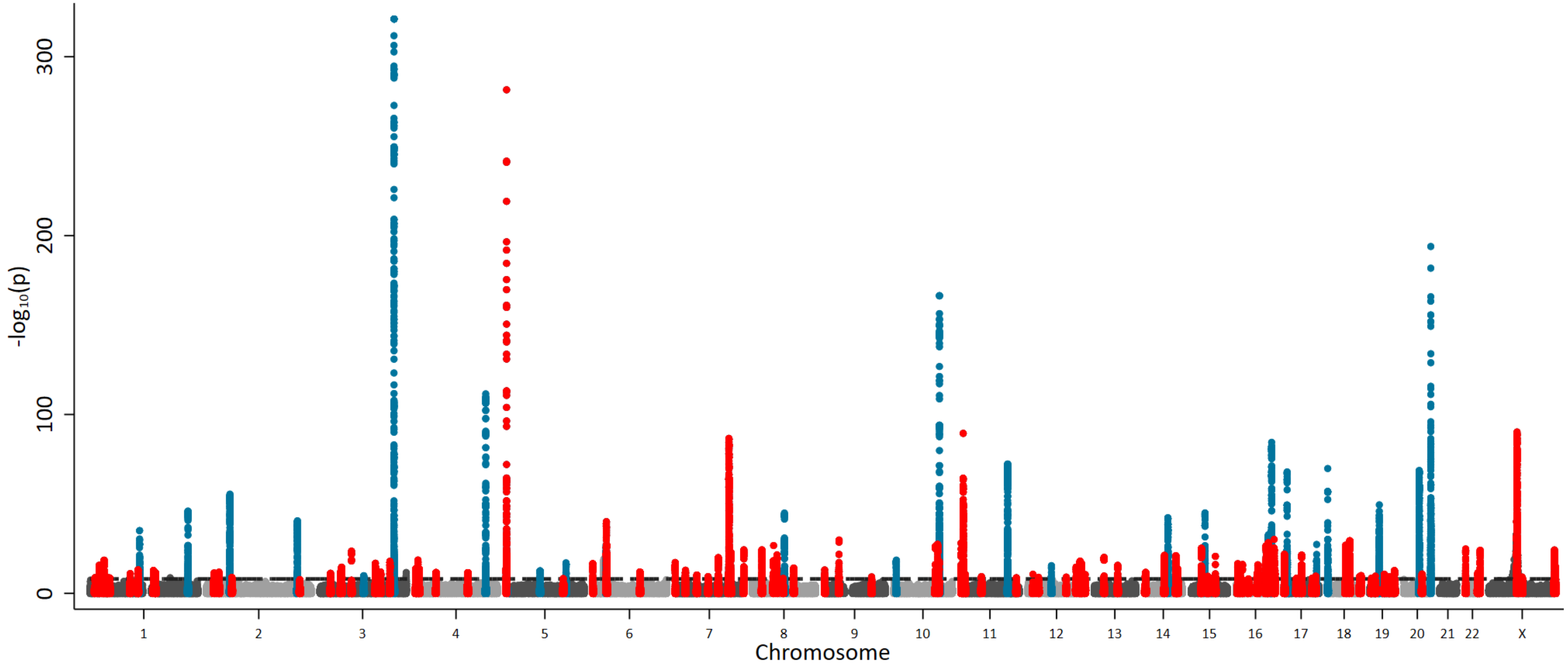


**Supplementary Figure 2. Manhattan plot unrestricted by p-value.** We highlight our 197 sentinel variant regions that are genome-wide significant ($P<8.31x10^{-9}$ horizontal dashed reference line) in the exact joint conditional model (**Supplementary Table 1**). We define the region as known (blue) if a previous variant within 1MB of our sentinel has previously been reported at genome wide significance. We consider our other regions novel (red) as the first evidence of a variant within 1MB of our sentinel that reaches genome wide significance.

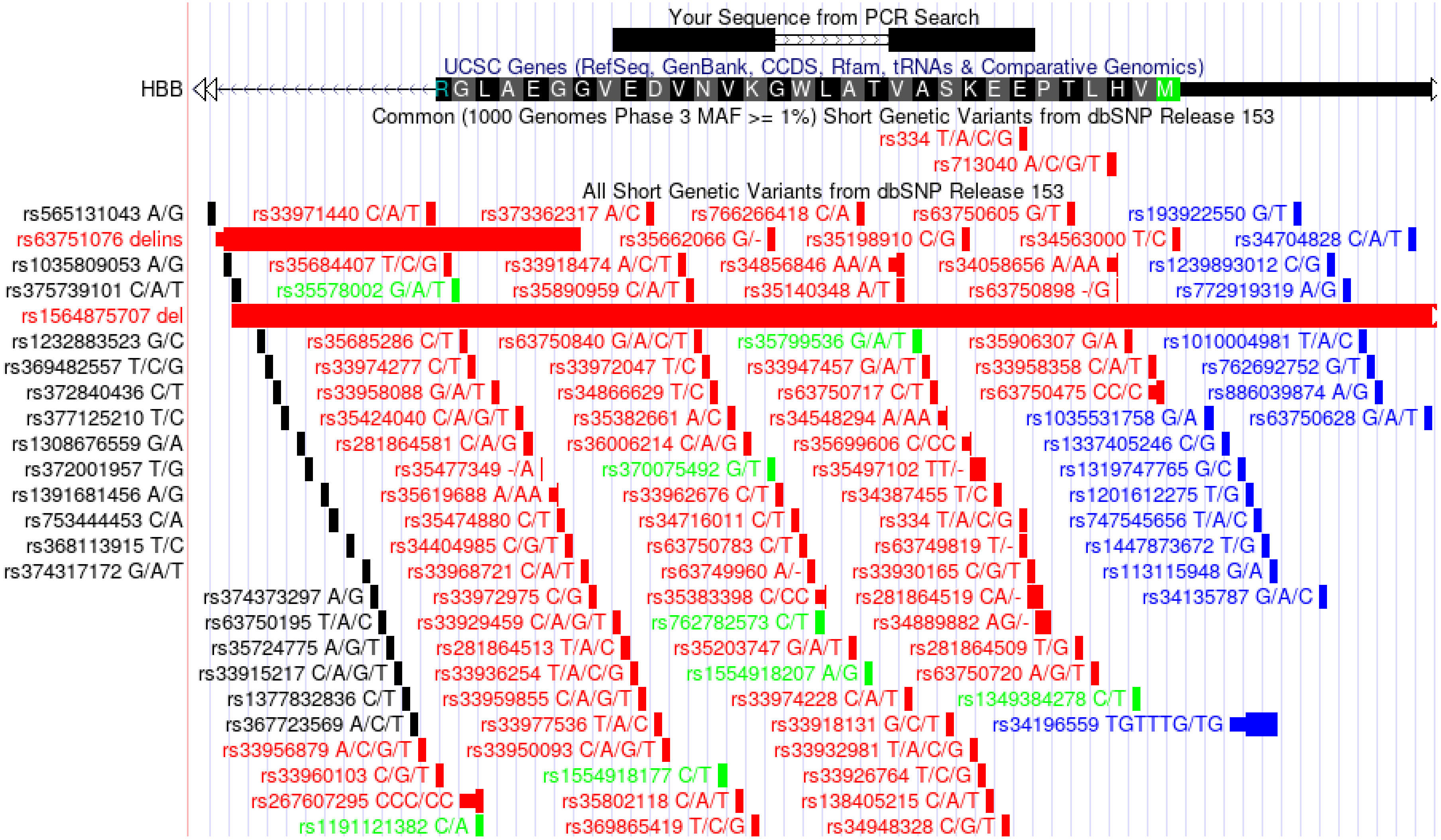


**Supplementary Figure 4. Alignment of single copy PCR gene primers within *HBB*.** Primer sequences are shown as black bars at the top with SNP positions illustrated below. The position of rs334 can be seen to sit just within the reverse primer binding sequence. Anther SNP rs713040 (R2>0.7 to rs1609812) sits just outside the sequence. Primers sequences and SNP positions are visualised using the *In-silico* PCR tool within the UCSC genome browser (https://genome.ucsc.edu)

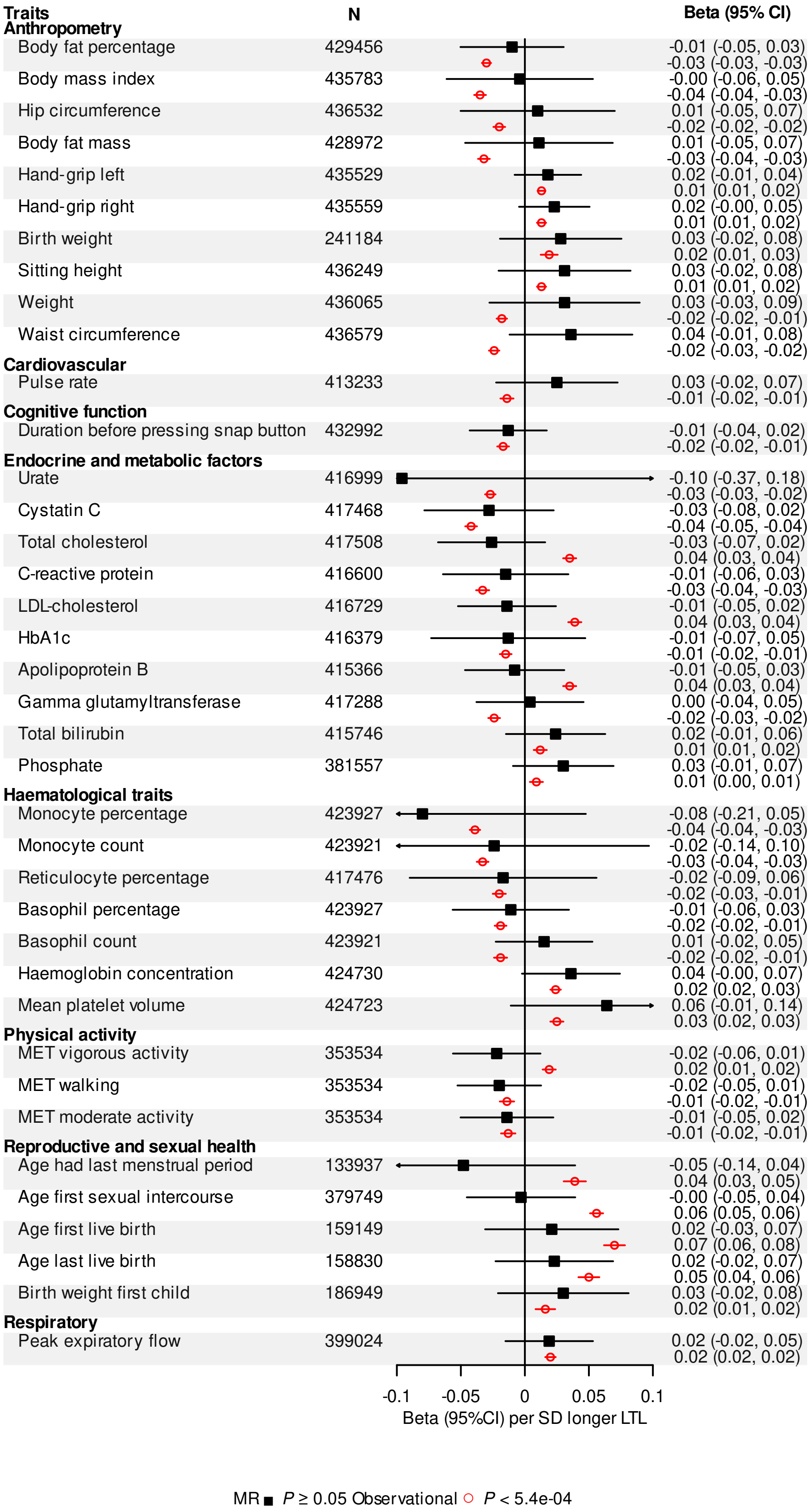


**Qsnnjckclr_pw Dgespc 3, @gmkcbga_j rp_grq _qqmag_rcb ugrf sqs_j JRJ mljw**, Kclbcjg_l p_lbmkgq_rgml &KP' _qqmag_rgmlq _pc qfmul ugrf _ qmjgb qos_pc _lb cvnpcqqcb _q `cr_ ncp qr_lb_pb bctg_rgml &QB' jmlecp eclcrga_jjw bcrcpkglcb jcsamawrc rcjmkcpc jclerf &JRJ', M`qcpt_rgml_j _qqmag_rgmlq _pc qfmul ugrf _l ckmrw agpajc _lb cvnpcqqcb gl QKB ncp QB jmlecp sqs_j JRJ, AG* aml dgbclac glrcpt_j,

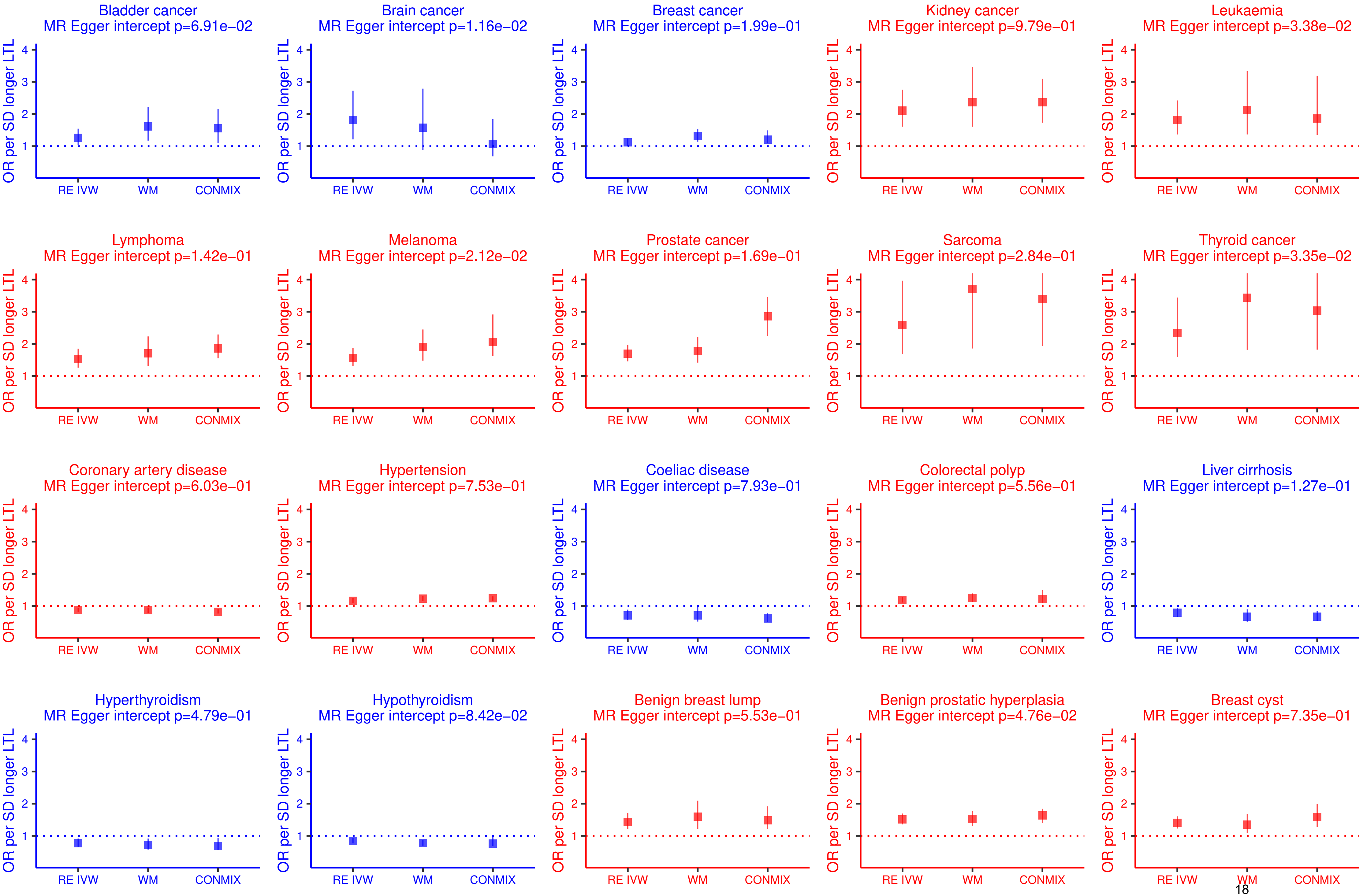

Bladder cancer
MR Egger intercept p=6.91e−02
Brain cancer
MR Egger intercept p=1.16e−02
Breast cancer
MR Egger intercept p=1.99e−01
Kidney cancer
MR Egger intercept p=9.79e−01
Leukaemia
MR Egger intercept p=3.38e−02
Lymphoma
MR Egger intercept p=1.42e−01
Melanoma
MR Egger intercept p=2.12e−02
Prostate cancer
MR Egger intercept p=1.69e−01
Sarcoma
MR Egger intercept p=2.84e−01
Thyroid cancer
MR Egger intercept p=3.35e−02
Coronary artery disease
MR Egger intercept p=6.03e−01
Hypertension
MR Egger intercept p=7.53e−01
Coeliac disease
MR Egger intercept p=7.93e−01
Colorectal polyp
MR Egger intercept p=5.56e−01
Liver cirrhosis
MR Egger intercept p=1.27e−01
Hyperthyroidism
MR Egger intercept p=4.79e−01
Hypothyroidism
MR Egger intercept p=8.42e−02
Benign breast lump
MR Egger intercept p=5.53e−01
Benign prostatic hyperplasia
MR Egger intercept p=4.76e−02
Breast cyst
MR Egger intercept p=7.35e−01
OR per SD longer LTL
1
2
3
4
RE IVW
WM
CONMIX

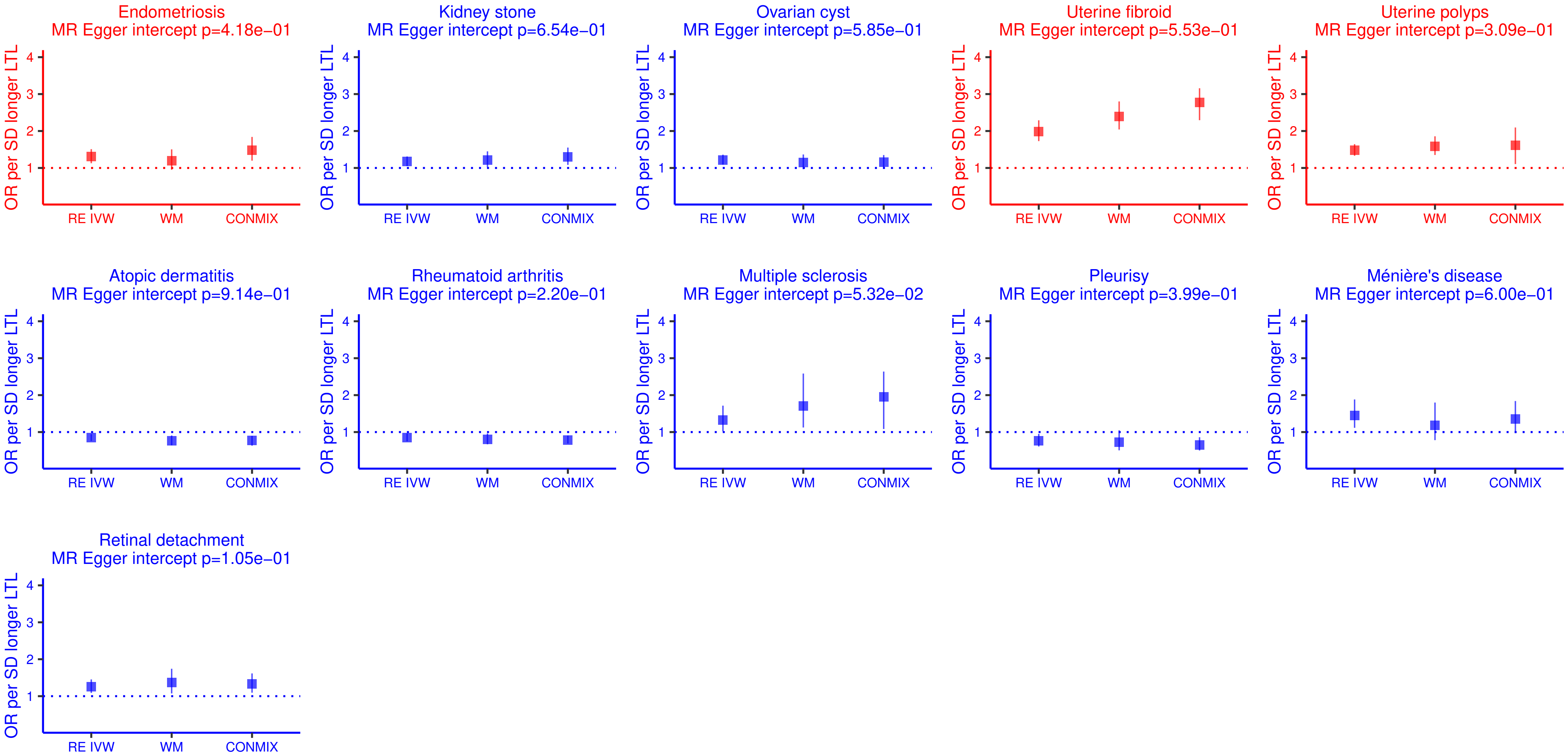


**Supplementary Figure 6. Mendelian randomisation sensitivity analysis for genetically determined LTL and diseases.** Estimates as OR per SD shorter genetically determined LTL produced from the inverse-variance weighted (IVW), weighted median (WM) and contamination mixture (conmix) methods are shown for comparison for each disease. The P-value for the MR Egger intercept is stated as an indicator of potential pleiotropy.

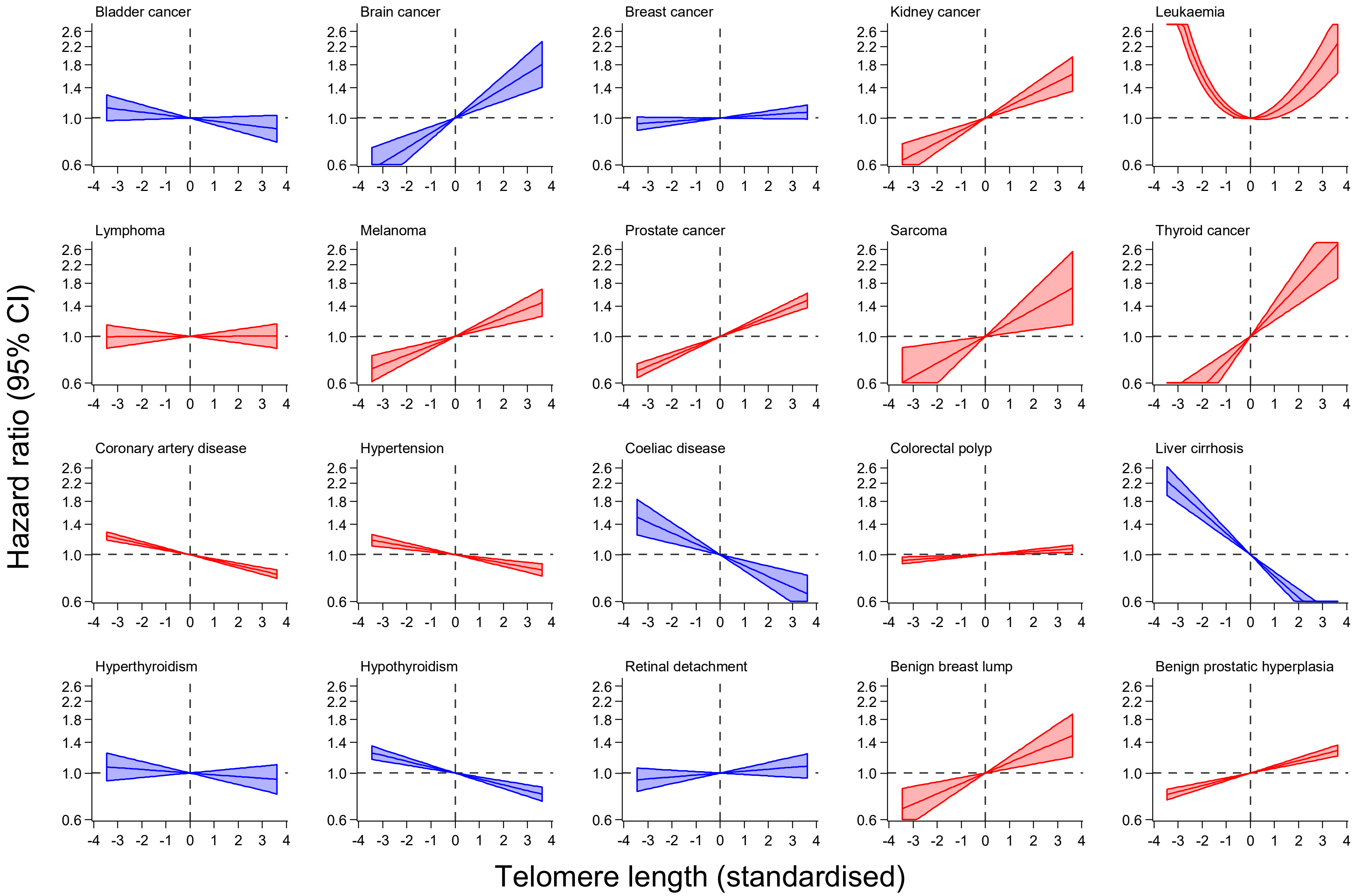

Bladder cancer
Brain cancer
Breast cancer
Kidney cancer
Leukaemia
Lymphoma
Melanoma
Prostate cancer
Sarcoma
Thyroid cancer
Coronary artery disease
Hypertension
Coeliac disease
Colorectal polyp
Liver cirrhosis
Hyperthyroidism
Hypothyroidism
Retinal detachment
Benign breast lump
Benign prostatic hyperplasia
Hazard ratio (95% CI)
Telomere length (standardised)

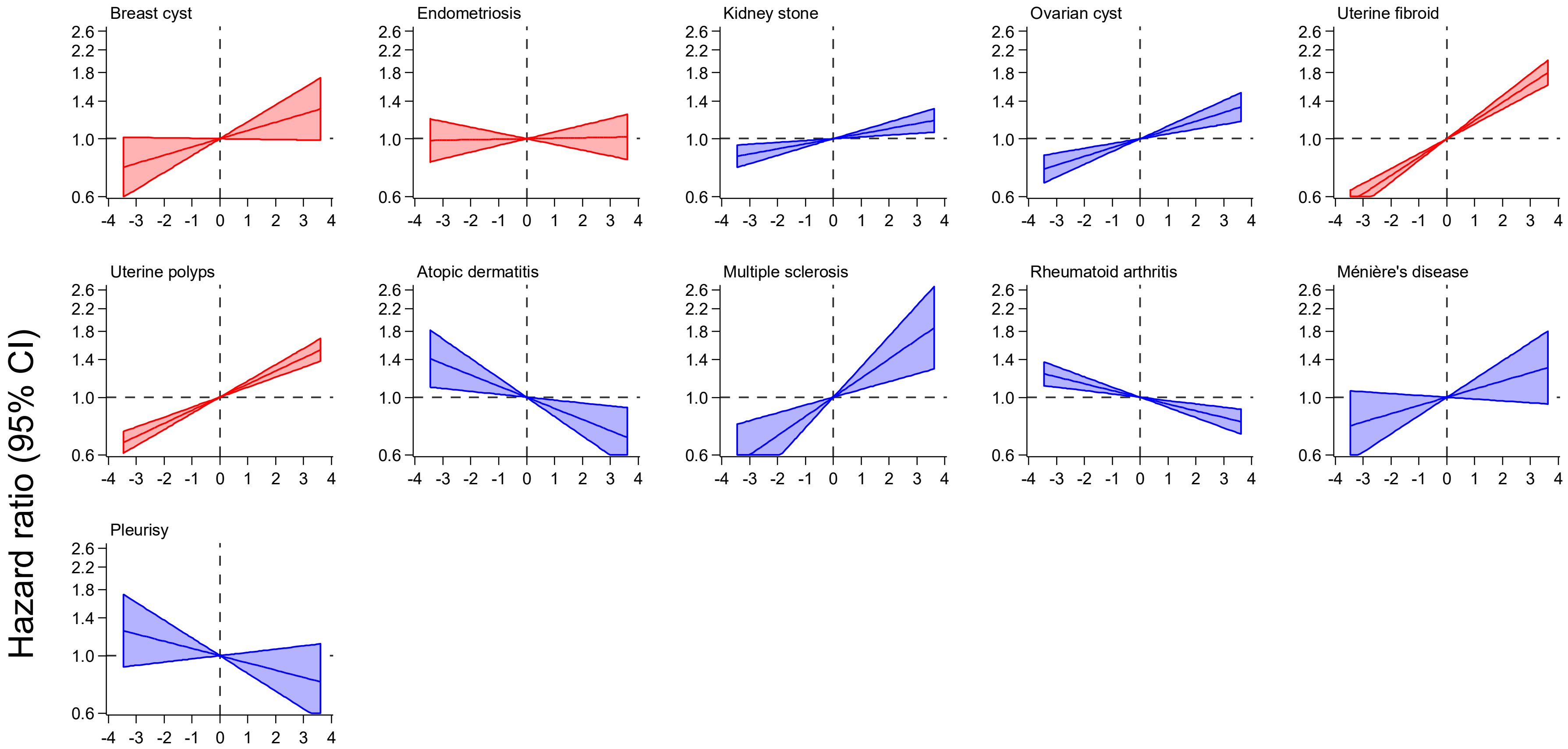


**Supplementary Figure 7. Assessing potential non-linear disease associations with LTL.** Fractional polynomial models were fitted for each disease, adjusting for age, sex, WBC and ethnic group. The continuous shape of each association (relative to a reference value of 0) is shown for each disease. Diseases that were associated with genetically determined LTL in the MR analyses at a Bonferroni level of significance ($P<4.1x10^{-4}$) are shown in red, those that were nominally significant ($P<0.05$) in blue.

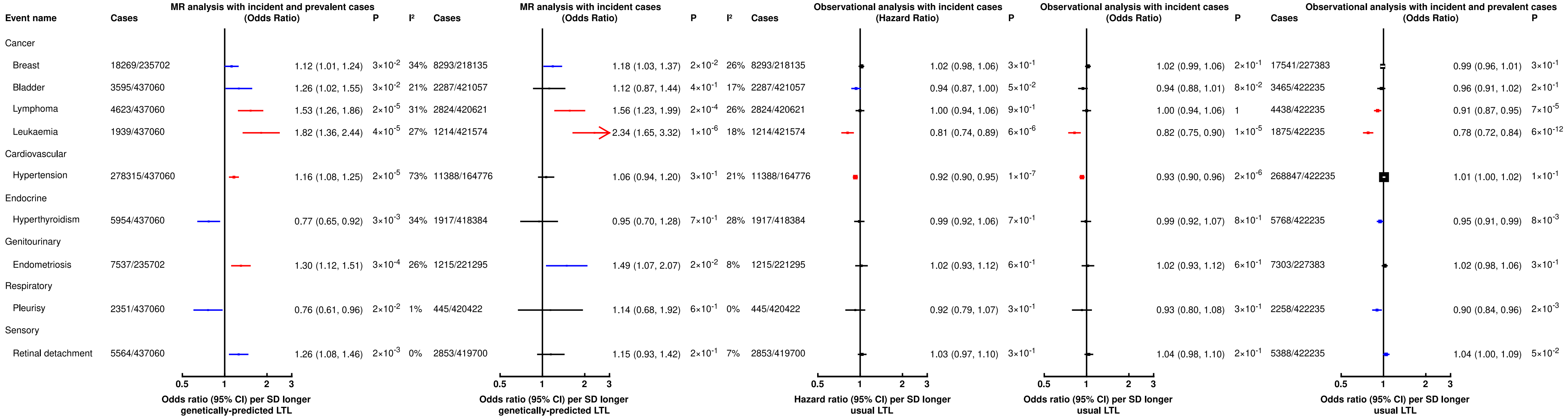


**Supplementary Figure 8. MR and observational analyses with different case definitions and statistical analysis approaches**. Left to right: (i) MR analysis with prevalent and incident analysis (main MR analysis, included here as a reference), (ii) MR analysis restricted to incident cases, (iii) observational Cox regression analysis restricted to incident cases (main observational analysis), (iv) observational logistic regression analysis restricted to incident cases, (v) observational logistic regression with incident and prevalent cases.

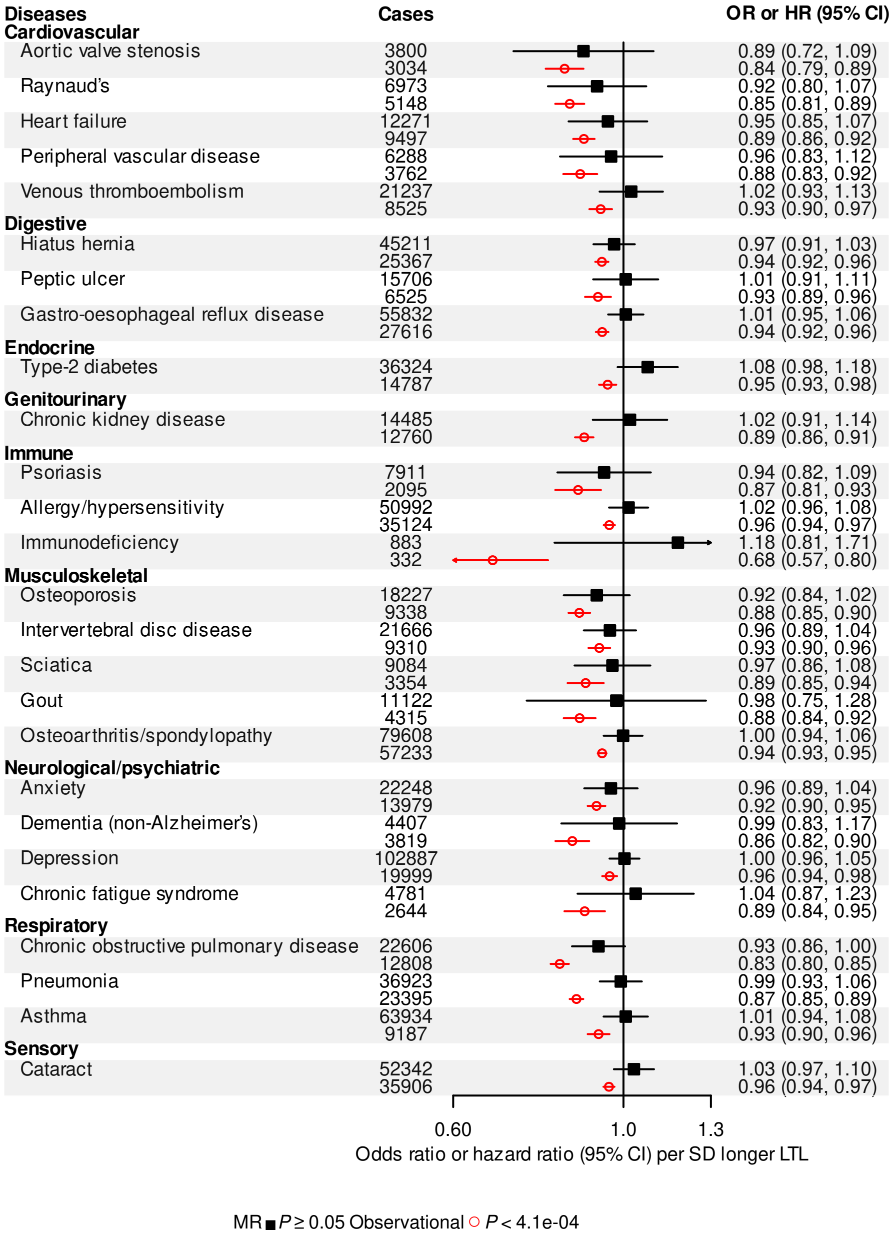


**Supplementary Figure 9. Diseases associated with usual LTL only.** Mendelian randomisation (MR) associations are shown with a solid square and expressed in odds ratio (OR) per standard deviation (SD) longer genetically-determined leucocyte telomere length (LTL). Observational associations are shown with an empty circle and expressed in hazard ratio (HR) per SD longer usual LTL. CI, confidence interval.

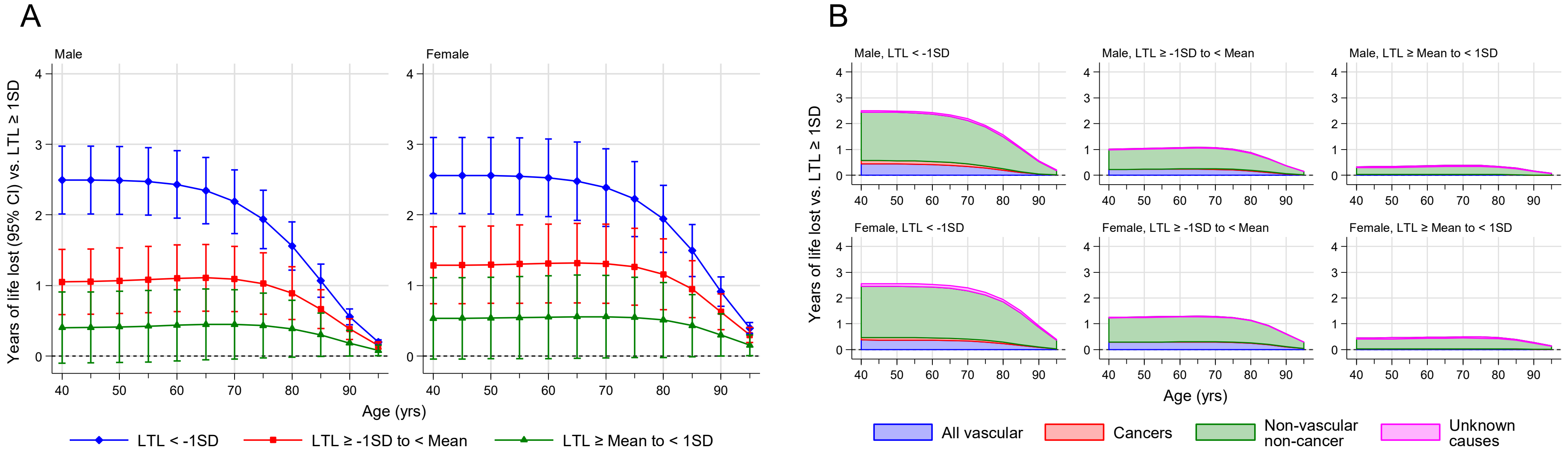


**Supplementary Figure 10. Years of life lost using EU 2015 mortality rates.** Years of life lost were estimated for four standardised LTL groups (group 1 (<-1SD), group 2 (-1 to <0 SD), group 3 (0 to <1SD), and group 4 (≥1SD)) from 40-95 years of age. Group 4 were used as the reference group. Data is shown for males and females separately. This was performed for all cause mortality (A) and disease specific mortality (B).

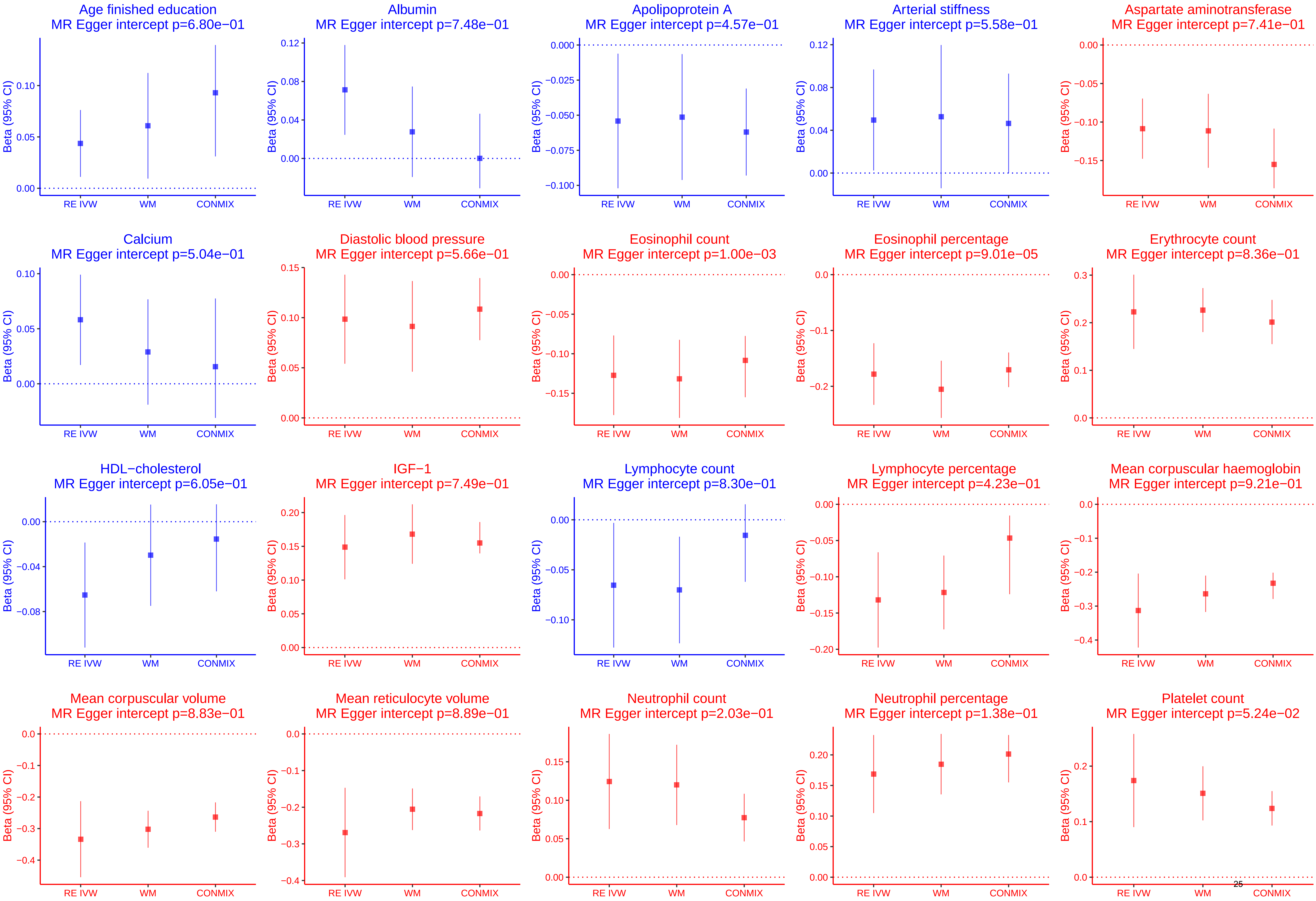

Age finished education
MR Egger intercept p=6.80e−01
Albumin
MR Egger intercept p=7.48e−01
Apolipoprotein A
MR Egger intercept p=4.57e−01
Arterial stiffness
MR Egger intercept p=5.58e−01
Aspartate aminotransferase
MR Egger intercept p=7.41e−01
Calcium
MR Egger intercept p=5.04e−01
Diastolic blood pressure
MR Egger intercept p=5.66e−01
Eosinophil count
MR Egger intercept p=1.00e−03
Eosinophil percentage
MR Egger intercept p=9.01e−05
Erythrocyte count
MR Egger intercept p=8.36e−01
HDL−cholesterol
MR Egger intercept p=6.05e−01
IGF−1
MR Egger intercept p=7.49e−01
Lymphocyte count
MR Egger intercept p=8.30e−01
Lymphocyte percentage
MR Egger intercept p=4.23e−01
Mean corpuscular haemoglobin
MR Egger intercept p=9.21e−01
Mean corpuscular volume
MR Egger intercept p=8.83e−01
Mean reticulocyte volume
MR Egger intercept p=8.89e−01
Neutrophil count
MR Egger intercept p=2.03e−01
Neutrophil percentage
MR Egger intercept p=1.38e−01
Platelet count
MR Egger intercept p=5.24e−02
Beta (95% CI)
RE IVW
WM
CONMIX

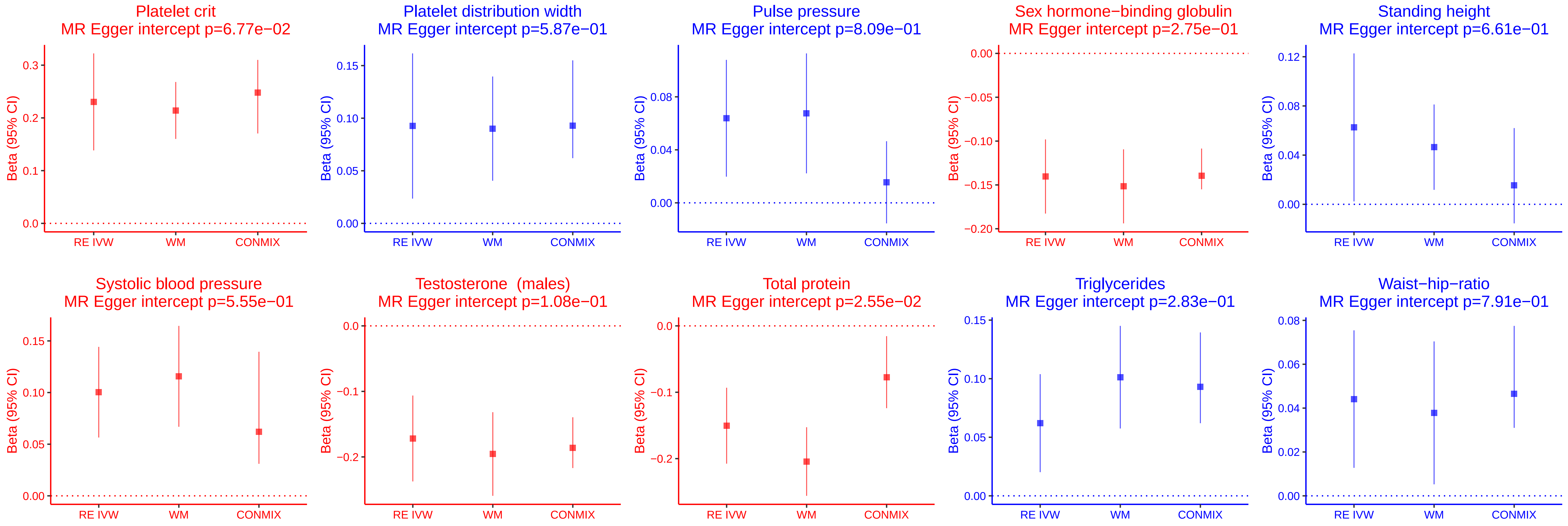


**Supplementary Figure 11. Mendelian randomisation sensitivity analyses for genetically determined LTL associations with biomedical traits**. Estimates as beta per SD shorter genetically determined LTL produced from the inverse-variance weighted (IVW), weighted median (WM) and contamination mixture (conmix) methods are shown for comparison for each disease. The P-value for the MR Egger intercept is stated as an indicator of potential pleiotropy.

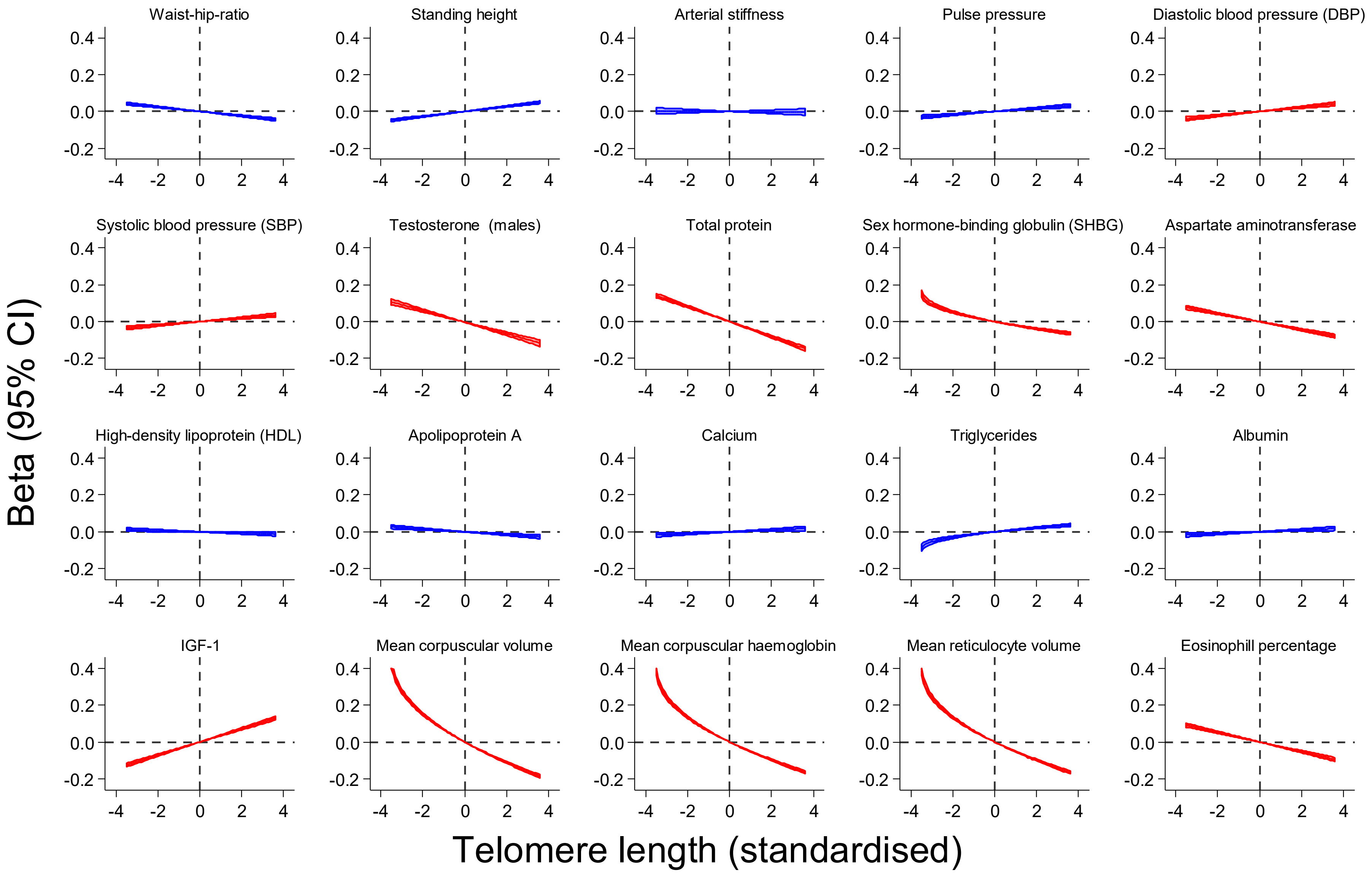
Waist-hip-ratio
Standing height
Arterial stiffness
Pulse pressure
Diastolic blood pressure (DBP)
Systolic blood pressure (SBP)
Testosterone (males)
Total protein
Sex hormone-binding globulin (SHBG)
Aspartate aminotransferase
High-density lipoprotein (HDL)
Apolipoprotein A
Calcium
Triglycerides
Albumin
IGF-1
Mean corpuscular volume
Mean corpuscular haemoglobin
Mean reticulocyte volume
Eosinophill percentage
Beta (95% CI)
Telomere length (standardised)
0.4
0.2
0.0
-0.2
-4
-2
0
2
4

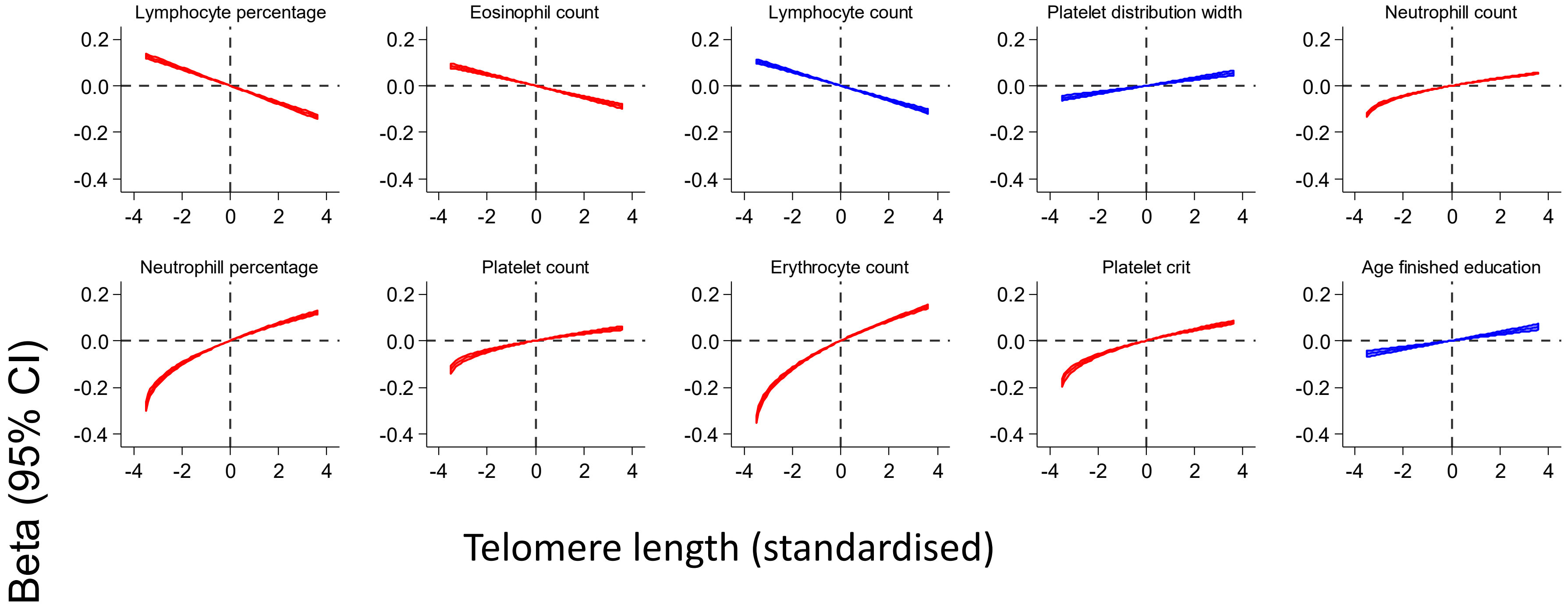


**Supplementary Figure 12. Assessing potential non-linear biomedical trait associations with LTL.** Fractional polynomial models were fitted for each biomedical trait, adjusting for age, sex, WBC and ethnic group. The continuous shape of each association (relative to a reference value of 0) is shown for each disease. Biomedical traits that were associated with genetically determined LTL in the MR analyses at a Bonferroni level of significance ($P<5.38x10^{-4}$) are shown in red, those that were nominally significant ($P<0.05$) in blue.

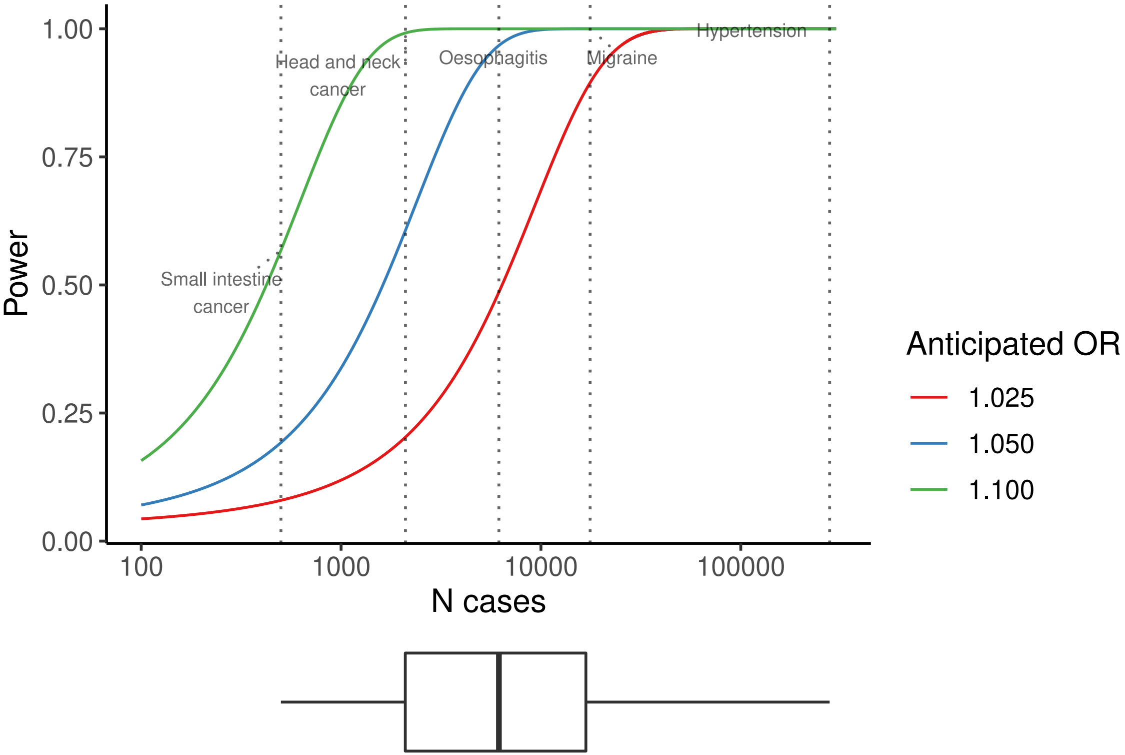


**Supplementary Figure 13. Power calculations for Mendelian randomization disease analysis.** Annotated diseases reflect the distribution of case counts, shown in the above boxplot: minimum number of cases (small intestine cancer), bottom quartile (head and neck cancer), median (oesophagitis), upper quartile (migraine), and maximum (hypertension). OR, odds ratio.

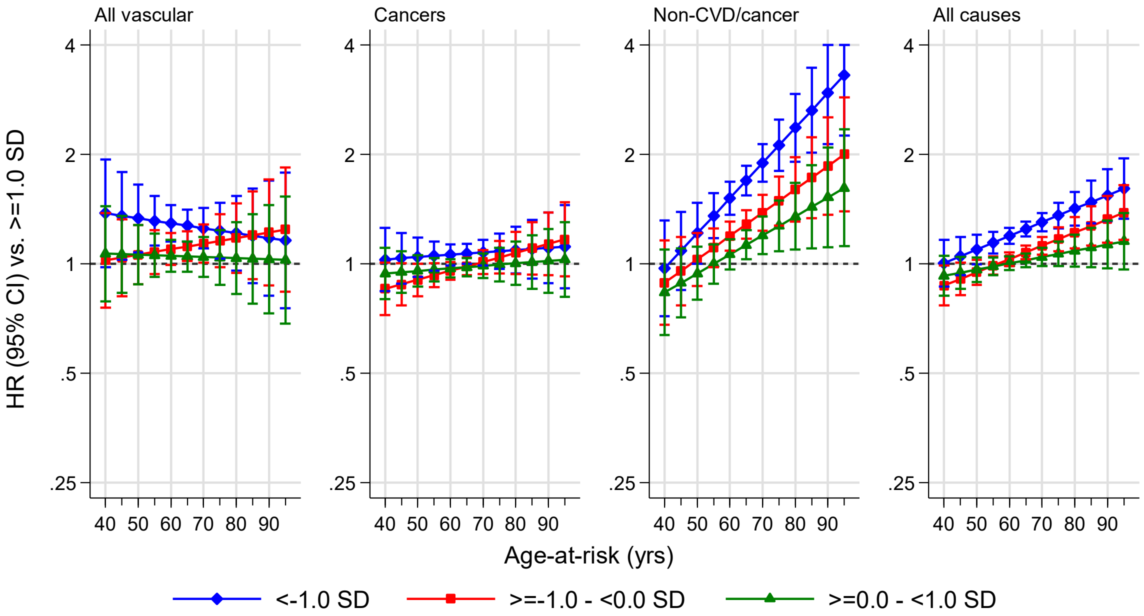


**Supplementary Figure 14. Age-at-risk specific hazard ratios for all-cause mortality by sex and LTL exposure groups estimated from UKB data.**

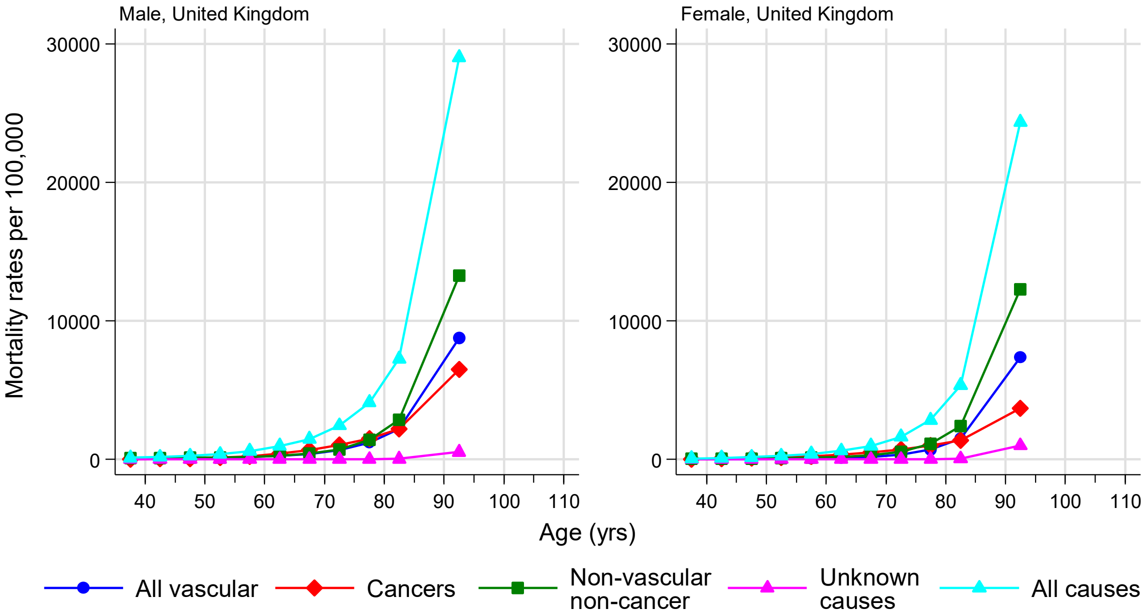


**Supplementary Figure 15. UK population mortality rates during year 2015 downloaded from EUROSTAT online database**. To maintain consistency with analyses conducted in UKB, the mortality rate for non-vascular non-cancer causes was recalculated as the difference of all-cause mortality and the sum of vascular mortality (I00-I99), cancer mortality (C00-D48), and unknown causes of mortality (R00-R99).

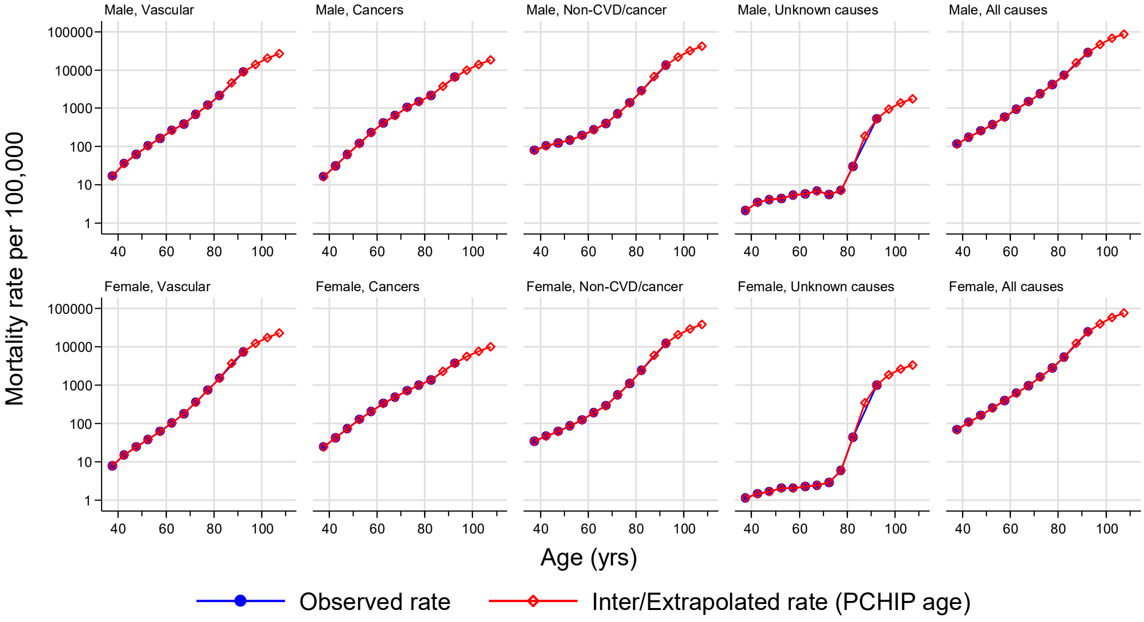


**Supplementary Figure 16. Assessment of adequacy of a piecewise cubic Hermite interpolation (PCHIP) method to smooth and extrapolate UK population mortality rates during 2015**. Data was downloaded from EUROSTAT online database beyond the database's upper bound age cut-off of 84 years to facilitate modelling.

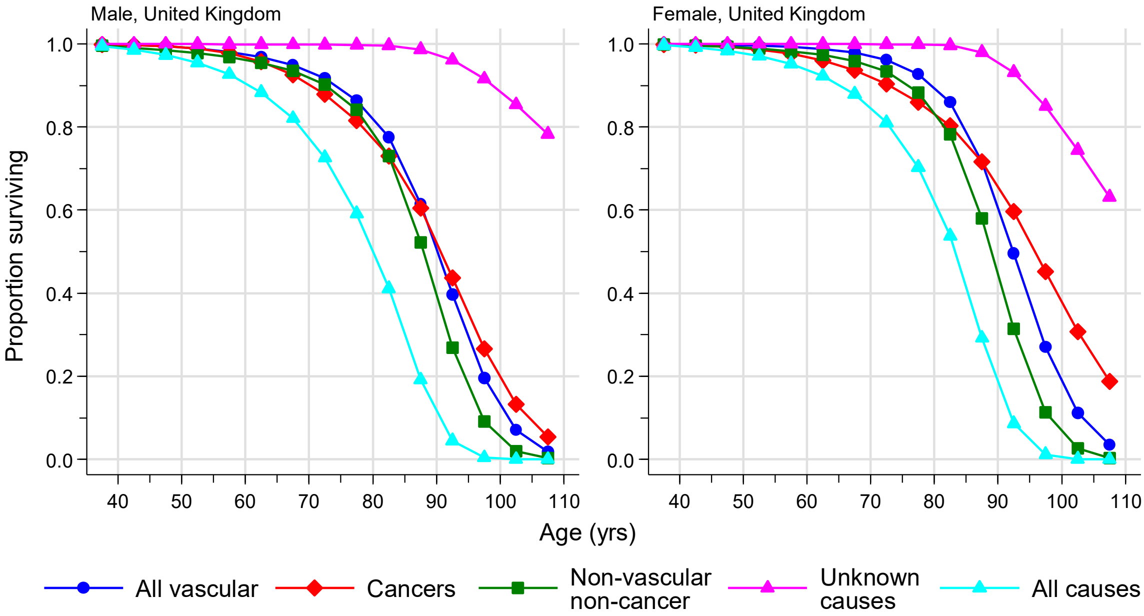


**Supplementary Figure 17.** Derived population survival curves for all-cause and cause-specific mortality from age 35 years based on smoothed and extrapolated UK population mortality rates during 2015.

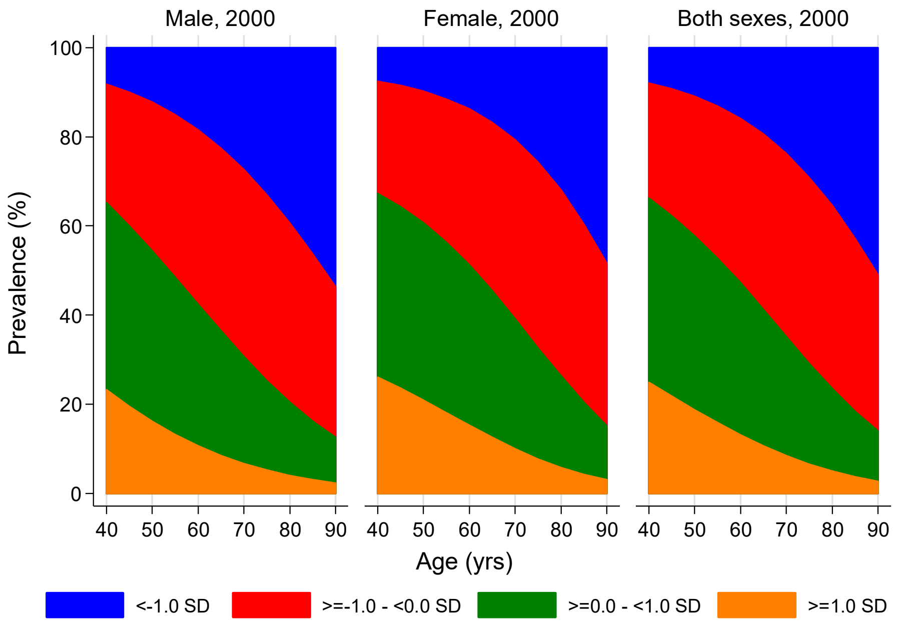


**Supplementary Figure 18.** Modelled age-specific prevalence of LTL exposure groups in UKB by sex and decade of recruitment.

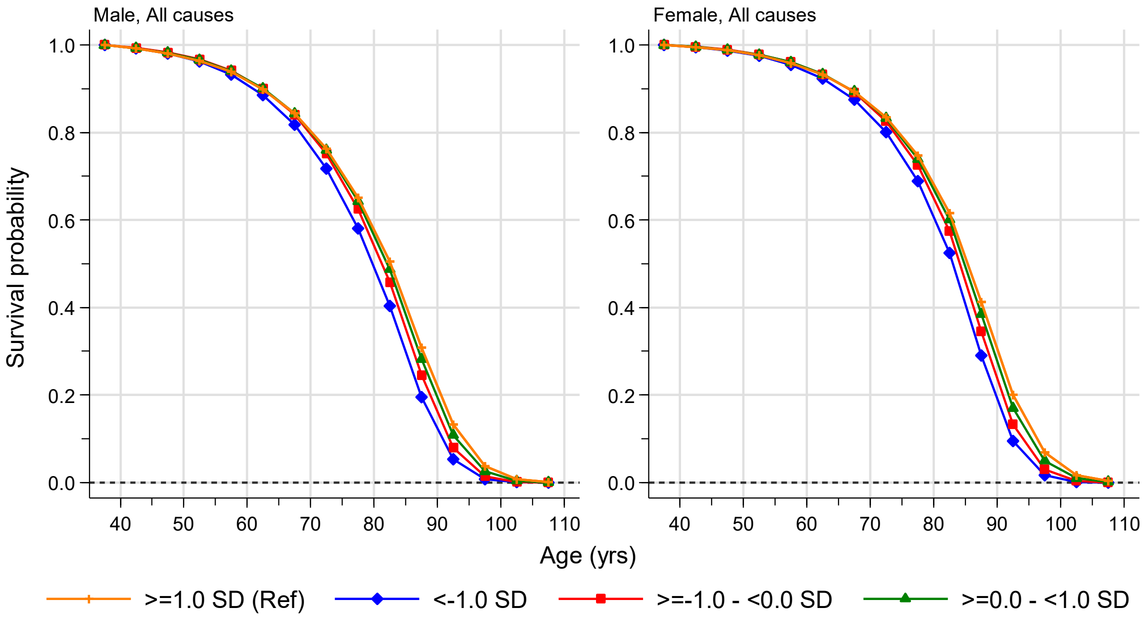


**Supplementary Figure 19. Inferred survival curves for UK population by sex and LTL exposure groups.**

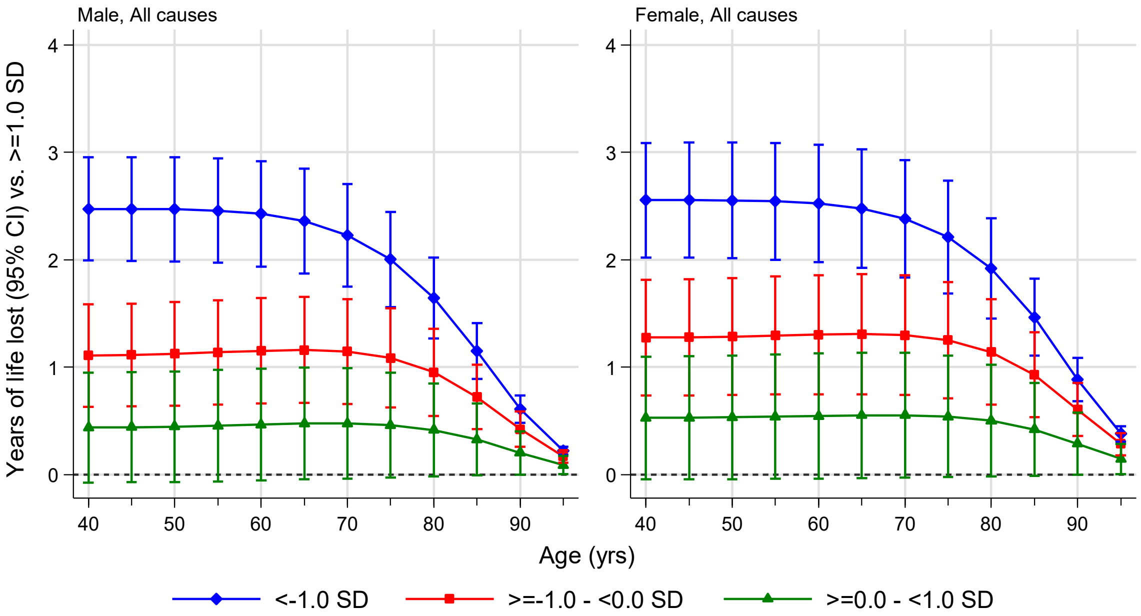


**Supplementary Figure 20. Estimated sex-specific reductions in life expectancy in the UK population according to LTL exposure groups.**
